# Defining the Clinical Complexity of hEDS and HSD: A Global Survey of Diagnostic Challenge, Comorbidities, and Unmet Needs

**DOI:** 10.1101/2025.06.05.25329074

**Authors:** Victoria Daylor, Molly Griggs, Amy Weintraub, Rebecca Byrd, Taylor Petrucci, Matthew Huff, Kathryn Byerly, Roman Fenner, Sydney Severance, Charlotte Griggs, Amol Sharma, Paldeep Atwal, Steven A. Kautz, Steven Shapiro, Kimberly Youkhana, Mark Lavallee, Allison Wilkerson, Michelle Nichols, Alan Snyder, Josef Eichinger, Sunil Patel, Anne Maitland, Cortney Gensemer, Russell A. Norris

## Abstract

**Background:** Hypermobile Ehlers-Danlos syndrome (hEDS) and hypermobility spectrum disorders (HSD) are connective tissue disorders marked by chronic pain, joint instability, and extensive multisystem involvement. Despite affecting an estimated 1 in 500 individuals, these conditions remain poorly understood, and current diagnostic categories lack clarity and consistency. This study aimed to characterize the clinical presentation, comorbidities, and healthcare burden of individuals with hEDS and HSD through a large-scale international survey.

**Methods and Findings:** A cross-sectional, anonymous online survey was distributed globally between September 2023 and March 2024. Of 9,258 responses, 3,906 participants met inclusion criteria and were included in analysis. The 418-item questionnaire covered symptom domains, diagnoses, healthcare access, and quality of life. Responses were statistically analyzed using chi-square and Mann-Whitney U tests with Bonferroni correction and compared to the All of Us dataset (n=354,400). Participants reported high rates of gastrointestinal disorders (84.3% hEDS, 69.0% HSD), with 21.2% of hEDS participants diagnosed with gastroparesis and 7.1% requiring feeding devices. Dysautonomia was common, affecting 71.4% of those with hEDS and 40.3% with HSD (p<0.0001), with postural orthostatic tachycardia syndrome (POTS) as the most frequently reported subtype. Neurological complications were also prevalent, with hEDS participants reporting significantly higher rates of tethered cord syndrome (4.6%, 0.9%), Chiari malformation (6.3%, 0.9%), and small fiber neuropathy (10.0%, 4.4%) compared to those with HSD. Chronic pain was nearly universal (98.8% hEDS, 92.7% HSD). On average, hEDS participants reported 24 comorbid diagnoses and HSD participants 17, with diagnostic delays averaging over 20 years. Notably, 50% of those reporting HSD met hEDS criteria, while 26% of those reporting hEDS did not meet full criteria, underscoring persistent diagnostic uncertainty.

**Conclusions:** This global survey underscores the profound multisystemic burden, diagnostic ambiguity, and unmet clinical needs faced by individuals with hEDS and HSD. The high prevalence of immune-mediated, neurological, gastrointestinal, and autonomic dysfunctions, alongside the frequent identification of triggering events such as infections and hormonal transitions, challenges the conventional framework that defines these disorders as purely connective tissue in origin. Instead, our findings support the hypothesis that hEDS and/or HSD may represent complex syndromes in which connective tissue fragility may be a downstream consequence rather than the primary cause. This reframing has critical implications for diagnosis, pathophysiology, and therapeutic development, and highlights the need for mechanistic studies that explore distinct etiologies beyond the connective tissue paradigm.

## Introduction

The Ehlers-Danlos syndromes (EDS) are a group of 14 heritable connective tissue disorders (CTDs) characterized by varying phenotypic presentation, often sharing commonalities of joint hypermobility, skin hyperextensibility, and ligament and tissue fragility (1, 2). This group of disorders is heterogeneous with most subtypes arising from mutations affecting collagen or other components of the extracellular matrix (ECM) (2, 3). While the majority of subtypes have clearly defined genetic markers, the most common subtype, hypermobile Ehlers-Danlos syndrome (hEDS), lacks a clear genetic or diagnostic test. As such, hEDS is diagnosed clinically based on the 2017 criteria established by the International Consortium on The Ehlers-Danlos Syndromes and Related Disorders. The diagnostic evaluation is structured as a clinician-administered checklist comprised of three criteria: 1) a minimum age-based score on the Beighton scale to confirm generalized joint hypermobility, 2) clinical evidence of a connective tissue disorder, including a minimum number of clinical characteristics, chronic pain, and/or a family history, and 3) exclusion of alternative diagnoses (2). Patients who demonstrate symptomatic joint hypermobility but do not meet the 2017 diagnostic threshold for hEDS are often diagnosed with hypermobility spectrum disorders (HSD). HSD can include generalized, peripheral, localized, or historical symptomatic joint hypermobility, and has yet to be clearly defined clinically as the etiology remains unknown (4). Based on a recent study, the combined diagnosed prevalence of hEDS and HSD is estimated at 1 in 500 individuals (5), making these diseases prevalent in the community.

Clinically, hEDS and HSD often present with overlapping features, including musculoskeletal complaints, extensive systemic involvement, and a range of comorbid conditions (6). This may include neurological, immunological, cardiovascular, endocrine, genitourinary, dental, and gastrointestinal (GI) components (7–9). Symptom onset typically occurs in early adolescence, with patients commonly reporting an increase in injuries, GI disturbances, and joint pain which are often mistaken for growing pains (10, 11). A triggering event, such as environmental, emotional, or viral factors, is often reported near the onset of symptoms (12). Patients may also experience overlapping comorbid conditions, including autonomic dysfunction, mast cell activation disease (MCAD), including mast cell activation syndrome (MCAS), and cardiovascular manifestations such as mitral valve prolapse or aortic root dilatation (13). Neurological complications like cerebrospinal fluid (CSF) leaks, cranio-cervical instability (CCI), tethered cord syndrome (TCS), and small fiber neuropathy (SFN) are also reported (14). Other structural manifestations include pelvic and rectal prolapse, hiatal hernias, Chiari malformation and spinal instabilities (8, 9, 15–17).

While these conditions are increasingly recognized in patients with hEDS and HSD, the full spectrum of associated manifestations remains poorly characterized. Many patients experience a lengthy journey to diagnosis due to the lack of a diagnostic test, barriers within the healthcare system, physician knowledge and attitudes, and the complexity of the clinical presentations (18). This “diagnostic odyssey” can result in delays in care, reduced quality of life and worsening of symptoms over time without access to proper management (12, 19).

Early perspectives helped shape the diagnostic criteria for these conditions, but they lacked robust data. Current clinical insights are primarily based on small datasets without control groups and limited in their scope to a few comorbidities or single-organ manifestations (2, 20–22). To bridge this gap, this study conducted a large-scale global survey of individuals with hEDS and HSD to provide a more comprehensive, statistically driven view of their multisystemic clinical presentation. This report provides a comprehensive assessment of the prevalence of hEDS symptoms and associated comorbidities, as well as their impact on patients’ daily lives. Our findings aim to inform clinical decision-making and guide more effective, patient-centered care strategies for this complex and heterogeneous population.

## Materials And Methods

### Ethics Approval

The study was reviewed by the Medical University of South Carolina Institutional Review Board (IRB) and determined to be exempt under category 2.

### Survey Design

This study employed a cross-sectional survey design to assess patients with hEDS or HSD using self-reported data collected via RedCap. The survey was designed through an iterative collaboration with research staff, physicians specializing in treating EDS patients, and a cohort of patients with hEDS, with the goal of successfully capturing a broad scope of the patient experience of complex chronic illnesses. The anonymous survey consisted of 194 questions covering 192 conditions, along with associated symptoms (**Supplementary** Fig 1 and 2). The survey was open for six months, from September 2023 to March 2024. Participants were recruited through social media, online forums, medical providers, word of mouth, and community organizations worldwide. To be eligible, participants had to be at least 18 years old and have a formal diagnosis of EDS or HSD.

### Statistical Analyses

Survey results (n=9258) were directly imported from RedCap using the RedCapTidieR package in R (23). As shown in **Fig 1**, we filtered for individuals who reported a diagnosis of hEDS or HSD, were at least 18 years old, and completed the full survey (n=5,653). Their self-reported hEDS and HSD diagnoses were validated using the first two criteria of the 2017 hEDS Diagnostic Criteria (2) through an R script. To mitigate bias in self-reported responses, diagnostic criteria questions were deliberately dispersed throughout the questionnaire to reduce the likelihood that participants would recognize and tailor their answers to the diagnostic framework. For Criterion 1, participants aged 18-50 with a Beighton score of ≥5 and those over 50 with a Beighton score of ≥4 were considered to have met the criterion. For Criterion 2, Features A, B, and C were assessed, with participants meeting at least two of these categories considered as satisfying the criterion. Criterion 3 was not applied due to the inability to verify physical examination findings. This process yielded two analysis groups: hEDS patients meeting the 2017 hEDS diagnostic criteria (hEDS group), and HSD patients who did not meet the hEDS criteria (HSD group). Those who reported hEDS but did not meet the criteria (n=1183) and those reporting HSD who did meet hEDS criteria (n=564) were excluded from analyses. No duplicates were identified. Due to the prevalence of certain conditions in the dataset, similar answer choices (e.g., “irritable bowel syndrome – Constipation” and “irritable bowel syndrome – Mixed”) were grouped as a single disorder for analysis unless otherwise specified.

**Fig 1.**
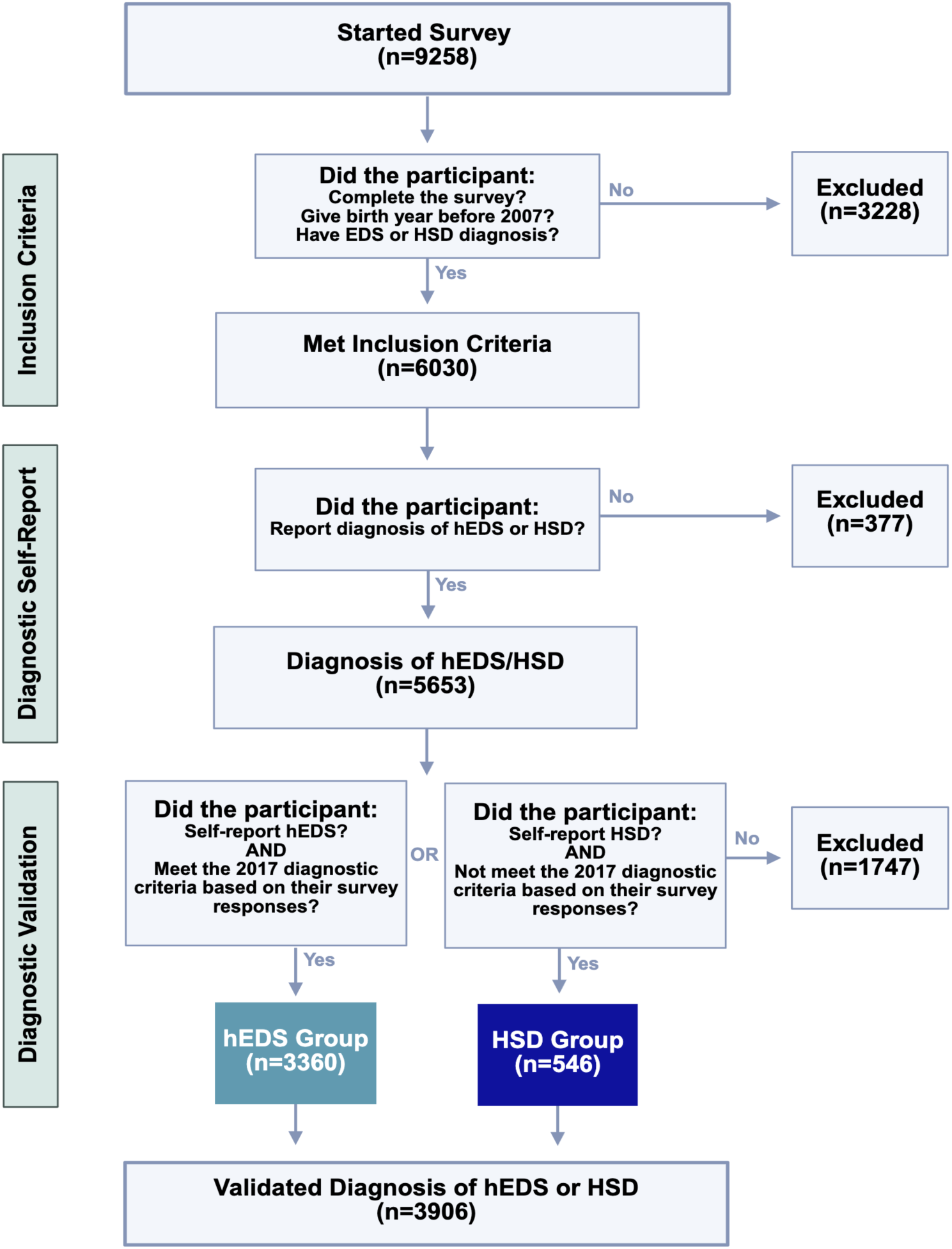
Inclusion criteria and diagnostic validation steps for survey respondents. Of 9,258 individuals who began the survey, participants were progressively excluded for age, incomplete data, or inconsistent diagnoses. Final groups (hEDS: n=3,360; HSD: n=546) reflect participants who both self-reported and met criteria consistent with diagnostic classification.

A subset of survey questions was selected for statistical analysis, with additional exclusions applied to items that had fewer than five responses, following the “Rule of 5” for normal approximation (24). To compare hEDS and HSD groups, categorical variables were analyzed using Chi-square tests. Contingency tables were constructed for 418 survey items, and Bonferroni correction was applied to adjust for multiple comparisons. A corrected p-value of ≤ 0.05 was considered statistically significant.

To evaluate the prevalence of comorbidities in the hEDS group relative to the general population, aggregate-level data from the All of Us data browser (n=354,400) were used. This study used data from the All of Us Research Program’s Controlled Tier Dataset 7, available to authorized users on the Researcher Workbench. Survey items related to formal medical diagnoses were included in this comparison if the hEDS group had ≥5 responses and the All of Us dataset had ≥40 responses. Chi-square analyses were performed with Bonferroni correction applied to 138 comparisons, using a significance threshold of p ≤ 0.05. For continuous variables, outliers were identified using the ROUT method (Q = 1%) in GraphPad prior to conducting Mann-Whitney U tests on the cleaned dataset. A p-value of ≤ 0.05 was used to define statistical significance.

To visualize patterns of co-occurring conditions in hEDS patients, a co-morbidity matrix was constructed for conditions that were reported as formal diagnosis. Responses marked “None of the Above” were excluded. The matrix was normalized using the Jaccard Index, which quantified the degree of overlap between each pair of co-morbidities by dividing the number of patients with both conditions by the total number of patients with either condition. For clarity, only the most frequently overlapping conditions were included in the final heatmap, ensuring legibility. Self-overlapping conditions and redundant pairings were removed, resulting in a triangular matrix format. The heatmap was generated using the ComplexHeatmaps package (25, 26).

## Results

### Study Cohort

#### Eligibility Criteria

A total of 9,258 participants initially started the survey. However, 3,228 individuals were excluded for incomplete responses, not meeting age requirements, or failing to report an EDS or HSD diagnosis. This left 6,030 participants who met the initial inclusion criteria to participate. Among these, 5,653 participants reported a diagnosis of hEDS or HSD. Those who did not report such a diagnosis (377 individuals) were excluded from this analysis. The remaining participants underwent a diagnostic validation process using the 2017 criteria. Those who self-reported hEDS and met the criteria based on survey responses were placed in hEDS group (n=3,360), while those who self-reported HSD but did not meet the 2017 criteria were categorized as Group 4 (n=546). However, 1,747 participants who did not meet the necessary diagnostic criteria were excluded from analysis: those who reported hEDS but did not meet the 2017 criteria (n=1,183) and those who reported HSD diagnosis but did meet the 2017 criteria for hEDS (n=564). Ultimately, the final validated sample consisted of 3,906 participants who had either a validated diagnosis of hEDS (hEDS group) or HSD (HSD group). This process ensured that only individuals meeting the established diagnostic criteria were included in the final dataset.

#### Demographics

The survey sample population was primarily composed of younger and middle-aged adults, with the largest proportion (54.8% hEDS, 58.1% HSD) falling between the ages of 26 and 44 (**Fig 2A**). Following in prevalence were younger adults (18–25 years) (19.9% hEDS, 17.4% HSD) and those aged 45–59 (20.4% hEDS, 20.7% HSD). Older adults (60+) represented only a small subset of the sample (4.9% hEDS, 3.8% HSD), potentially reflecting barriers to diagnosis or participation in online research among older populations.

**Fig 2.**
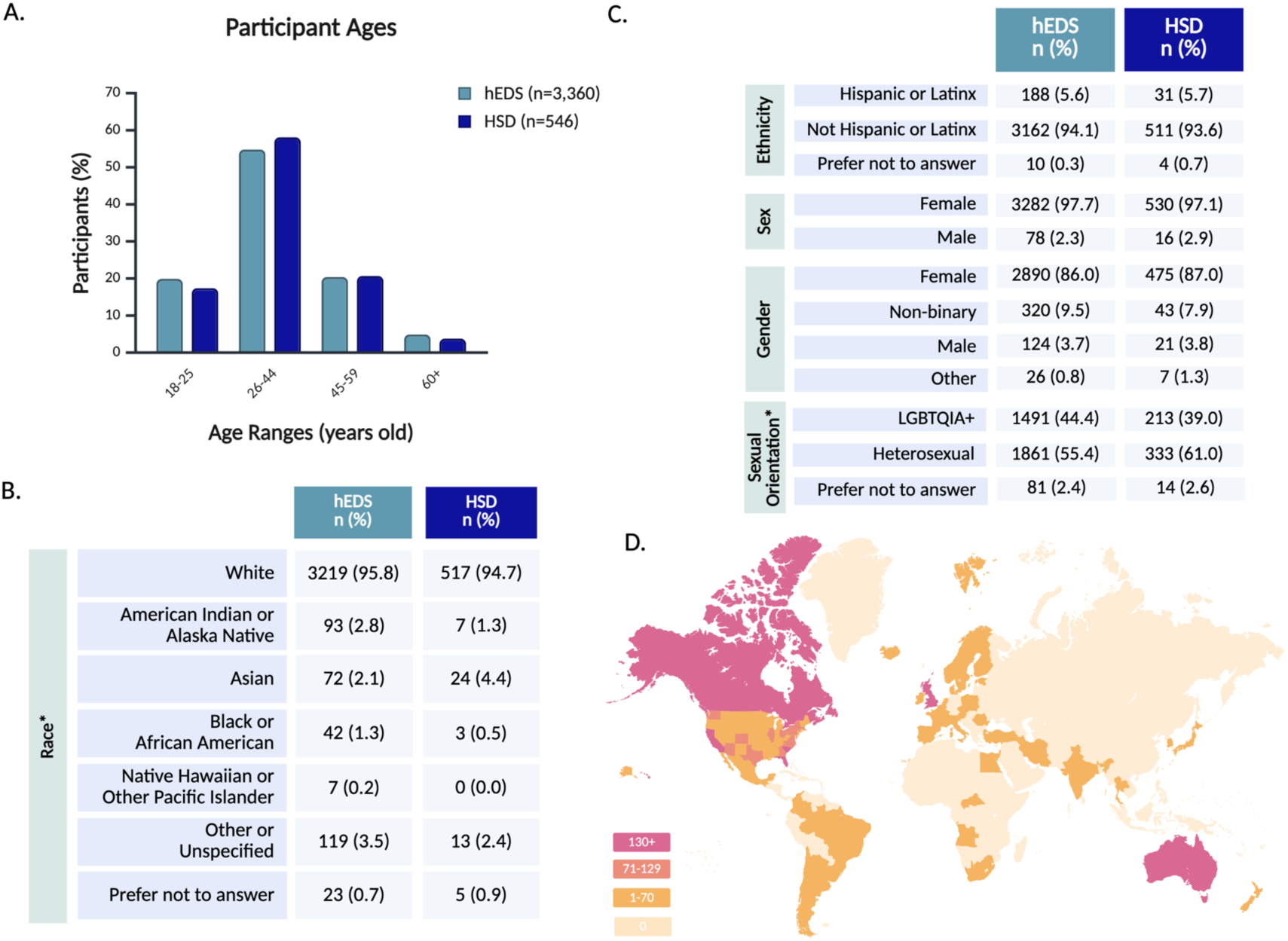
Participant demographics and global representation. Participant demographics are shown across age (A), race (B), and identity-related variables including ethnicity, sex, gender, and sexual orientation (C). Panel D depicts the geographic distribution of participants by country and U.S. state, with darker shading indicating higher regional response counts.

Racial demographics indicated a predominantly White sample (95.8% hEDS, 94.7% HSD), with smaller proportions identifying as American Indian/Native Alaskan (2.8% hEDS, 1.3% HSD), Asian (2.1% hEDS, 4.4% HSD), Black/African American (1.3% hEDS, 0.5% HSD), and Native Hawaiian or other Pacific Islander (0.2% hEDS, 0.0% HSD) (**Fig 2B**). Ethnicity data indicated that a minority of participants identified as Hispanic or Latinx (5.6% hEDS, 5.7% HSD), while the majority identified as not Hispanic or Latinx (94.1% hEDS, 93.6% HSD) (**Fig 2C**). This relative lack of racial diversity mirrors broader disparities in EDS and HSD diagnosis, as studies suggest these conditions are underrecognized in non-White populations (27).

The majority of participants were assigned female at birth (97.7% hEDS, 97.1% HSD), aligning with prior research that highlights a diagnostic bias toward females (15, 28). In terms of gender, 86.0% of hEDS and 87.0% of HSD participants identified as female, with a higher rate of non-binary participants (9.5% hEDS, 7.9% HSD) than male participants (3.7% hEDS, 3.8% HSD). While the majority of participants were cisgender females, gender diversity within the cohort was notable, with 12.0% of all respondents identifying with a gender different from their sex assigned at birth (**Supplementary Table 1**). Sexual orientation among respondents was highly diverse, with a large proportion identifying as part of the LGBTQIA+ spectrum (44.4% hEDS, 39.0% HSD), while the remainder identified as heterosexual (55.4% hEDS, 61.0% HSD) (**Fig 2C and Supplementary Table 2**).

Geographically, 67.5% of all respondents resided in the United States, with representation from every state, including notable participation from California (5.0%), South Carolina (3.5%), Florida (3.5%), and Texas (3.3%) (**Fig 2D and Supplementary Table 3**). Globally, the survey reached participants on every continent, besides Antarctica, with strong response rates from the United Kingdom (13.0%), Australia (4.4%), Canada (4.4%), New Zealand (1.6%), and the Netherlands (1.5%) (**Supplementary Table 4**). This worldwide engagement highlights the survey’s reach across diverse geographic and healthcare landscapes.

### Diagnostic Journey

On average, participants with hEDS reported their symptom onset at the age of 9.3 but were diagnosed at the age of 31.4, with a delay in diagnosis from time of symptom onset of 22.1 years (**Fig 3**). Participants with HSD reported later symptom onset at the age 12.3 (*p*<0.0001) but were diagnosed at the same age as hEDS participants (31.4 years old), leading to a shorter, though still lengthy, delay of 17.5 years from symptom onset to diagnosis. Both hEDS and HSD groups indicated their initial symptoms were musculoskeletal (62.9% hEDS, 73.6% HSD), GI (13.3% hEDS, 7.1% HSD), or autonomic (5.7% hEDS, 4.8% HSD) (**Supplementary Table 5**). The first suspicion of hEDS/HSD came most often from medical doctors (58.6%), followed by physical therapists (14.3%), and themselves or a friend/family member (6.5%). Participants with hEDS were significantly more likely to be diagnosed by geneticists than patients with HSD (36.7%, 14.8%, *p*<0.0001). Rheumatologists were the most common diagnosticians for HSD (39.4%), and the second most common for hEDS (26.5%). Prior to diagnosis, more than half of participants report being misdiagnosed with conditions including fibromyalgia, anxiety, and growing pains (60.1% hEDS, 51.6% HSD).

**Fig 3.**
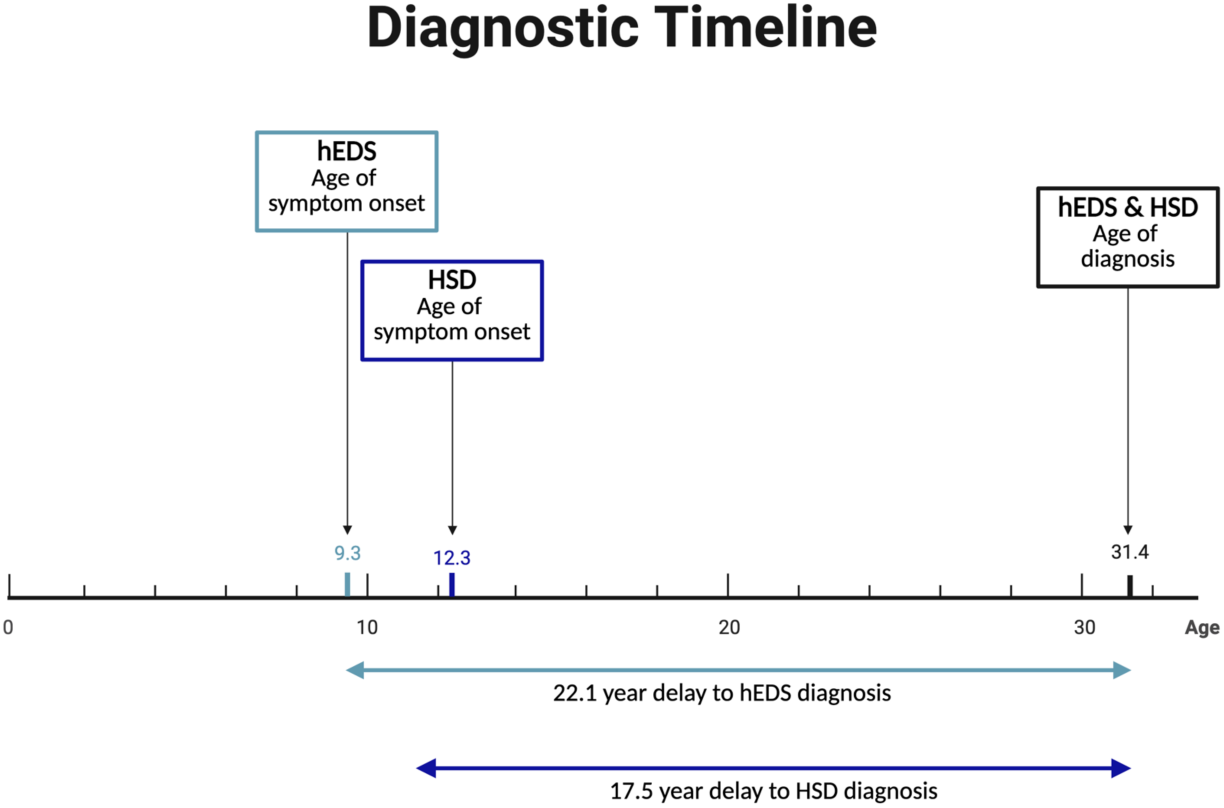
Prolonged diagnostic delays in hEDS and HSD. Despite early symptom onset—9.3 years for hEDS and 12.3 years for HSD—diagnoses were typically not made until age 31.4, resulting in average diagnostic delays of 22.1 and 17.5 years, respectively.

Notably, 70.1% of individuals with hEDS and 65.0% of those with HSD identified a specific event preceding or heightening their symptoms (**Table 1**). Among those with hEDS, the most cited triggers were puberty (30.0% hEDS, 21.6% HSD) (*p*<0.05) and viral or bacterial infections (25.6% hEDS, 23.1% HSD), followed by physical trauma (16.4% hEDS, 13.7% HSD), pregnancy (14.2% hEDS, 9.2% HSD), and psychological or emotional events (14.3% hEDS, 15.8% HSD). Notably a diagnosis of long COVID was reported by 6.6% of participants with hEDS and 6.2% of those with HSD with a substantial number of those participants receiving their long COVID diagnosis prior to being diagnosed with hEDS/HSD (35.7% hEDS, 50.0% HSD).

**Table 1.** Symptom-onset Triggers and COVID-related Responses in hEDS and HSD.

|  | hEDS<br>n (%) | HSD<br>n (%) | hEDS vs. HSD<br>p-Value† |
| --- | --- | --- | --- |
| Yes, event preceded symptoms of hEDS/HSD | 2355 (70.1) | 355 (65) | ns |
| No | 1005 (29.9) | 191 (35) | ns |
| Event type |  |  |  |
| Puberty | 1009 (30) | 118 (21.6) | 0.0293 |
| Viral or bacterial infection | 860 (25.6) | 126 (23.1) | ns |
| Physical accident | 551 (16.4) | 75 (13.7) | ns |
| Psychological or emotional event | 481 (14.3) | 86 (15.8) | ns |
| Pregnancy | 477 (14.2) | 50 (9.2) | ns |
| Other | 470 (14) | 68 (12.5) | ns |
| Diagnosed with Long COVID | 221 (6.6) | 34 (6.2) | ns |
| Diagnosed with Long COVID after hEDS/HSD | 142 (4.2) | 17 (3.1) | ns |
| Diagnosed with Long COVID prior to hEDS/HSD | 79 (2.4) | 17 (3.1) | ns |

hEDS N=3,360; HSD N=546; ns, not significant
† p values were calculated using Chi-square tests, with Bonferroni correction applied to adjust for multiple comparisons.

Participants reported a high burden of co-occurring conditions, with hEDS participants averaging 24 and HSD participants 17, based on 192 conditions assessed in the survey (**Fig 4A and Supplementary** Fig 2). Participants reported experiencing a range of symptoms related to dysautonomia (99.9% hEDS, 98.9% HSD), dermatological issues (99.6% hEDS, 93.2% HSD), and gastrointestinal concerns (99.4% hEDS, 98.5% HSD) (**Fig 4B**). When asked to rank the three most severe symptom categories from a provided list, responses were weighted by rank to reflect overall severity. Chronic pain was most frequently ranked as the most severe symptom (32.6% hEDS, 34.2% HSD), with notable differences between groups: hEDS participants more often prioritized gastrointestinal symptoms (13.5% hEDS, 9.6% HSD), while HSD participants more frequently ranked joint-related symptoms as more severe (12.2% hEDS, 14.1% HSD) (**Table 2, Supplementary Table 6**).

**Fig 4.**
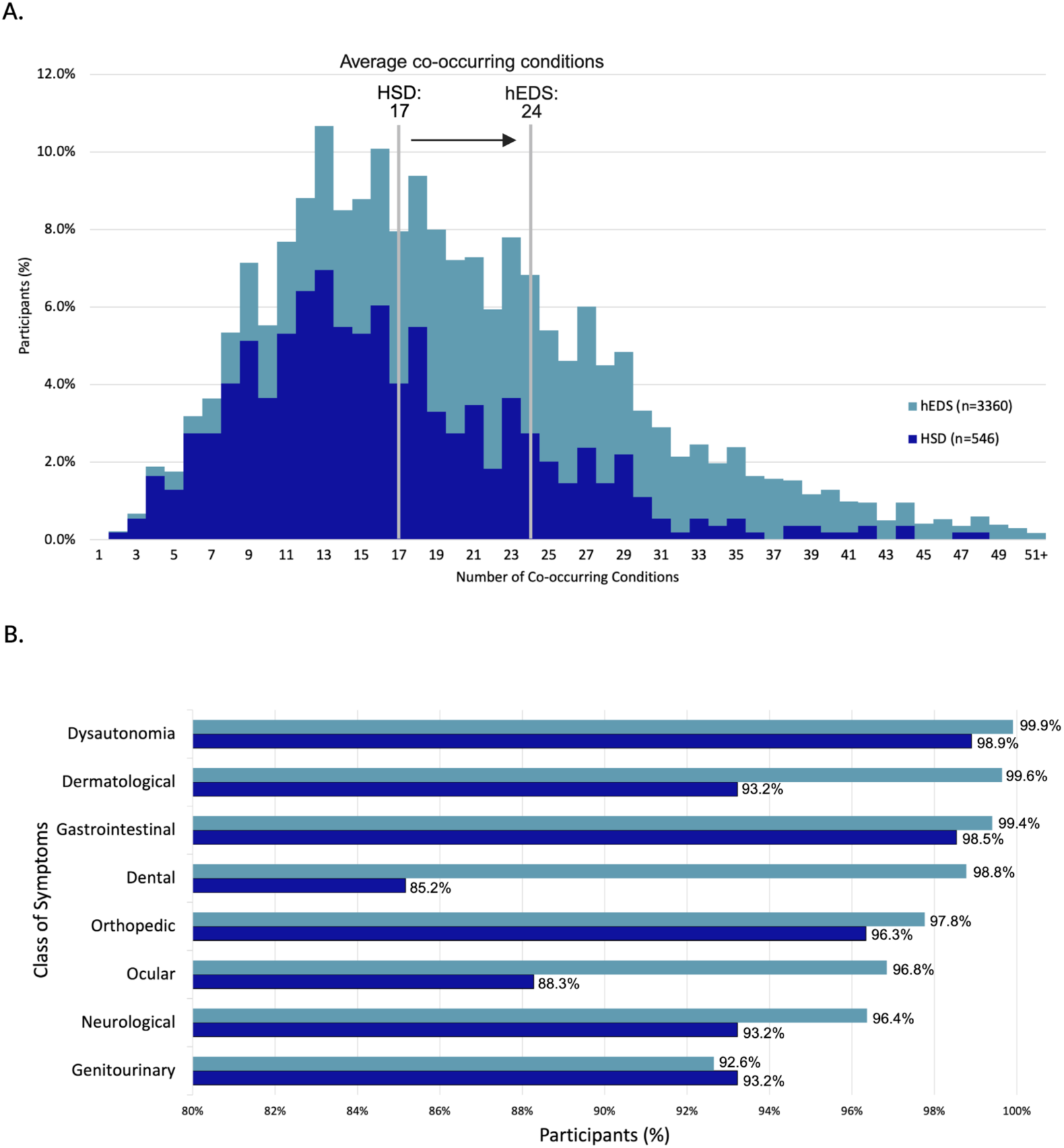
Burden and distribution of co-occurring conditions in survey participants. (A) Distribution of co-occurring conditions per participant, with group averages of 24 (hEDS) and 17 (HSD). (B) Percentage of participants reporting symptoms across eight major organ system categories.

**Table 2.** Most Severe Symptoms Reported by hEDS and HSD Participants.

| **hEDS Most Severe Symptoms** | **hEDS** | **HSD Most Severe Symptoms** | **HSD** |
| --- | --- | --- | --- |
| Chronic pain | 32.6% | Chronic pain | 34.2% |
| Gastrointestinal Symptoms | 13.5% | Joint Manifestations | 14.1% |
| Autonomic dysfunction | 12.8% | Autonomic dysfunction | 10.0% |
| Joint Manifestations | 12.2% | Gastrointestinal Symptoms | 9.6% |
| Neurological Symptoms | 7.2% | Neurological Symptoms | 7.1% |
| Mental health | 4.1% | Mental health | 5.0% |
| Allergic Symptoms | 3.5% | Sleep Issues | 4.0% |
| Sleep Issues | 3.3% | Allergic Symptoms | 3.1% |
| Other | 2.8% | Other | 2.8% |
| Gynecological Symptoms | 1.7% | Gynecological Symptoms | 2.2% |
| Neurodiversity | 0.9% | Neurodiversity | 1.3% |
| Urinary Symptoms | 0.9% | Urinary Symptoms | 1.1% |
| Dental Manifestations | 0.6% | Dental Manifestations | 0.9% |
| Vision Dysfunction | 0.5% | Vision Dysfunction | 0.7% |
| Dermatological Symptoms | 0.4% | Endocrine Dysfunction | 0.7% |
| Endocrine Dysfunction | 0.4% | Dermatological Symptoms | 0.5% |

Participants ranked their three most severe symptom domains from a predefined list. Percentages reflect weighted rankings based on participant responses.
hEDS N=3,360; HSD N=546

To evaluate the impact of hEDS and HSD on daily functioning and productivity, we assessed the time patients spent managing their healthcare. A substantial proportion of individuals, 48.2% of those with hEDS and 33.2% with HSD (p < 0.0001), reported spending five or more hours per week, on average, coordinating and receiving medical care (**Table 3**). This included working with insurance, coordinating with medical providers, and attending appointments, with the majority of participants coordinating this care themselves (92.0% hEDS, 90.5% HSD). Participants indicated that they see multiple medical specialists annually, with hEDS participants averaging a higher number (6.5 hEDS, 5.2 HSD) (*p*<0.0001) than HSD participants in the past year. The most frequently visited specialties included family medicine, cardiology, gastroenterology, neurology/neurosurgery, and emergency medicine.

**Table 3.**
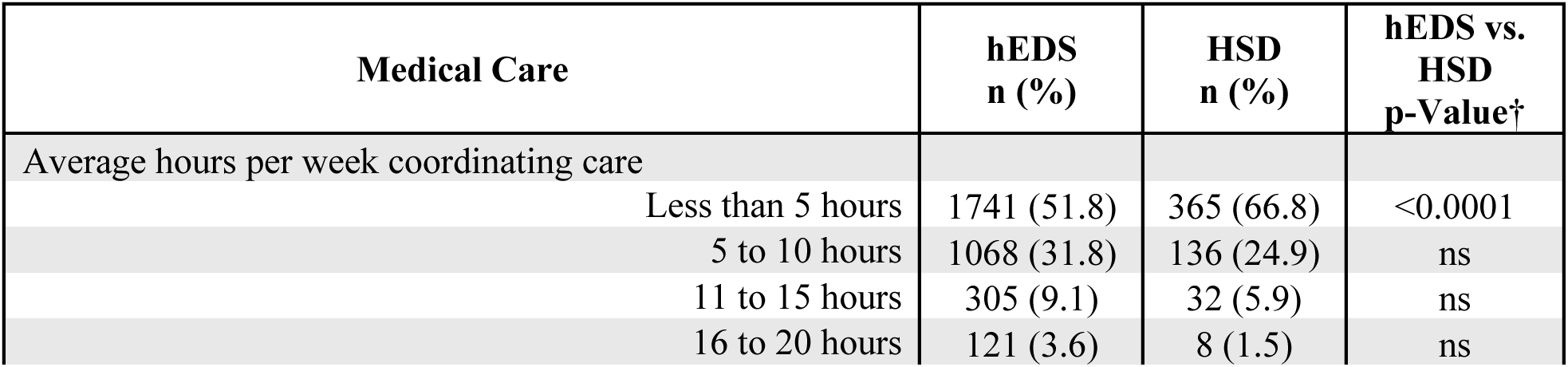
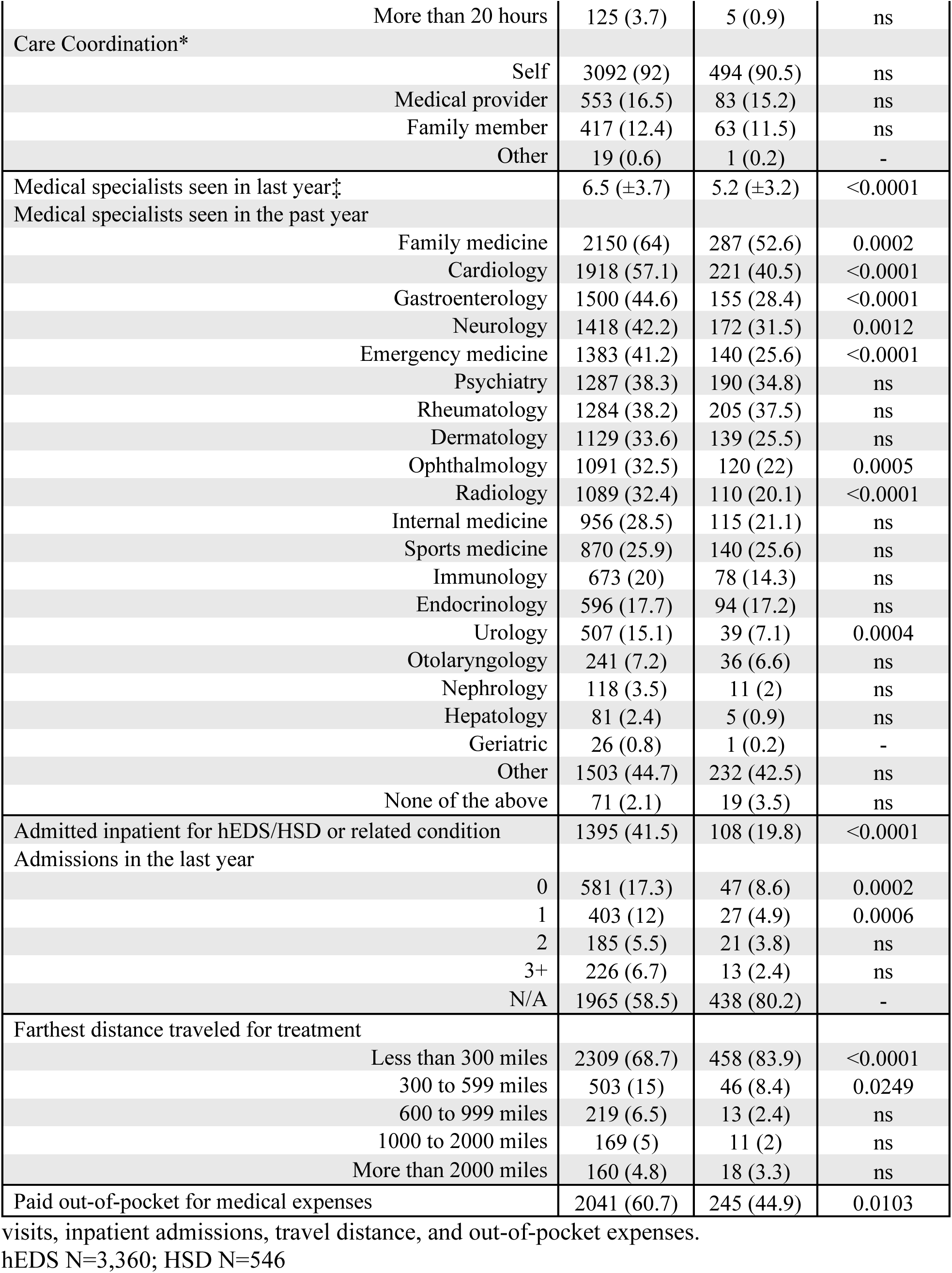
Medical Care Access and Burden in hEDS and HSD Participants.

| **Medical Care** | **hEDS**<br>**n (%)** | **HSD**<br>**n (%)** | **hEDS vs.**<br>**HSD**<br>**p-Value†** |
| --- | --- | --- | --- |
| Average hours per week coordinating care |  |  |  |
| Less than 5 hours | 1741 (51.8) | 365 (66.8) | <0.0001 |
| 5 to 10 hours | 1068 (31.8) | 136 (24.9) | ns |
| 11 to 15 hours | 305 (9.1) | 32 (5.9) | ns |
| 16 to 20 hours | 121 (3.6) | 8 (1.5) | ns |
| More than 20 hours | 125 (3.7) | 5 (0.9) | ns |
| Care Coordination* |  |  |  |
| Self | 3092 (92) | 494 (90.5) | ns |
| Medical provider | 553 (16.5) | 83 (15.2) | ns |
| Family member | 417 (12.4) | 63 (11.5) | ns |
| Other | 19 (0.6) | 1 (0.2) | - |
| Medical specialists seen in last year‡ | 6.5 (±3.7) | 5.2 (±3.2) | <0.0001 |
| Medical specialists seen in the past year |  |  |  |
| Family medicine | 2150 (64) | 287 (52.6) | 0.0002 |
| Cardiology | 1918 (57.1) | 221 (40.5) | <0.0001 |
| Gastroenterology | 1500 (44.6) | 155 (28.4) | <0.0001 |
| Neurology | 1418 (42.2) | 172 (31.5) | 0.0012 |
| Emergency medicine | 1383 (41.2) | 140 (25.6) | <0.0001 |
| Psychiatry | 1287 (38.3) | 190 (34.8) | ns |
| Rheumatology | 1284 (38.2) | 205 (37.5) | ns |
| Dermatology | 1129 (33.6) | 139 (25.5) | ns |
| Ophthalmology | 1091 (32.5) | 120 (22) | 0.0005 |
| Radiology | 1089 (32.4) | 110 (20.1) | <0.0001 |
| Internal medicine | 956 (28.5) | 115 (21.1) | ns |
| Sports medicine | 870 (25.9) | 140 (25.6) | ns |
| Immunology | 673 (20) | 78 (14.3) | ns |
| Endocrinology | 596 (17.7) | 94 (17.2) | ns |
| Urology | 507 (15.1) | 39 (7.1) | 0.0004 |
| Otolaryngology | 241 (7.2) | 36 (6.6) | ns |
| Nephrology | 118 (3.5) | 11 (2) | ns |
| Hepatology | 81 (2.4) | 5 (0.9) | ns |
| Geriatric | 26 (0.8) | 1 (0.2) | - |
| Other | 1503 (44.7) | 232 (42.5) | ns |
| None of the above | 71 (2.1) | 19 (3.5) | ns |
| Admitted inpatient for hEDS/HSD or related condition | 1395 (41.5) | 108 (19.8) | <0.0001 |
| Admissions in the last year |  |  |  |
| 0 | 581 (17.3) | 47 (8.6) | 0.0002 |
| 1 | 403 (12) | 27 (4.9) | 0.0006 |
| 2 | 185 (5.5) | 21 (3.8) | ns |
| 3+ | 226 (6.7) | 13 (2.4) | ns |
| N/A | 1965 (58.5) | 438 (80.2) | - |
| Farthest distance traveled for treatment |  |  |  |
| Less than 300 miles | 2309 (68.7) | 458 (83.9) | <0.0001 |
| 300 to 599 miles | 503 (15) | 46 (8.4) | 0.0249 |
| 600 to 999 miles | 219 (6.5) | 13 (2.4) | ns |
| 1000 to 2000 miles | 169 (5) | 11 (2) | ns |
| More than 2000 miles | 160 (4.8) | 18 (3.3) | ns |
| Paid out-of-pocket for medical expenses | 2041 (60.7) | 245 (44.9) | 0.0103 |

visits, inpatient admissions, travel distance, and out-of-pocket expenses.
hEDS N=3,360; HSD N=546

Inpatient hospitalizations were more common in hEDS participants, with 41.5% of hEDS respondents compared to 19.8% of HSD patients (*p*<0.0001). More hEDS participants reported being admitted to the hospital at least once in the last year compared to HSD participants (24.2% hEDS, 11.2% HSD). When asked the furthest distance participants had traveled to receive medical care pertaining to hEDS/HSD, 31.3% of hEDS and 9.5% of HSD participants report having traveled 300 or more miles, with a subset of participants that have traveled more than 2,000 miles (4.8% hEDS, 3.3% HSD). Regarding the financial burden of these diseases, 78.2% of hEDS and 68.1% HSD participants report paying out-of-pocket for medical care from a specialist not covered by insurance (*p*<0.01).

### Multisystemic Clinical Presentation

#### Pain

Chronic pain is a key feature of symptomatic joint hypermobility, with 98.8% of hEDS participants reporting it, compared with 92.7% of HSD participants (*p*<0.0001) (**Table 4**). The most frequently reported pain sites were the neck (79.9% hEDS, 67.2% HSD), lower back (77.9% hEDS, 65.8% HSD), and shoulder (76.7% hEDS, 65.9% HSD) in both groups (each *p*<0.0001).

**Table 4.**
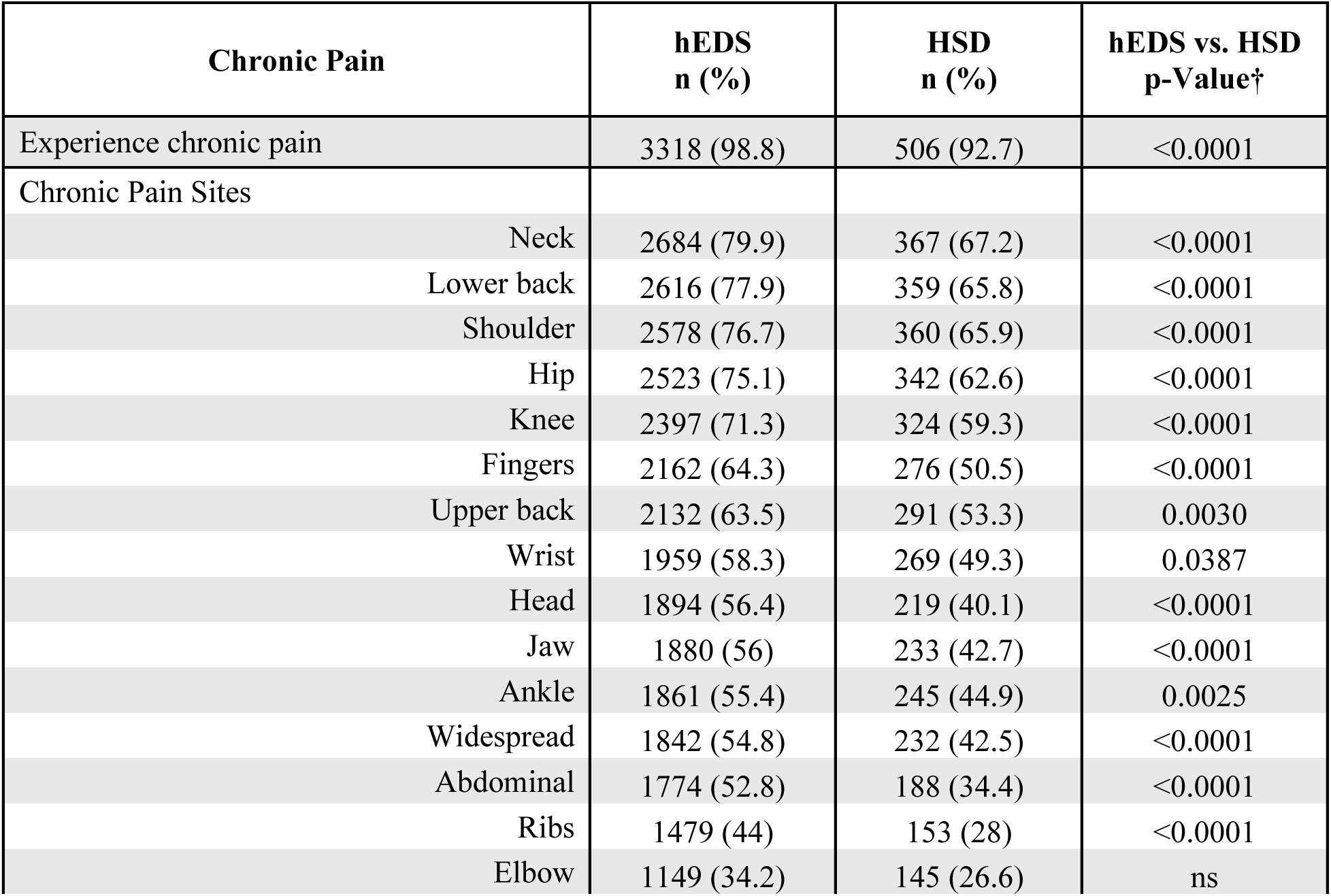
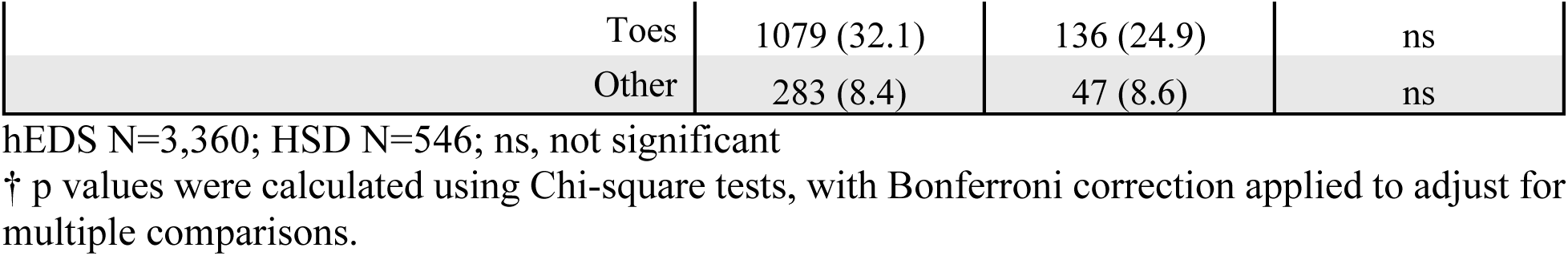
Prevalence of Chronic Pain and Pain Locations in hEDS and HSD.

Given the pain burden in these populations, pain management is a common concern for patients. Responses of participants with hEDS were analyzed regarding which medications they have tried for their pain, and of those, which medications participants considered effective (**Fig 5**). hEDS participants report medical marijuana (66%) and ketamine (55.9%) as the most effective for pain.

**Fig 5.**
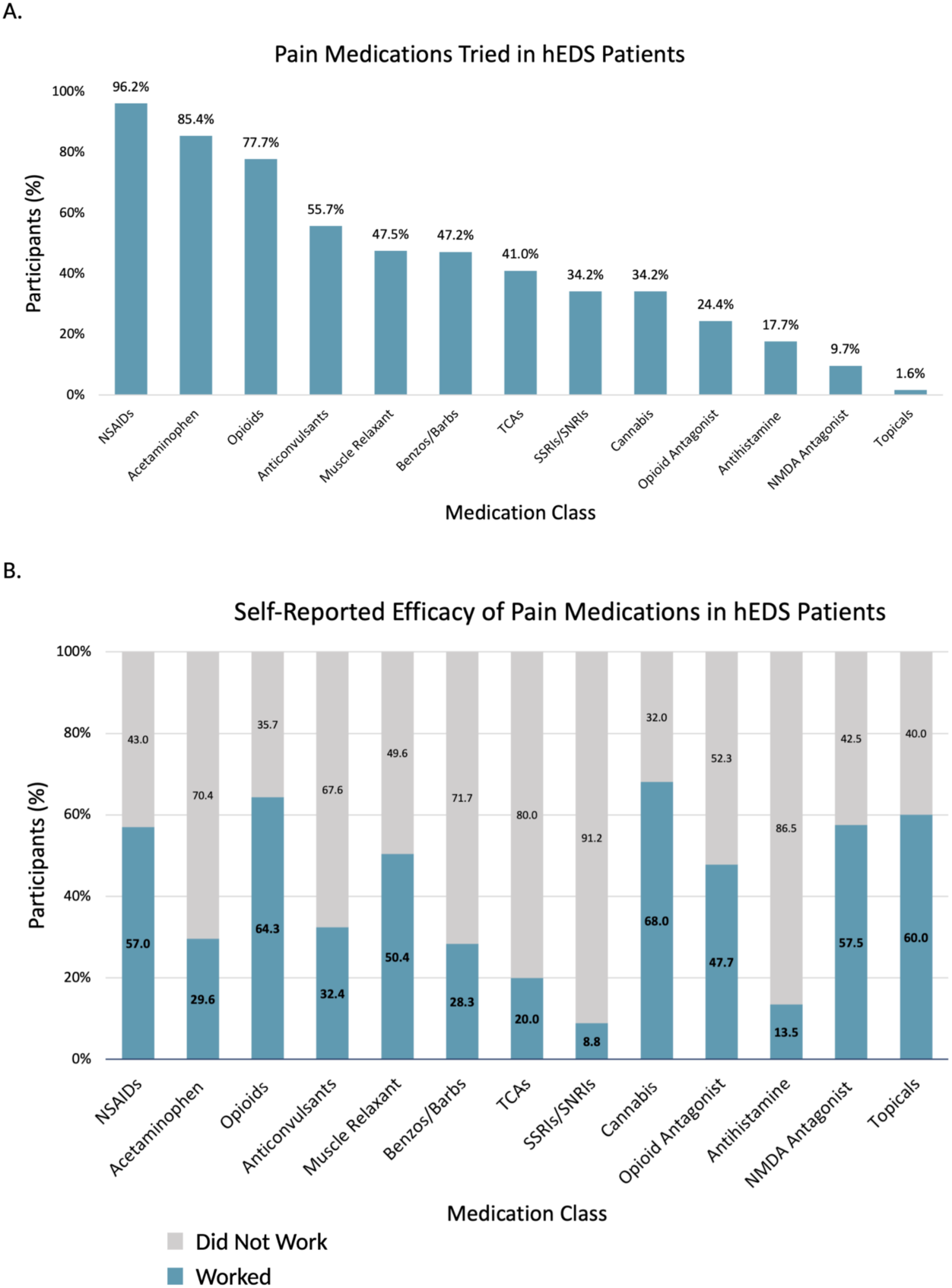
Pain medication use and perceived efficacy in hEDS participants (n = 3,360). (A) Percentage of hEDS participants reporting use of each medication class. (B) Self-reported effectiveness of each medication class, shown as proportion who reported it worked.

Recently, selective serotonin reuptake inhibitors (SSRIs) and serotonin-norepinephrine reuptake inhibitors (SNRIs) have been increasingly used for pain, but only 8.8% of hEDS participants that have tried SS/SNRIs for pain considered them effective. With limited treatment options available, participants report turning to non-traditional practices (85.0% and 81.5%, respectively) for their treatment, including: massage, meditation, and acupuncture (**Table 5**). A significant proportion of participants also experienced complications related to both local and general anesthesia, with 49.4% of hEDS respondents and 28.4% of HSD respondents reporting complications (*p*<0.0001), including shortened anesthetic effect (61.3% hEDS, 40.0% HSD) and insufficient pain control (76.5% hEDS, 66.5% HSD, *p*<0.001) (**Table 6**). Intubation difficulties (5.7% hEDS, 2.7% HSD) and allergic reactions to anesthesia (4.1% hEDS, 1.5%) were also reported in both groups.

**Table 5.** Alternative Medicine Modalities Tried by hEDS and HSD Participants.

| Alternative Medicine | hEDS<br>n (%) | HSD<br>n (%) |
| --- | --- | --- |
| Tried alternative medicine | 2858 (85.1) | 445 (81.5) |
| Alternative medicine type |  |  |
| Massage | 2366 (70.4) | 369 (67.6) |
| Chiropractic | 1846 (54.9) | 250 (45.8) |
| Meditation | 1553 (46.2) | 218 (39.9) |
| Acupuncture | 1491 (44.4) | 232 (42.5) |
| Functional medicine | 745 (22.2) | 99 (18.1) |
| Naturopathy | 632 (18.8) | 77 (14.1) |
| Homeopathy | 596 (17.7) | 76 (13.9) |
| Chinese medicine | 423 (12.6) | 69 (12.6) |
| Hypnosis | 220 (6.5) | 35 (6.4) |
| Ayurveda | 167 (5) | 19 (3.5) |
| Other | 239 (7.1) | 44 (8.1) |

hEDS N=3,360; HSD N=546

**Table 6.** Anesthesia-Related Complications and Reactions in hEDS and HSD.

| Anesthesia Complications | hEDS<br>n (%) | HSD<br>n (%) | hEDS vs. HSD<br>p-Value† |
| --- | --- | --- | --- |
| Anesthesia Complications | 1660 (49.4) | 155 (28.4) | <0.0001 |
| Anesthesia type |  |  |  |
| Local anesthesia only | 1287 (38.3) | 105 (19.2) | ns |
| General anesthesia only | 1191 (35.4) | 95 (17.4) | ns |
| Both general and local anesthesia | 844 (25.1) | 47 (8.6) | 0.0007 |
| Complication Type |  |  |  |
| Shortened effect | 1270 (37.8) | 103 (18.9) | ns |
| Insufficient pain control | 1017 (30.3) | 62 (11.4) | 0.0002 |
| Intubation complication | 193 (5.7) | 15 (2.7) | ns |
| Anaphylaxis or allergic reaction | 137 (4.1) | 8 (1.5) | ns |
| Other | 400 (11.9) | 46 (8.4) | ns |

hEDS N=3,360; HSD N=546; ns, not significant
† p values were calculated using Chi-square tests, with Bonferroni correction applied to adjust for multiple comparisons.

#### Orthopedic

Musculoskeletal complications were highly prevalent in both hEDS and HSD. hEDS participants had significantly higher rates of tendonitis (48.5% hEDS, 39.4% HSD, *p*<0.05), ligament tears (32.4% hEDS, 23.3% HSD, *p*<0.01), and slipping rib syndrome (11.4% hEDS, 4.0% HSD, *p*<0.0001) compared to those with HSD (**Table 7**). Other orthopedic disorders and injuries were also present, including, bursitis (35.7% hEDS, 28.8% HSD), tendon ruptures (16.2% hEDS, 11.5% HSD), and congenital hip dislocation (4.8% hEDS, 2.2% HSD). Compared to the All of Us dataset, hEDS participants showed significantly higher rates (*p*<0.0001) for most orthopedic conditions surveyed, including tendonitis, bursitis, ligament tears, and slipping rib syndrome.

**Table 7.** Prevalence of Orthopedic Diagnoses in hEDS, HSD, and All of Us.

| Orthopedic Disorders | hEDS<br>n (%) | HSD<br>n (%) | hEDS vs.<br>HSD<br>p-Value† | All of Us<br>n (%) | hEDS vs.<br>All of Us<br>p-Value† |
| --- | --- | --- | --- | --- | --- |
| Tendonitis | 1631 (48.5) | 215 (39.4) | 0.0352 | 23000 (6.5) | <0.0001 |
| Bursitis | 1199 (35.7) | 157 (28.8) | ns | 29060 (8.2) | <0.0001 |
| Ligament tear | 1090 (32.4) | 127 (23.3) | 0.0091 | 61560 (17.4) | <0.0001 |
| Tendon rupture | 543 (16.2) | 63 (11.5) | ns | 1320 (0.4) | <0.0001 |
| Slipping rib syndrome | 383 (11.4) | 22 (4) | 0.0001 | 7040 (2) | <0.0001 |
| Congenital hip dislocation | 161 (4.8) | 12 (2.2) | ns | 100 (0) | <0.0001 |
| Bowing of legs | 159 (4.7) | 17 (3.1) | ns | 940 (0.3) | <0.0001 |
| Osgood-Schlatter disease | 155 (4.6) | 19 (3.5) | ns | - | - |
| Pectus excavatum | 125 (3.7) | 10 (1.8) | ns | 340 (0.1) | <0.0001 |
| Joint contracture | 70 (2.1) | 7 (1.3) | ns | 180 (0.1) | <0.0001 |
| Club foot | 70 (2.1) | 5 (0.9) | ns | 40 (0) | <0.0001 |
| Congenital muscle hypotonia | 42 (1.3) | 3 (0.5) | - | - | - |
| None of the above | 885 (26.3) | 205 (37.5) | <0.0001 | - | - |

hEDS N=3,360; HSD N=546; All of Us N=354,400; ns, not significant
† p values were calculated using Chi-square tests, with Bonferroni correction applied to adjust for multiple comparisons.
– indicates the response was excluded from analysis.

Participants with hEDS reported significantly higher prevalence of joint subluxations (96.8% hEDS, 85.5% HSD), joint dislocations (64.3% hEDS, 35.9% HSD), and unexplained fractures (19.8% hEDS, 9.5% HSD) compared to participants with HSD (each *p*<0.0001) (**Table 8**). Correspondingly, a much smaller proportion of hEDS participants reported having none of these joint issues (2.2% hEDS, 11.7% HSD) (each *p*<0.0001). The shoulders (80.5% hEDS, 60.1% HSD), hips (72.3% hEDS, 46.9% HSD), and knees (69.8% hEDS, 50.4% HSD) were among the most frequently affected joints. Additionally, several anatomical abnormalities were reported in the hEDS group, including missing or extra ribs (1.2%), sacralization of the L5 vertebrae (1.2%), and missing or extra vertebrae (0.3%) (**Table 9**).

**Table 8.** Orthopedic Symptoms and Joint Sites Impacted in hEDS and HSD.

| Orthopedic Symptoms | hEDS<br>n (%) | HSD<br>n (%) | hEDS vs. HSD<br>p-Value† |
| --- | --- | --- | --- |
| Joint subluxations | 3254 (96.8) | 467 (85.5) | <0.0001 |
| Joint dislocations | 2159 (64.3) | 196 (35.9) | <0.0001 |
| Unexplained fractures | 666 (19.8) | 52 (9.5) | <0.0001 |
| None of the above | 75 (2.2) | 64 (11.7) | <0.0001 |
| Joints Affected |  |  |  |
| Shoulder | 2704 (80.5) | 328 (60.1) | <0.0001 |
| Hip | 2428 (72.3) | 256 (46.9) | <0.0001 |
| Knee | 2345 (69.8) | 275 (50.4) | <0.0001 |
| Fingers | 2313 (68.8) | 258 (47.3) | <0.0001 |
| Ribs | 2022 (60.2) | 198 (36.3) | <0.0001 |
| Ankle | 2003 (59.6) | 221 (40.5) | <0.0001 |
| Wrist | 1872 (55.7) | 180 (33) | <0.0001 |
| Toes | 1387 (41.3) | 124 (22.7) | <0.0001 |
| Elbow | 1315 (39.1) | 139 (25.5) | <0.0001 |
| Spine | 1288 (38.3) | 129 (23.6) | <0.0001 |
| Other | 454 (13.5) | 47 (8.6) | ns |

hEDS N=3,360; HSD N=546; ns, not significant
† p values were calculated using Chi-square tests, with Bonferroni correction applied to adjust for multiple comparisons.

**Table 9.** Reported Anatomical Abnormalities in hEDS Participants.

| Anatomical Abnormalities | hEDS<br>N=3,360<br>n (%) |
| --- | --- |
| Uterus | 549 (16.3) |
| Ribs | 40 (1.2) |
| Lumbar/sacral | 38 (1.1) |
| Extra or missing vertebrae | 9 (0.3) |

#### Neurological

Migraine was the most frequently reported neurological diagnosis among both groups, while affecting significantly more hEDS participants than HSD participants (64.3% hEDS, 46.7% HSD, *p*<0.0001) (**Table 10**). hEDS participants also reported significantly higher rates of scoliosis, Raynaud’s phenomenon, upper cervical instability, and occipital neuralgia, when compared to both HSD and All of Us datasets (each *p*<0.0001). Other neurological abnormalities were more common in hEDS than HSD, particularly tinnitus (31.1% hEDS, 22.2% HSD, *p*<0.05), herniated disc(s) (26.3% hEDS, 17.8% HSD, *p*<0.05), spinal stenosis (13.2% hEDS, 6.6% HSD, *p*<0.01), small fiber neuropathy (10.0% hEDS, 4.4% HSD, *p*<0.05), Chiari malformation (6.3% hEDS, 0.9% HSD, *p*<0.001), kyphoscoliosis (5.9% hEDS, 1.6% HSD, *p*<0.05), and tethered cord syndrome (4.6% hEDS, 0.9% HSD, *p*<0.05). There was no significant difference in the prevalence of voice disorders between the three groups (4.0% hEDS, 1.1% HSD, 3.2% All of Us), while hearing impairments were reported at a significantly lower rate in the hEDS cohort compared to the general population (*p*<0.0001). Neurological symptoms were highly prevalent among survey respondents, with significant differences observed between the hEDS and HSD groups. Weakness was the most reported symptom (79.6% hEDS, 71.6% HSD) (*p*<0.05), followed closely by muscle spasms (78.3% hEDS, 67.2% HSD) (*p*<0.0001), tingling (76.0% hEDS, 67.0% HSD) (*p*<0.01), and numbness (70.4% hEDS, 52.7% HSD) (*p*<0.0001) (**Table 11**).

**Table 10.**
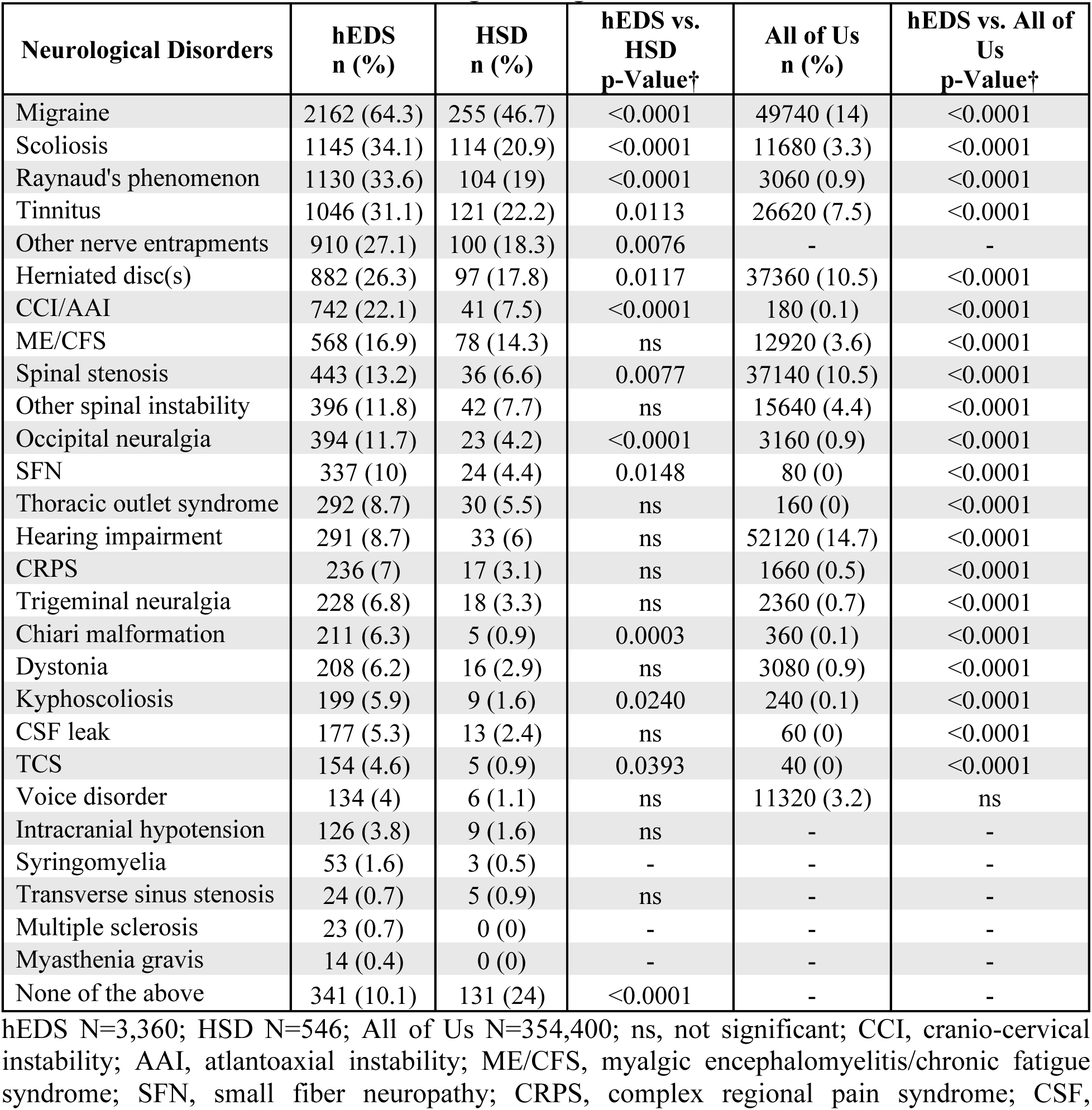
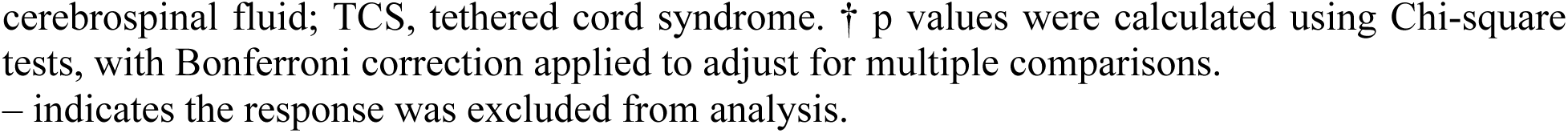
Prevalence of Neurological Diagnoses in hEDS, HSD, and All of Us.

| Neurological Disorders | hEDS<br>n (%) | HSD<br>n (%) | hEDS vs.<br>HSD<br>p-Value† | All of Us<br>n (%) | hEDS vs. All of<br>Us<br>p-Value† |
| --- | --- | --- | --- | --- | --- |
| Migraine | 2162 (64.3) | 255 (46.7) | <0.0001 | 49740 (14) | <0.0001 |
| Scoliosis | 1145 (34.1) | 114 (20.9) | <0.0001 | 11680 (3.3) | <0.0001 |
| Raynaud's phenomenon | 1130 (33.6) | 104 (19) | <0.0001 | 3060 (0.9) | <0.0001 |
| Tinnitus | 1046 (31.1) | 121 (22.2) | 0.0113 | 26620 (7.5) | <0.0001 |
| Other nerve entrapments | 910 (27.1) | 100 (18.3) | 0.0076 | - | - |
| Herniated disc(s) | 882 (26.3) | 97 (17.8) | 0.0117 | 37360 (10.5) | <0.0001 |
| CCI/AAI | 742 (22.1) | 41 (7.5) | <0.0001 | 180 (0.1) | <0.0001 |
| ME/CFS | 568 (16.9) | 78 (14.3) | ns | 12920 (3.6) | <0.0001 |
| Spinal stenosis | 443 (13.2) | 36 (6.6) | 0.0077 | 37140 (10.5) | <0.0001 |
| Other spinal instability | 396 (11.8) | 42 (7.7) | ns | 15640 (4.4) | <0.0001 |
| Occipital neuralgia | 394 (11.7) | 23 (4.2) | <0.0001 | 3160 (0.9) | <0.0001 |
| SFN | 337 (10) | 24 (4.4) | 0.0148 | 80 (0) | <0.0001 |
| Thoracic outlet syndrome | 292 (8.7) | 30 (5.5) | ns | 160 (0) | <0.0001 |
| Hearing impairment | 291 (8.7) | 33 (6) | ns | 52120 (14.7) | <0.0001 |
| CRPS | 236 (7) | 17 (3.1) | ns | 1660 (0.5) | <0.0001 |
| Trigeminal neuralgia | 228 (6.8) | 18 (3.3) | ns | 2360 (0.7) | <0.0001 |
| Chiari malformation | 211 (6.3) | 5 (0.9) | 0.0003 | 360 (0.1) | <0.0001 |
| Dystonia | 208 (6.2) | 16 (2.9) | ns | 3080 (0.9) | <0.0001 |
| Kyphoscoliosis | 199 (5.9) | 9 (1.6) | 0.0240 | 240 (0.1) | <0.0001 |
| CSF leak | 177 (5.3) | 13 (2.4) | ns | 60 (0) | <0.0001 |
| TCS | 154 (4.6) | 5 (0.9) | 0.0393 | 40 (0) | <0.0001 |
| Voice disorder | 134 (4) | 6 (1.1) | ns | 11320 (3.2) | ns |
| Intracranial hypotension | 126 (3.8) | 9 (1.6) | ns | - | - |
| Syringomyelia | 53 (1.6) | 3 (0.5) | - | - | - |
| Transverse sinus stenosis | 24 (0.7) | 5 (0.9) | ns | - | - |
| Multiple sclerosis | 23 (0.7) | 0 (0) | - | - | - |
| Myasthenia gravis | 14 (0.4) | 0 (0) | - | - | - |
| None of the above | 341 (10.1) | 131 (24) | <0.0001 | - | - |

hEDS N=3,360; HSD N=546; All of Us N=354,400; ns, not significant; CCI, cranio-cervical instability; AAI, atlantoaxial instability; ME/CFS, myalgic encephalomyelitis/chronic fatigue syndrome; SFN, small fiber neuropathy; CRPS, complex regional pain syndrome; CSF,
cerebrospinal fluid; TCS, tethered cord syndrome. † p values were calculated using Chi-square tests, with Bonferroni correction applied to adjust for multiple comparisons. – indicates the response was excluded from analysis.

**Table 11.** Prevalence of Neurological Symptoms in hEDS and HSD.

| Neurological Symptoms | hEDS<br>n (%) | HSD<br>n (%) | hEDS vs. HSD<br>p-Value† |
| --- | --- | --- | --- |
| Weakness | 2676 (79.6) | 391 (71.6) | 0.0121 |
| Muscle spasm | 2632 (78.3) | 367 (67.2) | <0.0001 |
| Tingling | 2555 (76) | 366 (67) | 0.0037 |
| Numbness | 2365 (70.4) | 288 (52.7) | <0.0001 |
| None of the above | 122 (3.6) | 37 (6.8) | ns |

hEDS N=3,360; HSD N=546; ns, not significant
† p values were calculated using Chi-square tests, with Bonferroni correction applied to adjust for multiple comparisons.

#### Autonomic

Autonomic dysfunction was commonly reported among participants, with 71.4% of hEDS and 40.3% of HSD participants reporting at least one diagnosed autonomic disorder (*p*<0.0001), with a higher prevalence of each type in the hEDS group (**Table 12**). Postural orthostatic tachycardia syndrome (POTS) was the most frequently diagnosed autonomic disorder, affecting hEDS participants at a higher rate (52.7%) than those with HSD (26.0%) (*p*<0.0001). General dysautonomia—clinically coded as “disorder of the autonomic nervous system, unspecified”— was also significantly more prevalent in hEDS than HSD (27.0% hEDS, 14.7% HSD, *p*<0.0001), along with orthostatic hypotension (16.2% hEDS, 8.1% HSD, *p*<0.001) and orthostatic intolerance (15.2%, hEDS, 7.9% HSD, *p*<0.01). Symptoms of dysautonomia were highly prevalent, with fatigue (98.6% hEDS, 94.5% HSD), dizziness (95.7%, 88.3%), and brain fog (95.6%, 89.9%) being the most highly reported (each *p*<0.0001) (**Table 13**). Heart palpitations (94.6% hEDS, 85.7% HSD) and thermoregulatory dysfunction (92.0%, 80.6%) were also frequent, suggesting significant autonomic involvement in both groups (each *p*<0.0001). Despite these high symptom burdens, 28.6% of hEDS and 59.7% of HSD participants reported having no formal autonomic diagnosis.

**Table 12.**
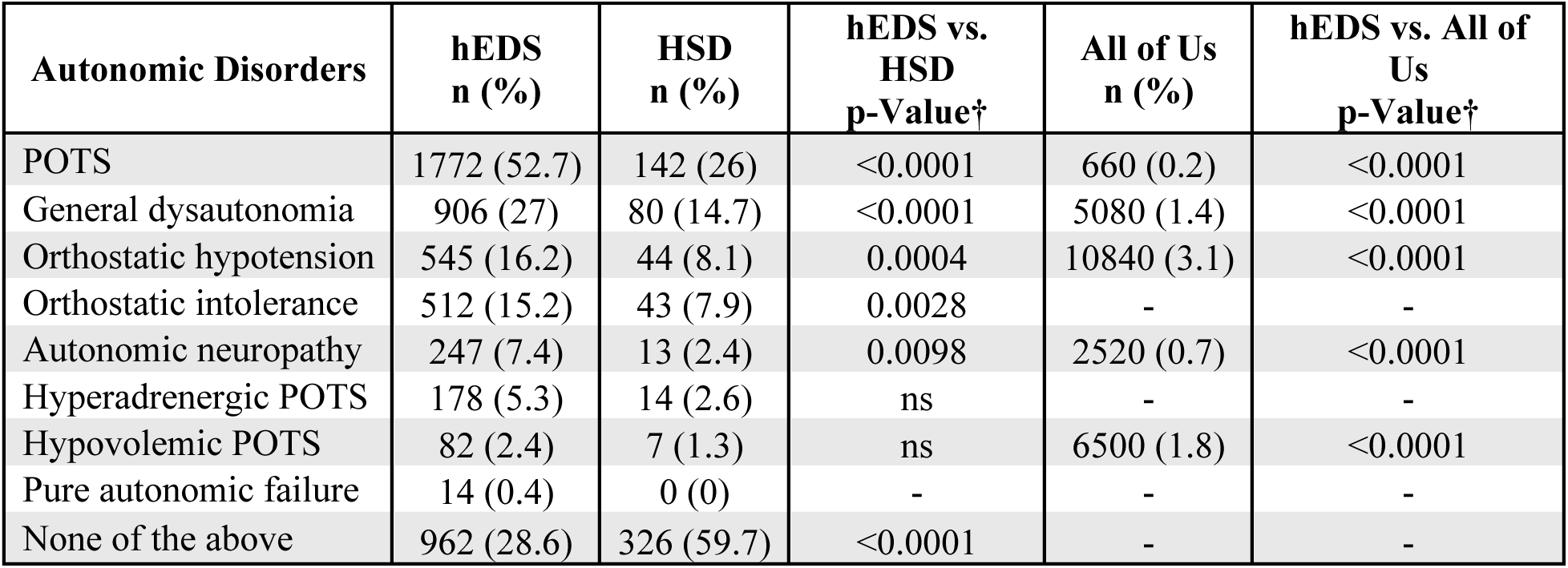
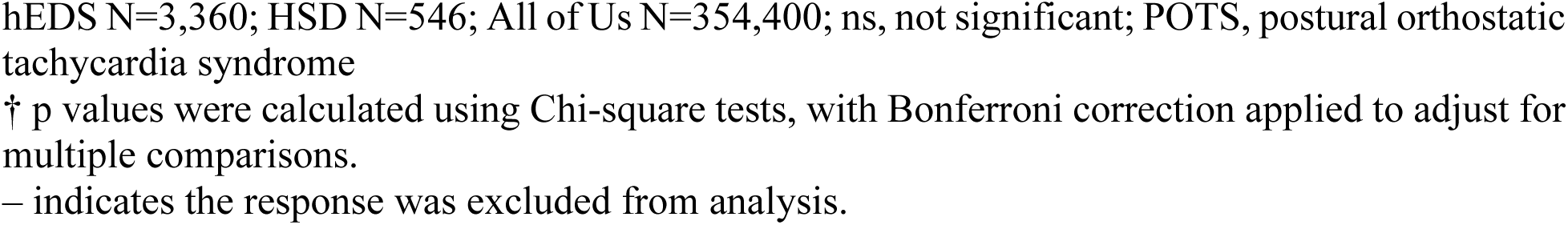
Prevalence of Autonomic Disorders in hEDS, HSD, and All of Us.

| Autonomic Disorders | hEDS<br>n (%) | HSD<br>n (%) | hEDS vs.<br>HSD<br>p-Value† | All of Us<br>n (%) | hEDS vs. All of<br>Us<br>p-Value† |
| --- | --- | --- | --- | --- | --- |
| POTS | 1772 (52.7) | 142 (26) | <0.0001 | 660 (0.2) | <0.0001 |
| General dysautonomia | 906 (27) | 80 (14.7) | <0.0001 | 5080 (1.4) | <0.0001 |
| Orthostatic hypotension | 545 (16.2) | 44 (8.1) | 0.0004 | 10840 (3.1) | <0.0001 |
| Orthostatic intolerance | 512 (15.2) | 43 (7.9) | 0.0028 | - | - |
| Autonomic neuropathy | 247 (7.4) | 13 (2.4) | 0.0098 | 2520 (0.7) | <0.0001 |
| Hyperadrenergic POTS | 178 (5.3) | 14 (2.6) | ns | - | - |
| Hypovolemic POTS | 82 (2.4) | 7 (1.3) | ns | 6500 (1.8) | <0.0001 |
| Pure autonomic failure | 14 (0.4) | 0 (0) | - | - | - |
| None of the above | 962 (28.6) | 326 (59.7) | <0.0001 | - | - |

hEDS N=3,360; HSD N=546; All of Us N=354,400; ns, not significant; POTS, postural orthostatic tachycardia syndrome
† p values were calculated using Chi-square tests, with Bonferroni correction applied to adjust for multiple comparisons.
– indicates the response was excluded from analysis.

**Table 13.** Prevalence of Autonomic Symptoms in hEDS and HSD.

| Autonomic Symptoms | hEDS<br>n (%) | HSD<br>n (%) | hEDS vs. HSD<br>p-Value† |
| --- | --- | --- | --- |
| Fatigue | 3312 (98.6) | 516 (94.5) | <0.0001 |
| Dizziness | 3216 (95.7) | 482 (88.3) | <0.0001 |
| Brain fog | 3211 (95.6) | 491 (89.9) | <0.0001 |
| Heart palpitations | 3177 (94.6) | 468 (85.7) | <0.0001 |
| Thermoregulatory dysfunction | 3090 (92) | 440 (80.6) | <0.0001 |
| Vertigo | 2782 (82.8) | 371 (67.9) | <0.0001 |
| Trouble swallowing | 2171 (64.6) | 251 (46) | <0.0001 |
| Fainting | 2166 (64.5) | 239 (43.8) | <0.0001 |
| None of the above | 3 (0.1) | 6 (1.1) | – |

hEDS N=3,360; HSD N=546
† p values were calculated using Chi-square tests, with Bonferroni correction applied to adjust for multiple comparisons.
– indicates the response was excluded from analysis.

#### Gastrointestinal

Gastrointestinal (GI) symptoms were common in both groups, with a higher prevalence of at least one GI disorder in the hEDS group compared to HSD (84.3% hEDS, 69.0% HSD) (**Table 14**). Irritable bowel syndrome (IBS) was the most reported GI diagnosis (48.4% hEDS, 36.8% HSD, *p*<0.001). Among IBS subtypes, the mixed variant was most prevalent (27.1% hEDS, 16.3% HSD, *p*<0.0001). Gastroesophageal reflux disease (GERD) was diagnosed in nearly half of the hEDS group (47.6%) compared to 30.8% of HSD participants (*p*<0.0001). Dysmotility was also more frequently diagnosed in hEDS than HSD (26.2% hEDS, 10.4% HSD, *p*<0.0001), with the gastric type (“gastroparesis”) reported the most frequently in both groups (21.2% hEDS, 7.9% HSD, *p*<0.0001). hEDS participants reported a significantly higher prevalence of 13 of 19 GI disorders that were compared to the All of Us dataset (each *p*<0.0001), while obesity (21.7% hEDS, 31.5% All of Us), abdominal hernia (10.1% hEDS, 13.7% All of Us), cholelithiasis (3.5% hEDS, 6.5% All of Us), and liver disease (3.1% hEDS, 17.1% All of Us) were all significantly higher in the general population than hEDS (each *p*<0.0001). Notable, colorectal polyps and ulcerative colitis were not significantly different between hEDS and All of Us groups.

**Table 14.**
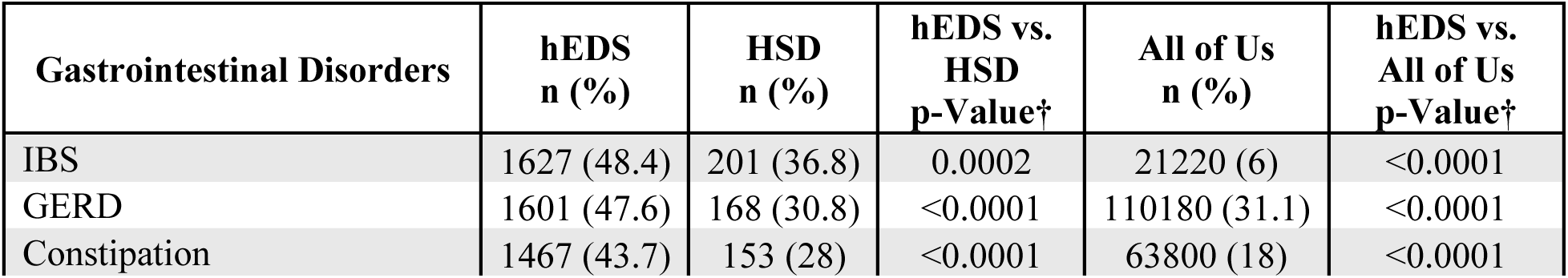
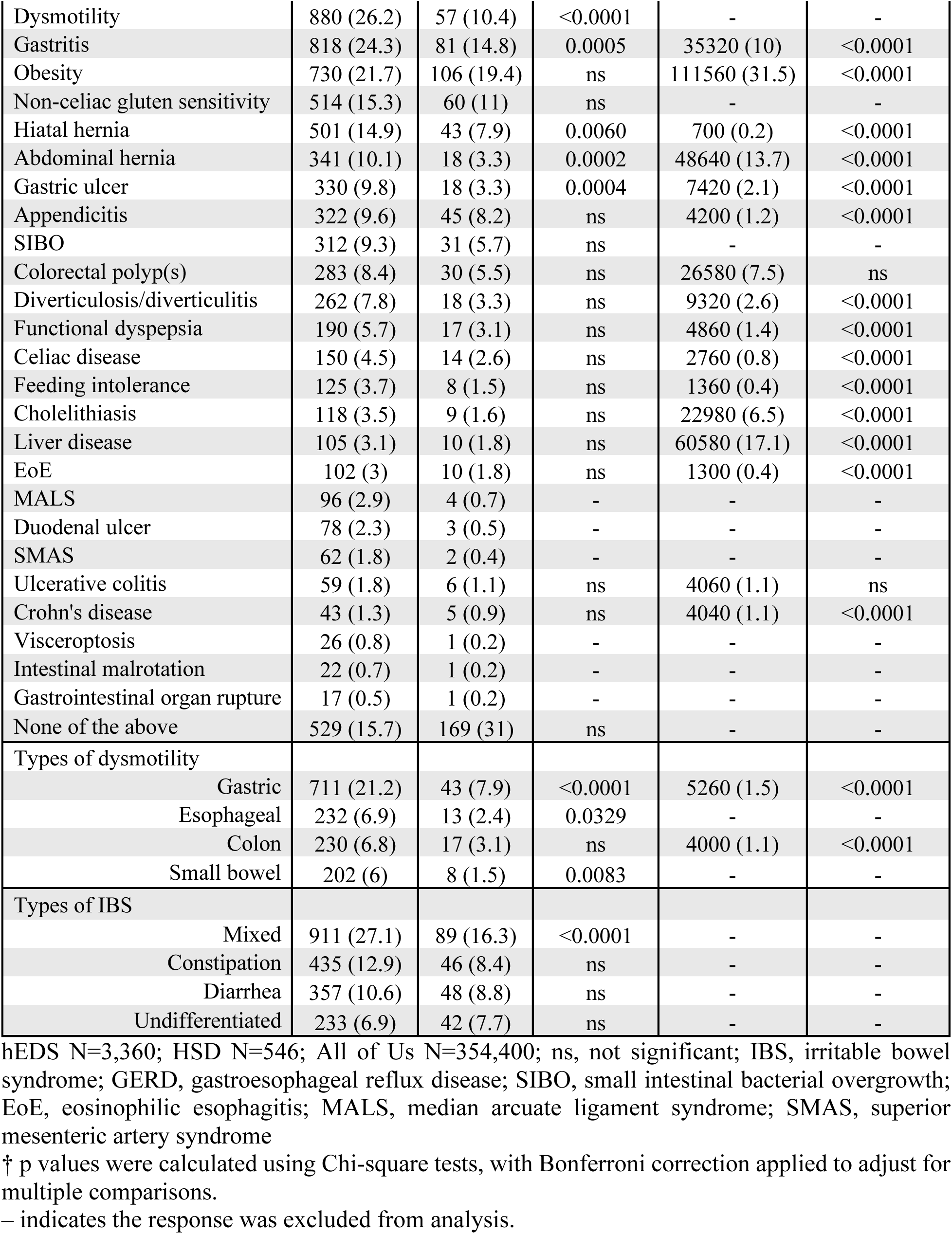
Prevalence of Gastrointestinal Diagnoses in hEDS, HSD, and All of Us.

The most frequently reported symptoms were abdominal pain (92.9% hEDS, 80.4% HSD), nausea (90.6%, 77.8%), bloating (89.6%, 80.4%), and constipation (87.1%, 75.5%) (each *p*<0.0001) (**Table 15**). Severe GI dysfunction requiring feeding devices was significantly more frequent in hEDS (7.1%) than in HSD (2.0%) (*p*<0.01), with nasogastric (3.9%), nasojejunal (3.3%), and total parenteral nutrition (2.2%) being the most frequently utilized options by hEDS participants (**Table 16**). Similarly, colostomies and ileostomies, while rare overall (1.5% hEDS, 1.8% HSD and 1.1% hEDS, 0.4% HSD, respectively), were present in both groups.

**Table 15.** Prevalence of Gastrointestinal Symptoms in hEDS and HSD.

| Gastrointestinal Symptoms | hEDS<br>n (%) | HSD<br>n (%) | hEDS vs. HSD<br>p-Value† |
| --- | --- | --- | --- |
| Abdominal pain | 3120 (92.9) | 439 (80.4) | <0.0001 |
| Nausea | 3044 (90.6) | 425 (77.8) | <0.0001 |
| Bloating | 3012 (89.6) | 439 (80.4) | <0.0001 |
| Constipation | 2925 (87.1) | 412 (75.5) | <0.0001 |
| Diarrhea | 2778 (82.7) | 391 (71.6) | <0.0001 |
| Reflux | 2640 (78.6) | 350 (64.1) | <0.0001 |
| Heartburn | 2488 (74) | 337 (61.7) | <0.0001 |
| Vomiting | 1968 (58.6) | 211 (38.6) | <0.0001 |
| Rectal bleeding | 1523 (45.3) | 166 (30.4) | <0.0001 |
| None of the above | 20 (0.6) | 8 (1.5) | ns |

hEDS N=3,360; HSD N=546; ns, not significant
† p values were calculated using Chi-square tests, with Bonferroni correction applied to adjust for multiple comparisons.

**Table 16.**
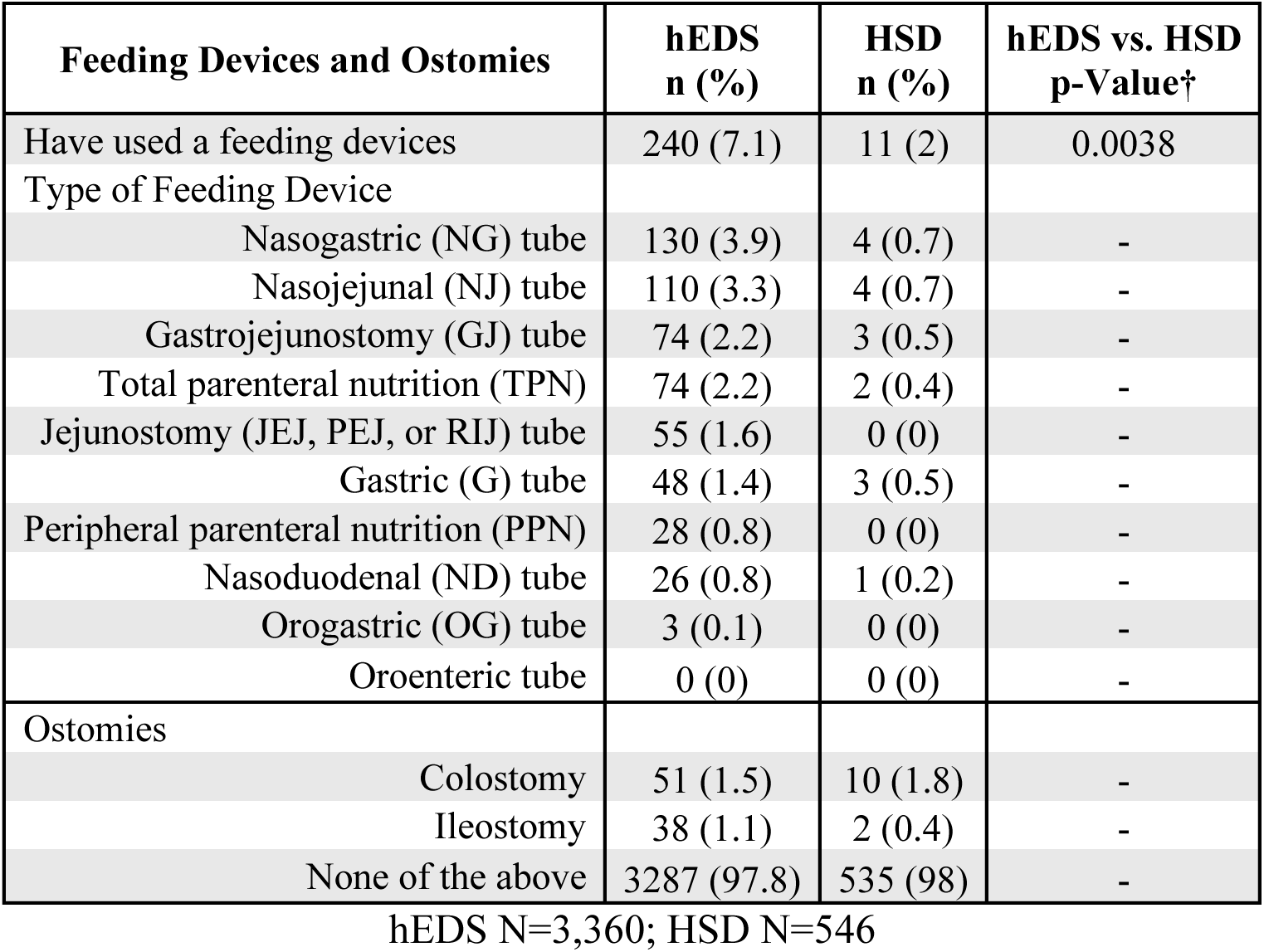
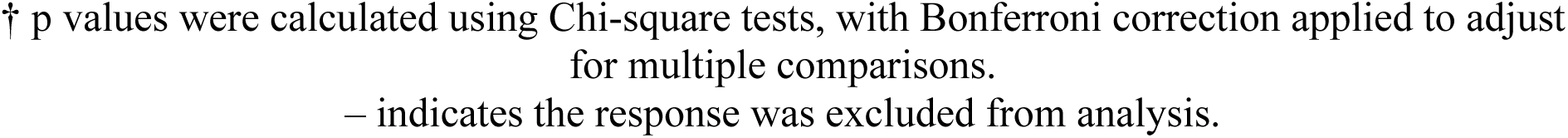
Use of Feeding Devices and Ostomies in hEDS and HSD.

| Feeding Devices and Ostomies | hEDS<br>n (%) | HSD<br>n (%) | hEDS vs. HSD<br>p-Value† |
| --- | --- | --- | --- |
| Have used a feeding devices | 240 (7.1) | 11 (2) | 0.0038 |
| Type of Feeding Device |  |  |  |
| Nasogastric (NG) tube | 130 (3.9) | 4 (0.7) | - |
| Nasojejunal (NJ) tube | 110 (3.3) | 4 (0.7) | - |
| Gastrojejunostomy (GJ) tube | 74 (2.2) | 3 (0.5) | - |
| Total parenteral nutrition (TPN) | 74 (2.2) | 2 (0.4) | - |
| Jejunostomy (JEJ, PEJ, or RIJ) tube | 55 (1.6) | 0 (0) | - |
| Gastric (G) tube | 48 (1.4) | 3 (0.5) | - |
| Peripheral parenteral nutrition (PPN) | 28 (0.8) | 0 (0) | - |
| Nasoduodenal (ND) tube | 26 (0.8) | 1 (0.2) | - |
| Orogastric (OG) tube | 3 (0.1) | 0 (0) | - |
| Oroenteric tube | 0 (0) | 0 (0) | - |
| Ostomies |  |  |  |
| Colostomy | 51 (1.5) | 10 (1.8) | - |
| Ileostomy | 38 (1.1) | 2 (0.4) | - |
| None of the above | 3287 (97.8) | 535 (98) | - |

hEDS N=3,360; HSD N=546
† p values were calculated using Chi-square tests, with Bonferroni correction applied to adjust for multiple comparisons.
– indicates the response was excluded from analysis.

#### Cardiopulmonary

The majority of respondents in both groups reported no diagnosed cardiovascular conditions (62.0% hEDS, 78.9% HSD, *p*<0.0001) (**Table 17**). Of those diagnosed with a cardiac condition, mitral valve defect was the most frequently reported in hEDS (16.3% hEDS, 5.5% HSD, *p*<0.0001), while the most frequently reported issue reported by HSD participants were arrhythmias (other than supraventricular tachycardia and atrial fibrillation) (14.4% hEDS, 9.7% HSD). Other structural heart abnormalities, including tricuspid valve defects (6.0% hEDS, 2.4% HSD) and aortic valve defects (3.3% hEDS, 0.5% HSD), were present, but less frequent in both groups. Less common but notable conditions included atrial fibrillation (2.9% hEDS, 1.3% HSD), stoke/transient ischemic attack (2.4% hEDS, 0.9% HSD), and aortic aneurysm (1.0% hEDS, 0.0% HSD). Compared to the All of Us dataset, hEDS had significantly higher prevalences of mitral valve defects (*p*<0.0001), stroke (*p*<0.0001), and tricuspid valve defects (*p*<0.001). Contrastingly, lung disease and atrial fibrillation had a significantly lower prevalence in hEDS when compared to All of Us data. Many of these cardiopulmonary diseases showed an age-effect of diagnoses, with peak diagnoses in the 26–44-year-old age range (**Supplementary Table 7**).

**Table 17.**
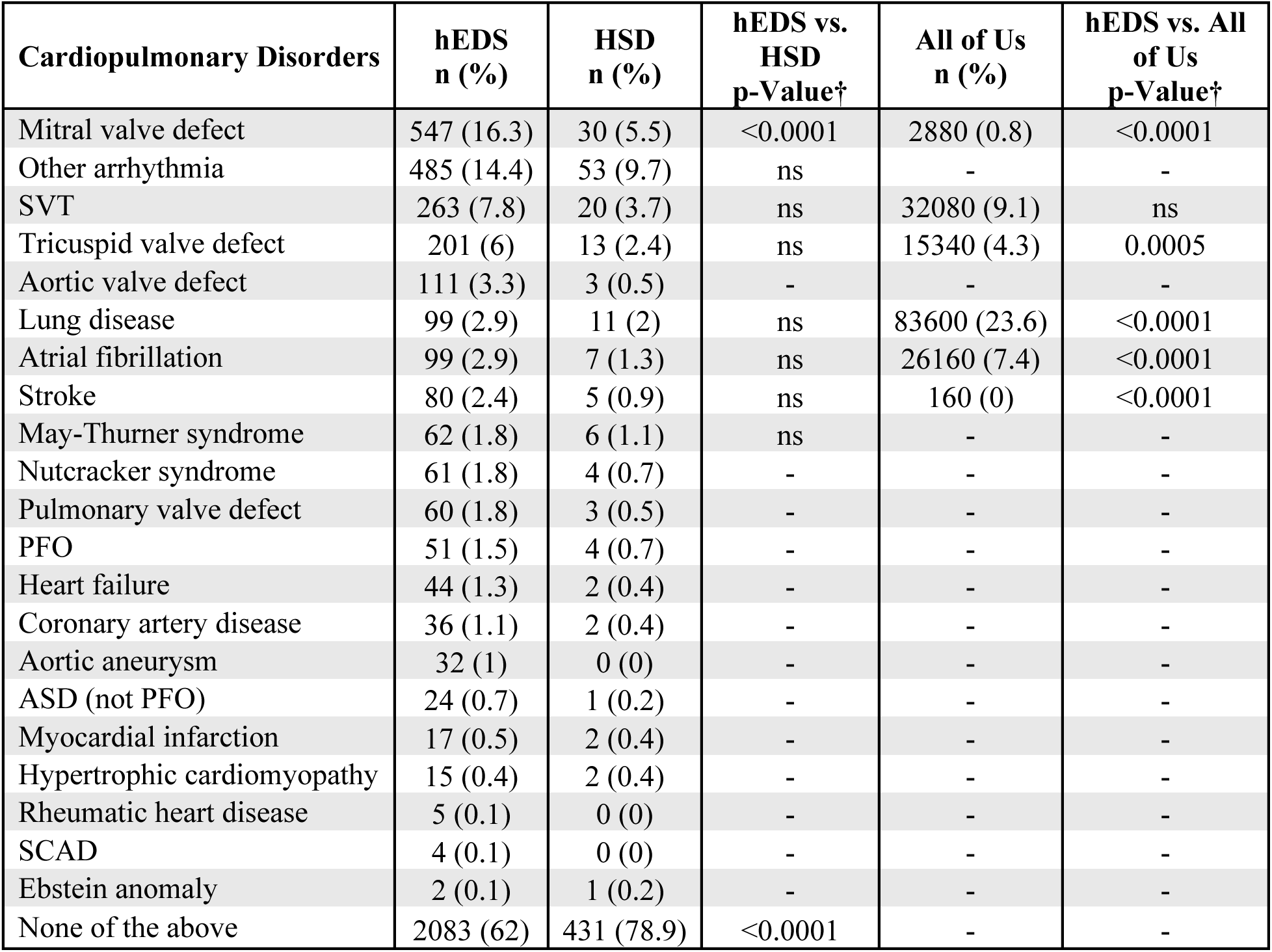
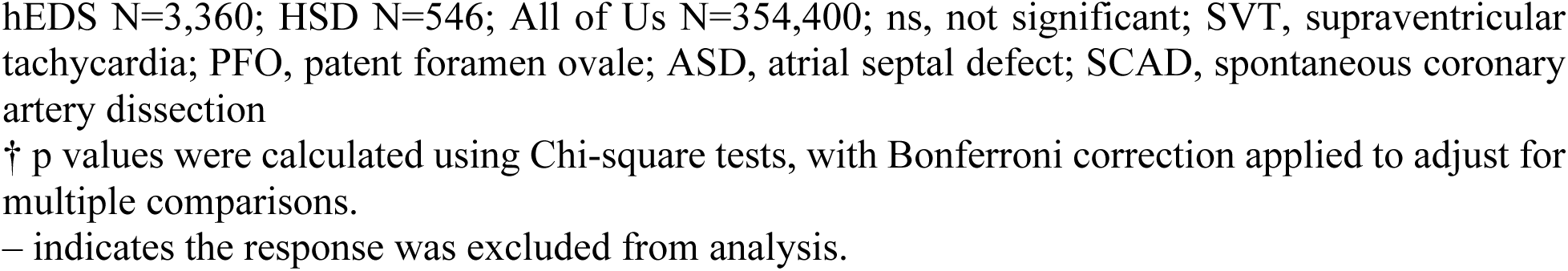
Prevalence of Cardiopulmonary Diagnoses in hEDS, HSD, and All of Us.

| Cardiopulmonary Disorders | hEDS<br>n (%) | HSD<br>n (%) | hEDS vs.<br>HSD<br>p-Value† | All of Us<br>n (%) | hEDS vs. All<br>of Us<br>p-Value† |
| --- | --- | --- | --- | --- | --- |
| Mitral valve defect | 547 (16.3) | 30 (5.5) | <0.0001 | 2880 (0.8) | <0.0001 |
| Other arrhythmia | 485 (14.4) | 53 (9.7) | ns | - | - |
| SVT | 263 (7.8) | 20 (3.7) | ns | 32080 (9.1) | ns |
| Tricuspid valve defect | 201 (6) | 13 (2.4) | ns | 15340 (4.3) | 0.0005 |
| Aortic valve defect | 111 (3.3) | 3 (0.5) | - | - | - |
| Lung disease | 99 (2.9) | 11 (2) | ns | 83600 (23.6) | <0.0001 |
| Atrial fibrillation | 99 (2.9) | 7 (1.3) | ns | 26160 (7.4) | <0.0001 |
| Stroke | 80 (2.4) | 5 (0.9) | ns | 160 (0) | <0.0001 |
| May-Thurner syndrome | 62 (1.8) | 6 (1.1) | ns | - | - |
| Nutcracker syndrome | 61 (1.8) | 4 (0.7) | - | - | - |
| Pulmonary valve defect | 60 (1.8) | 3 (0.5) | - | - | - |
| PFO | 51 (1.5) | 4 (0.7) | - | - | - |
| Heart failure | 44 (1.3) | 2 (0.4) | - | - | - |
| Coronary artery disease | 36 (1.1) | 2 (0.4) | - | - | - |
| Aortic aneurysm | 32 (1) | 0 (0) | - | - | - |
| ASD (not PFO) | 24 (0.7) | 1 (0.2) | - | - | - |
| Myocardial infarction | 17 (0.5) | 2 (0.4) | - | - | - |
| Hypertrophic cardiomyopathy | 15 (0.4) | 2 (0.4) | - | - | - |
| Rheumatic heart disease | 5 (0.1) | 0 (0) | - | - | - |
| SCAD | 4 (0.1) | 0 (0) | - | - | - |
| Ebstein anomaly | 2 (0.1) | 1 (0.2) | - | - | - |
| None of the above | 2083 (62) | 431 (78.9) | <0.0001 | - | - |

hEDS N=3,360; HSD N=546; All of Us N=354,400; ns, not significant; SVT, supraventricular tachycardia; PFO, patent foramen ovale; ASD, atrial septal defect; SCAD, spontaneous coronary artery dissection
† p values were calculated using Chi-square tests, with Bonferroni correction applied to adjust for multiple comparisons.
– indicates the response was excluded from analysis.

#### Endocrine

Endocrine disorders were reported in a subset of participants, with 26.0% of hEDS and 20.3% of HSD participants reporting one or more endocrine disorders (**Table 18**). Notably, there were no significant differences between the hEDS and HSD groups. Hypothyroidism (non-autoimmune) was the most common diagnosis (11.6% hEDS, 9.0% HSD), as well as osteopenia (10.5% hEDS, 7.7% HSD), and Hashimoto’s thyroiditis (6.9% hEDS, 5.5% HSD). While not common, hEDS participants reported a significantly higher prevalence than the All of Us group in conditions including central adrenal insufficiency (non-autoimmune), hyperthyroidism (non-autoimmune), gestational diabetes, pineal cyst, pituitary adenoma, and Grave’s disease (each *p*<0.0001). hEDS participants reported lower rates than the All of Us group for osteoporosis, type 2 diabetes, and hyperparathyroidism (each *p*<0.0001).

**Table 18.** Prevalence of Endocrinological Diagnoses in hEDS, HSD, and All of Us.

| Endocrinological Disorders | hEDS<br>n (%) | HSD<br>n (%) | hEDS vs.<br>HSD<br>p-Value† | All of Us<br>n (%) | hEDS vs. All<br>of Us<br>p-Value† |
| --- | --- | --- | --- | --- | --- |
| Hypothyroidism | 389 (11.6) | 49 (9) | ns | 36920 (10.4) | ns |
| Osteopenia | 354 (10.5) | 42 (7.7) | ns | 3220 (0.9) | <0.0001 |
| Hashimoto's disease | 231 (6.9) | 30 (5.5) | ns | 2880 (0.8) | <0.0001 |
| Osteoporosis | 189 (5.6) | 18 (3.3) | ns | 32160 (9.1) | <0.0001 |
| Adrenal insufficiency | 104 (3.1) | 18 (3.3) | ns | 2240 (0.6) | <0.0001 |
| Hyperthyroidism | 83 (2.5) | 6 (1.1) | ns | 740 (0.2) | <0.0001 |
| Diabetes, type 2 | 82 (2.4) | 14 (2.6) | ns | 74920 (21.1) | <0.0001 |
| Diabetes, gestational | 77 (2.3) | 7 (1.3) | ns | 3180 (0.9) | <0.0001 |
| Pineal cyst | 62 (1.8) | 5 (0.9) | ns | 40 (0) | <0.0001 |
| Pituitary tumor/adenoma | 56 (1.7) | 6 (1.1) | ns | 200 (0.1) | <0.0001 |
| Grave's disease | 33 (1) | 6 (1.1) | ns | 300 (0.1) | <0.0001 |
| Hyperparathyroidism | 27 (0.8) | 5 (0.9) | ns | 10560 (3) | <0.0001 |
| Diabetes, type 1 | 23 (0.7) | 1 (0.2) | - | - | - |
| Cushing disease | 22 (0.7) | 1 (0.2) | - | - | - |
| Addison's disease | 12 (0.4) | 1 (0.2) | - | - | - |
| Congenital adrenal hyperplasia | 9 (0.3) | 1 (0.2) | - | - | - |
| Multiple endocrine neoplasia | 2 (0.1) | 0 (0) | - | - | - |
| None of the above | 2485 (74) | 435 (79.7) | - | - | - |

hEDS N=3,360; HSD N=546; All of Us N=354,400; ns, not significant
† p values were calculated using Chi-square tests, with Bonferroni correction applied to adjust for multiple comparisons.
– indicates the response was excluded from analysis.

#### Hematologic

Most participants (88.4% hEDS, 93.4% HSD) reported no formal hematologic diagnoses (**Table 19**). Pernicious anemia was the most frequently reported condition (4.6% hEDS, 2.6% HSD) in both hEDS and HSD groups, followed by deep vein thrombosis (3.5% hEDS, 1.1% HSD). The only difference when comparing hEDS and the All of Us data was a higher prevalence of pernicious anemia (*p*<0.0001). Easy or severe bruising was reported by 80.2% of hEDS participants and 61.7% of HSD participants (*p*<0.0001).

**Table 19.**
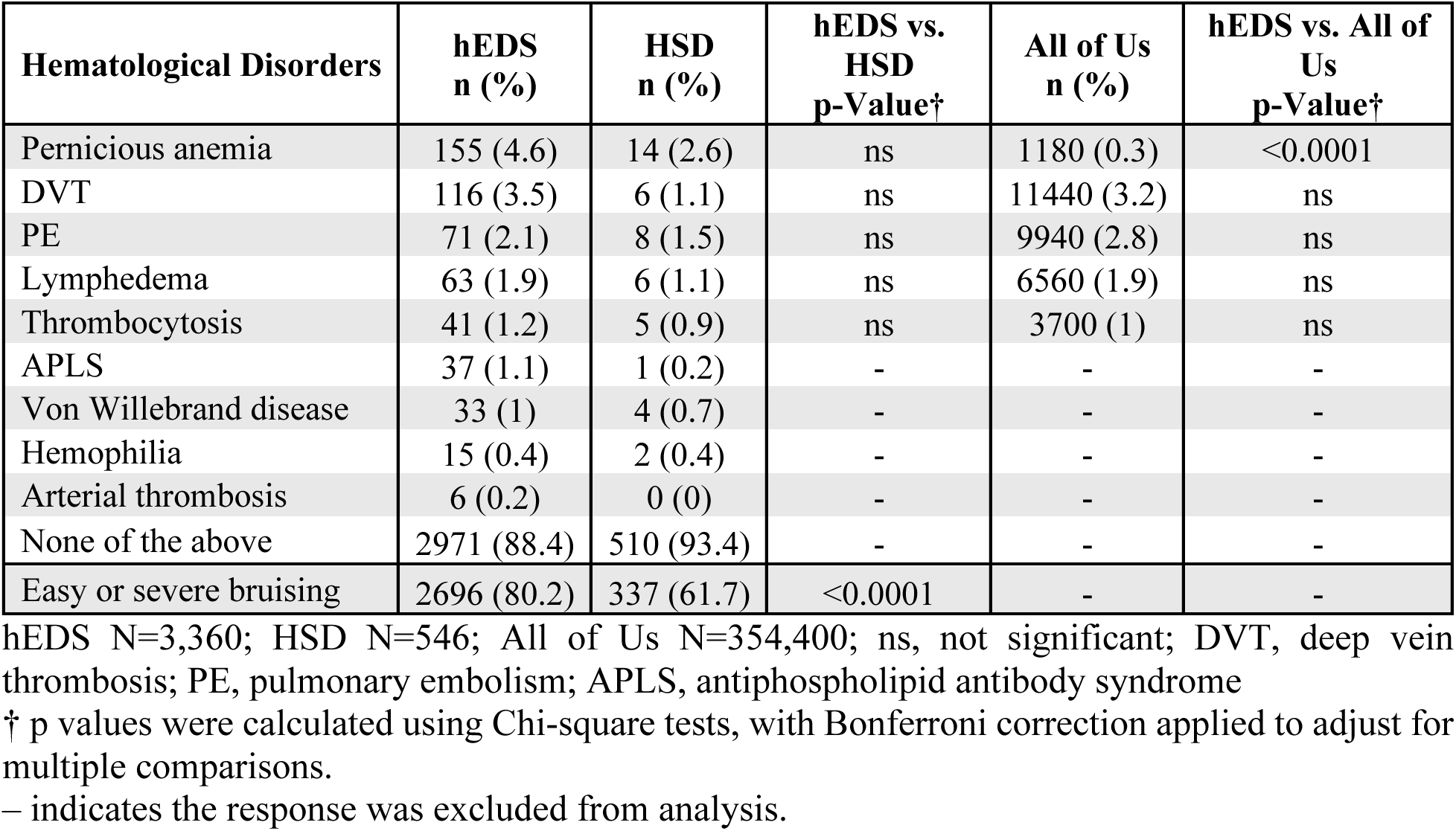
Prevalence of Hematological Diagnoses and Bruising in hEDS, HSD, and All of Us.

#### Reproductive

Reproductive health concerns were prevalent among participants, with conditions such as endometriosis (18.5% hEDS, 12.1% HSD) and polycystic ovary syndrome (PCOS) (17.0% hEDS, 14.7% HSD) frequently reported (**Table 20**). Vaginismus (7.5% hEDS, 5.3% HSD) and vulvodynia (7.0% hEDS, 5.7% HSD) were reported at similar frequencies in both groups, but notably, pelvic organ prolapse was significantly more common in hEDS participants (13.5%) compared to HSD (3.5%) (*p*<0.0001). Erectile dysfunction was the most frequently reported reproductive condition in participants assigned male at birth (12.8% hEDS, 18.8% HSD). The most frequently reported reproductive symptoms included pelvic pain (80.4% hEDS, 66.1% HSD), irregular periods (70.3% hEDS, 56.8% HSD), pain during sex (63.4% hEDS, 50.2% HSD), and bleeding during sex (38.2% hEDS, 25.5% HSD) (each *p*<0.0001) (**Table 21**). Among those assigned female at birth, participants experienced their first menstrual period at ages 12.3 (hEDS) and 12.4 (HSD) on average. Of participants who have been pregnant, the majority from both groups reported pregnancy complications, although hEDS participants reported a significantly higher prevalence (67.2% hEDS, 54.8% HSD, *p*<0.0001). Pregnancy complications included spontaneous abortion (39.8% hEDS, 30.6% HSD, *p*<0.05), preterm labor (23.2% hEDS, 12.9% HSD, *p*<0.05), failure to progress (21.98% hEDS, 20.4% HSD), premature rupture of membranes (12.2% hEDS, 10.8% HSD), and stillbirth (2.7% hEDS, 2.2% HSD).

**Table 20.** Prevalence of Reproductive Health Conditions in hEDS, HSD, and All of Us.

| Reproductive Disorders | hEDS<br>n (%) | HSD<br>n (%) | hEDS vs.<br>HSD<br>p-Value† | All of Us<br>n (%) | hEDS vs. All of<br>Us<br>p-Value† |
| --- | --- | --- | --- | --- | --- |
| Endometriosis | 622 (18.5) | 66 (12.1) | ns | 7620 (2.2) | <0.0001 |
| PCOS | 571 (17) | 80 (14.7) | ns | 5000 (1.4) | <0.0001 |
| Pelvic organ prolapse | 455 (13.5) | 19 (3.5) | <0.0001 | 10020 (2.8) | <0.0001 |
| Vaginismus | 246 (7.3) | 28 (5.1) | ns | 320 (0.1) | <0.0001 |
| Vulvodynia | 231 (6.9) | 30 (5.5) | ns | 620 (0.2) | <0.0001 |
| Infertility | 198 (5.9) | 36 (6.6) | ns | 7340 (2.1) | <0.0001 |
| Pelvic congestion syndrome | 101 (3) | 10 (1.8) | ns | 260 (0.1) | <0.0001 |
| Hypogonadism | 11 (0.3) | 3 (0.5) | - | - | - |
| Erectile dysfunction | 10 (0.3) | 3 (0.5) | - | - | - |
| Enlarged prostate gland | 7 (0.2) | 0 (0) | - | - | - |
| Premature ejaculation | 3 (0.1) | 2 (0.4) | - | - | - |
| Peyronie's disease | 1 (0) | 0 (0) | - | - | - |
| Testicular torsion | 1 (0) | 0 (0) | - | - | - |
| Penile fracture | 1 (0) | 0 (0) | - | - | - |
| Cryptorchidism | 0 (0) | 1 (0.2) | - | - | - |

hEDS N=3,360; HSD N=546; All of Us N=354,400; ns, not significant; PCOS, polycystic ovary syndrome
† p values were calculated using Chi-square tests, with Bonferroni correction applied to adjust for multiple comparisons.
– indicates the response was excluded from analysis.

**Table 21.**
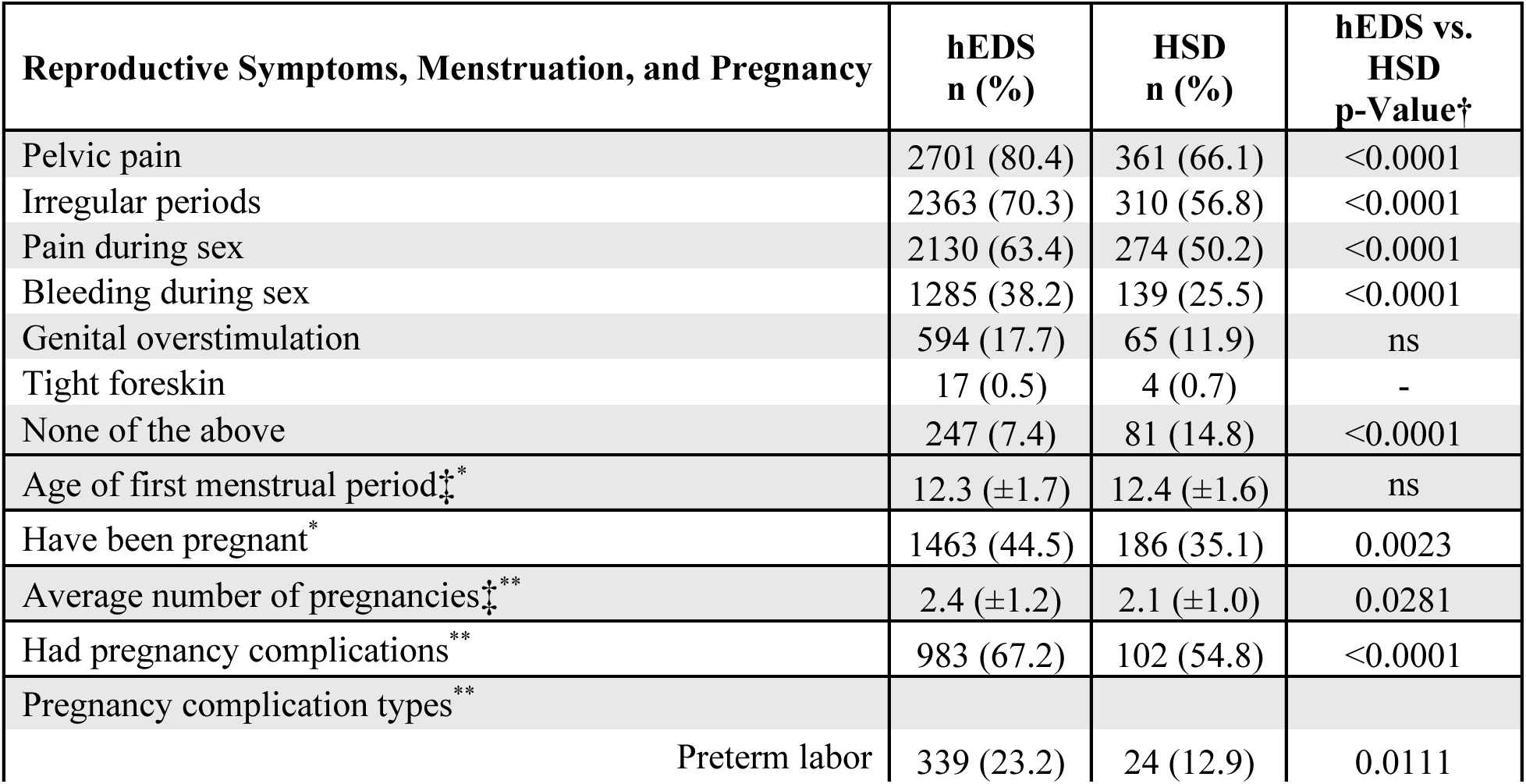
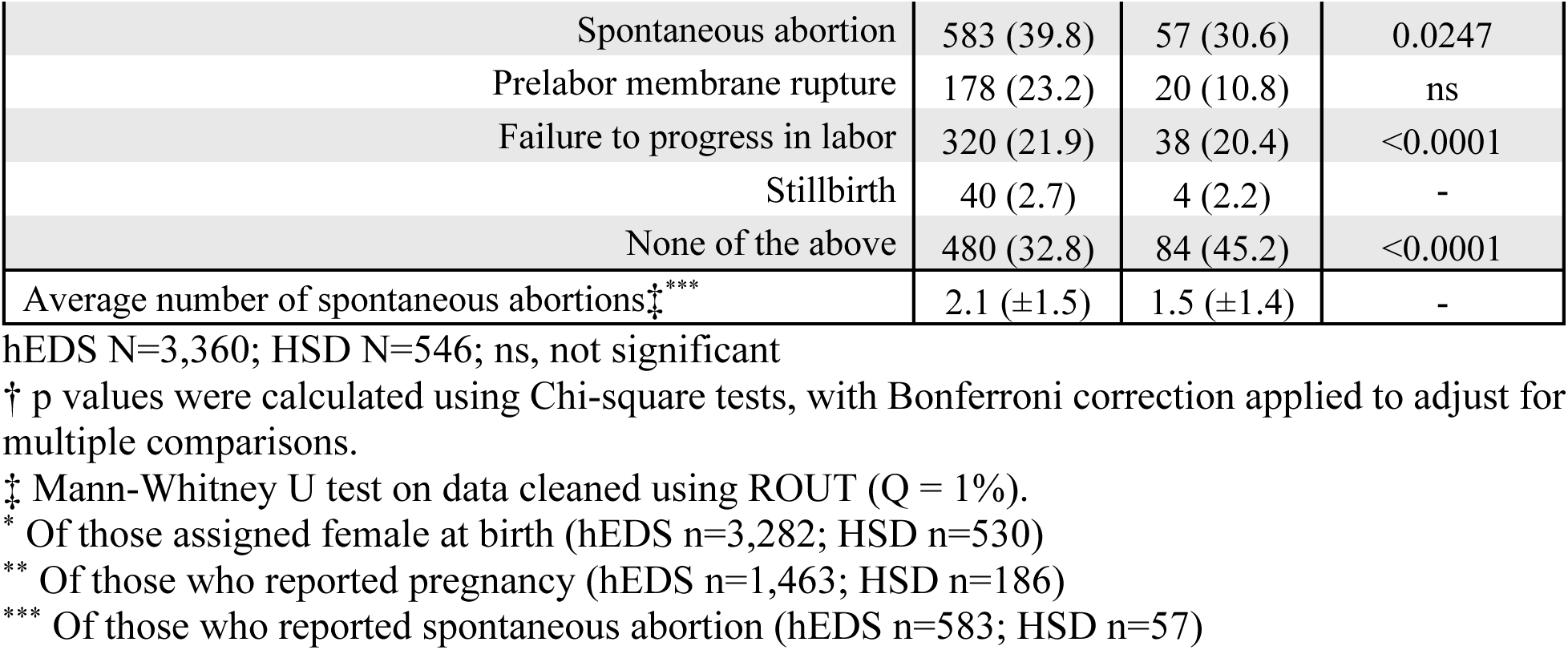
Reproductive Symptoms, Menstruation, and Pregnancy in hEDS and HSD.

#### Urological

While both groups had high rates of urinary dysfunction, hEDS participants report having at least one urinary disorder at a significantly higher rate when compared to HSD (52.5% hEDS, 35.3% HSD, *p*<0.0001) (**Table 22**). Urinary conditions reported include recurrent urinary tract infections (UTIs) (29.2% hEDS, 17.9% HSD, *p*<0.0001), urinary incontinence (21.2% hEDS, 11.2% HSD, *p*<0.0001), voiding dysfunction (14.9% hEDS, 7.9% HSD, *p*<0.01), kidney stone(s) (13.4% hEDS, 7.1% HSD, *p*<0.05), and overactive bladder syndrome (13.1% hEDS, 7.1% HSD, *p*<0.05).

**Table 22.** Prevalence of Urinary Disorders in hEDS, HSD, and All of Us.

| Urinary Disorders | hEDS<br>n (%) | HSD<br>n (%) | hEDS vs.<br>HSD<br>p-Value<sup>†</sup> | All of Us<br>n (%) | hEDS vs. All<br>of Us<br>p-Value<sup>†</sup> |
| --- | --- | --- | --- | --- | --- |
| Recurrent UTIs | 980 (29.2) | 98 (17.9) | <0.0001 | 740 (0.2) | <0.0001 |
| Urinary incontinence | 711 (21.2) | 61 (11.2) | <0.0001 | 38020 (10.7) | <0.0001 |
| Voiding dysfunction | 500 (14.9) | 43 (7.9) | 0.0065 | 60 (0) | <0.0001 |
| Kidney stone(s) | 450 (13.4) | 39 (7.1) | 0.0240 | 25060 (7.1) | <0.0001 |
| Overactive bladder syndrome | 441 (13.1) | 39 (7.1) | 0.0439 | 8580 (2.4) | <0.0001 |
| Bladder pain syndrome | 215 (6.4) | 20 (3.7) | ns | 1520 (0.4) | <0.0001 |
| Urinary hesitancy | 196 (5.8) | 18 (3.3) | ns | 3500 (1) | <0.0001 |
| Kidney disease | 85 (2.5) | 9 (1.6) | ns | 87560 (24.7) | <0.0001 |
| Vesicoureteral reflux | 27 (0.8) | 1 (0.2) | - | - | - |
| None of the above | 1597 (47.5) | 353 (64.7) | <0.0001 | - | - |

hEDS N=3,360; HSD N=546; All of Us N=354,400; ns, not significant; UTIs, urinary tract infections
† p values were calculated using Chi-square tests, with Bonferroni correction applied to adjust for multiple comparisons.
– indicates the response was excluded from analysis.

#### Dermatological

The prevalence of having at least one or more dermatological condition was similar between hEDS and HSD (40.7% hEDS, 35.5% HSD) (**Table 23**). Among them, atopic dermatitis was the most frequently reported (31.3% hEDS, 28.9% HSD), followed by hyperhidrosis (6.6% hEDS, 3.8% HSD). While there were no statistical differences between diagnoses in the hEDS and HSD groups, the hEDS group had significantly higher rates of atopic dermatitis, hyperhidrosis, and hidradenitis suppurativa when compared to the All of Us group (each *p*<0.0001). Skin-related symptoms were highly prevalent among respondents, especially the hEDS group (99.6% hEDS, 96.3% HSD, *p*<0.0001), which may reflect the inclusion of these symptoms in the 2017 hEDS diagnostic criteria (**Table 24**). Soft, velvety skin was reported by 91.0% of hEDS participants and 60.8% of HSD participants, while abnormally stretchy skin was noted in 77.4% of hEDS and 32.6% of HSD participants (each *p*<0.0001). Additionally, unexplained stretch marks were reported at significantly higher rates in hEDS (73.6%) compared to HSD (37.5%) (*p*<0.0001).

**Table 23.** Prevalence of Dermatological Disorders in hEDS, HSD, and All of Us.

| Dermatological Disorders | hEDS<br>n (%) | HSD<br>n (%) | hEDS vs.<br>HSD<br>p-Value† | All of Us<br>n (%) | hEDS vs. All of<br>Us<br>p-Value† |
| --- | --- | --- | --- | --- | --- |
| Atopic dermatitis (eczema) | 1052 (31.3) | 158 (28.9) | ns | 9540 (2.7) | <0.0001 |
| Hyperhidrosis | 223 (6.6) | 21 (3.8) | ns | 12280 (3.5) | <0.0001 |
| Hidradenitis suppurativa | 93 (2.8) | 10 (1.8) | ns | 2640 (0.7) | <0.0001 |
| Hypohidrosis | 36 (1.1) | 3 (0.5) | - | - | - |
| Acrogeria | 1 (0) | 0 (0) | - | - | - |
| None of the above | 1993 (59.3) | 352 (64.5) | ns | - | - |

hEDS N=3,360; HSD N=546; All of Us N=354,400; ns, not significant
† p values were calculated using Chi-square tests, with Bonferroni correction applied to adjust for multiple comparisons.
– indicates the response was excluded from analysis.

**Table 24.** Prevalence of Dermatological Symptoms in hEDS and HSD.

| Dermatological Symptoms | hEDS<br>n (%) | HSD<br>n (%) | hEDS vs. HSD<br>p-Value† |
| --- | --- | --- | --- |
| “Soft & velvety” skin | 3059 (91) | 332 (60.8) | <0.0001 |
| Abnormally stretchy skin | 2602 (77.4) | 178 (32.6) | <0.0001 |
| Unexplained stretch marks | 2473 (73.6) | 205 (37.5) | <0.0001 |
| Poor wound healing | 2445 (72.8) | 266 (48.7) | <0.0001 |
| Atrophic scarring | 2114 (62.9) | 149 (27.3) | <0.0001 |
| Keratosis pilaris | 1820 (54.2) | 250 (45.8) | ns |
| Recurrent Hives (urticaria) | 1636 (48.7) | 182 (33.3) | <0.0001 |
| Hypertrophic scarring | 1331 (39.6) | 147 (26.9) | <0.0001 |
| Recurrent folliculitis or abscesses | 1212 (36.1) | 132 (24.2) | <0.0001 |
| Acne (if 30+ years old) | 979 (29.1) | 133 (24.4) | ns |
| Keloid scarring | 751 (22.4) | 67 (12.3) | <0.0001 |
| None of the above | 12 (0.4) | 20 (3.7) | <0.0001 |

hEDS N=3,360; HSD N=546; ns, not significant
† p values were calculated using Chi-square tests, with Bonferroni correction applied to adjust for multiple comparisons.

#### Allergic and Immunologic

Allergic and immunologic conditions were frequently reported in both groups, with 89.7% of hEDS and 78.0% of HSD participants reporting at least one allergy-related disorder (*p*<0.0001) (**Table 25**). Allergies were present at a higher rate in hEDS participants than HSD participants (80.6% hEDS, 69.4% HSD, *p*<0.0001). Allergies in the hEDS group included seasonal (57.7%), drug (45.5%), environmental (42.7%), food (34.8%), metal (15.2%), and latex (14.2%). Asthma (42.1% hEDS, 29.9% HSD), mast cell activation syndrome (MCAS) (29.9% hEDS, 12.1% HSD), and chronic urticaria (14.7% hEDS, 4.8% HSD) were relatively frequent in both groups, although higher in hEDS participants (*p*<0.0001). Additionally, anaphylaxis episodes were reported significantly more frequently in hEDS compared to HSD (29.3% hEDS, 15.4% HSD, *p*<0.0001). In total, 29.9% of hEDS participants and 22.0% of those with HSD reported at least one of the autoimmune conditions assessed in this study. Reported autoimmune conditions with comparable prevalence between groups included psoriasis (8.0% hEDS, 5.1% HSD), rheumatoid arthritis (4.1% hEDS, 3.5% HSD), and systemic lupus erythematosus (1.8% hEDS, 1.1% HSD).

**Table 25.** Prevalence of Allergy and Immune-Related Diagnoses in hEDS, HSD, and All of Us.

| Allergy and Immune-related Disorders | hEDS<br>n (%) | HSD<br>n (%) | hEDS vs.<br>HSD<br>p-Value† | All of Us<br>n (%) | hEDS vs. All<br>of Us<br>p-Value† |
| --- | --- | --- | --- | --- | --- |
| Allergies | 2707 (80.6) | 379 (69.4) | <0.0001 | 24160 (6.8) | <0.0001 |
| Asthma | 1415 (42.1) | 163 (29.9) | <0.0001 | 65180 (18.4) | <0.0001 |
| MCAS | 1003 (29.9) | 66 (12.1) | <0.0001 | 100 (0) | <0.0001 |
| Chronic sinusitis | 899 (26.8) | 94 (17.2) | 0.0011 | 40320 (11.4) | <0.0001 |
| Chronic urticaria | 493 (14.7) | 26 (4.8) | <0.0001 | 220 (0.1) | <0.0001 |
| Histamine intolerance | 365 (10.9) | 36 (6.6) | ns | - | - |
| Immune deficiency | 243 (7.2) | 24 (4.4) | ns | - | - |
| Nasal polyps | 236 (7) | 20 (3.7) | ns | 880 (0.2) | <0.0001 |
| Cold urticaria | 170 (5.1) | 10 (1.8) | ns | 160 (0) | <0.0001 |
| Systemic mastocytosis | 19 (0.6) | 1 (0.2) | - | - | - |
| None of the above | 346 (10.3) | 120 (22) | <0.0001 | - | - |
| Types of Allergies |  |  | - | - | - |
| Seasonal | 1938 (57.7) | 248 (45.4) | <0.0001 | - | - |
| Drug | 1529 (45.5) | 166 (30.4) | <0.0001 | - | - |
| Environmental | 1435 (42.7) | 174 (31.9) | 0.0010 | - | - |
| Food | 1169 (34.8) | 142 (26) | 0.0285 | - | - |
| Metal | 510 (15.2) | 42 (7.7) | 0.0018 | - | - |
| Latex | 477 (14.2) | 40 (7.3) | 0.0064 | - | - |
| Anaphylaxis Episodes | 986 (29.3) | 84 (15.4) | <0.0001 | - | - |
| Psoriasis | 221 (6.6) | 21 (3.8) | ns | 10780 (3) | <0.0001 |
| Lyme disease | 154 (4.6) | 20 (3.7) | ns | 3120 (0.9) | <0.0001 |
| Cancer | 143 (4.3) | 16 (2.9) | ns | 65980 (18.6) | <0.0001 |
| RA | 137 (4.1) | 19 (3.5) | ns | 11380 (3.2) | ns |
| Sjögren syndrome | 127 (3.8) | 14 (2.6) | ns | 5600 (1.6) | <0.0001 |
| Ankylosing spondylitis | 73 (2.2) | 11 (2) | ns | 1360 (0.4) | <0.0001 |
| SLE | 62 (1.8) | 6 (1.1) | ns | 5020 (1.4) | ns |
| Scleroderma | 15 (0.4) | 2 (0.4) | - | - | - |
| Other | 168 (5) | 25 (4.6) | ns | - | - |
| None of the above | 2608 (77.6) | 452 (82.8) | ns | - | - |

hEDS N=3,360; HSD N=546; All of Us N=354,400; ns, not significant; MCAS, mast cell activation syndrome; RA, rheumatoid arthritis; SLE, systemic lupus erythematosus
† p values were calculated using Chi-square tests, with Bonferroni correction applied to adjust for multiple comparisons.
– indicates the response was excluded from analysis.

#### Ocular

Ophthalmologic disorders were common among both hEDS and HSD groups, with 82.1% of hEDS and 72.7% of HSD participants reporting one or more ocular disorders (**Table 26**). hEDS had notably higher prevalences, compared to All of Us data, of astigmatism, myopia, hyperopia, macular degeneration, and keratoconus (*p*<0.0001), but a similar prevalence of retinal detachment. Ophthalmologic symptoms were also common, with light sensitivity reported by 88.4% of hEDS participants and 79.5% of HSD participants (*p*<0.0001) (**Table 27**). Visual disturbances (83.4% hEDS, 73.3% HSD, *p*<0.0001), dry eyes (68.7% hEDS, 61.9% HSD), and double vision (47.8% hEDS, 29.9% HSD, *p*<0.0001) were also prevalent.

**Table 26.** Prevalence of Ocular Diagnoses in hEDS, HSD, and All of Us.

| Ocular Disorders | hEDS<br>n (%) | HSD<br>n (%) | hEDS vs. HSD<br>p-Value† | All of Us<br>n (%) | hEDS vs. All of Us<br>p-Value† |
| --- | --- | --- | --- | --- | --- |
| Astigmatism | 2139 (63.7) | 280 (51.3) | <0.0001 | 30440 (8.6) | <0.0001 |
| Myopia | 1904 (56.7) | 276 (50.5) | ns | 29380 (8.3) | <0.0001 |
| Hyperopia | 669 (19.9) | 75 (13.7) | ns | 14840 (4.2) | <0.0001 |
| Macular degeneration | 69 (2.1) | 6 (1.1) | ns | 3160 (0.9) | <0.0001 |
| Keratoconus | 42 (1.3) | 5 (0.9) | ns | 760 (0.2) | <0.0001 |
| Retinal detachment | 43 (1.3) | 6 (1.1) | ns | 4900 (1.4) | ns |
| Blue sclerae | 554 (16.5) | 19 (3.5) | - | - | - |
| Lens subluxation | 21 (0.6) | 1 (0.2) | - | - | - |
| Lens dislocation | 5 (0.1) | 0 (0) | - | - | - |
| None of the above | 600 (17.9) | 149 (27.3) | - | - | - |

hEDS N=3,360; HSD N=546; All of Us N=354,400; ns, not significant
† p values were calculated using Chi-square tests, with Bonferroni correction applied to adjust for multiple comparisons.
– indicates the response was excluded from analysis.

**Table 27.** Prevalence of Ocular Symptoms in hEDS and HSD.

| Ocular Symptoms | hEDS<br>n (%) | HSD<br>n (%) | hEDS vs. HSD<br>p-Value† |
| --- | --- | --- | --- |
| Light sensitivity | 2971 (88.4) | 434 (79.5) | <0.0001 |
| Visual disturbances | 2802 (83.4) | 400 (73.3) | <0.0001 |
| Dry eyes | 2307 (68.7) | 338 (61.9) | ns |
| Double vision | 1606 (47.8) | 163 (29.9) | <0.0001 |
| Loss of peripheral vision | 709 (21.1) | 59 (10.8) | <0.0001 |
| None of the above | 106 (3.2) | 37 (6.8) | 0.0208 |

hEDS N=3,360; HSD N=546; ns, not significant
† p values were calculated using Chi-square tests, with Bonferroni correction applied to adjust for multiple comparisons.

#### Dental

Dental issues were reported in a majority of hEDS (61.2%) and 43.2% of HSD participants (*p*<0.0001) (**Table 28**). Dental disorders were reported at high rates, including temporomandibular joint (TMJ) disorder (49.4% hEDS, 32.4% HSD, *p*<0.0001), enamel defects (19.7% hEDS, 12.5% HSD, *p*<0.05), and early onset periodontitis (17.3% hEDS, 11.7% HSD). All dental disorders included in this survey were significantly more frequent in the hEDS group than the All of Us group (each *p*<0.0001). Symptoms reported included jaw pain (81.9% hEDS, 68.7% HSD) and subluxation and/or dislocation of the temporomandibular joint (TMJ) (69.1% hEDS, 47.8% HSD) (each *p*<0.0001) (**Table 29**).

**Table 28.** Prevalence of Dental Disorders in hEDS, HSD, and All of Us.

| Dental Disorders | hEDS<br>n (%) | HSD<br>n (%) | hEDS vs.<br>HSD<br>p-Value† | All of Us<br>n (%) | hEDS vs. All of<br>Us<br>p-Value† |
| --- | --- | --- | --- | --- | --- |
| TMJ disorder | 1660 (49.4) | 177 (32.4) | <0.0001 | 11120 (3.1) | <0.0001 |
| Enamel defects | 661 (19.7) | 68 (12.5) | 0.0319 | 300 (0.1) | <0.0001 |
| Early onset periodontitis | 580 (17.3) | 64 (11.7) | ns | 340 (0.1) | <0.0001 |
| None of the above | 1302 (38.8) | 310 (56.8) | <0.0001 | - | - |

hEDS N=3,360; HSD N=546; All of Us N=354,400; ns, not significant; TMJ, temporomandibular joint
† p values were calculated using Chi-square tests, with Bonferroni correction applied to adjust for multiple comparisons.

**Table 29.**
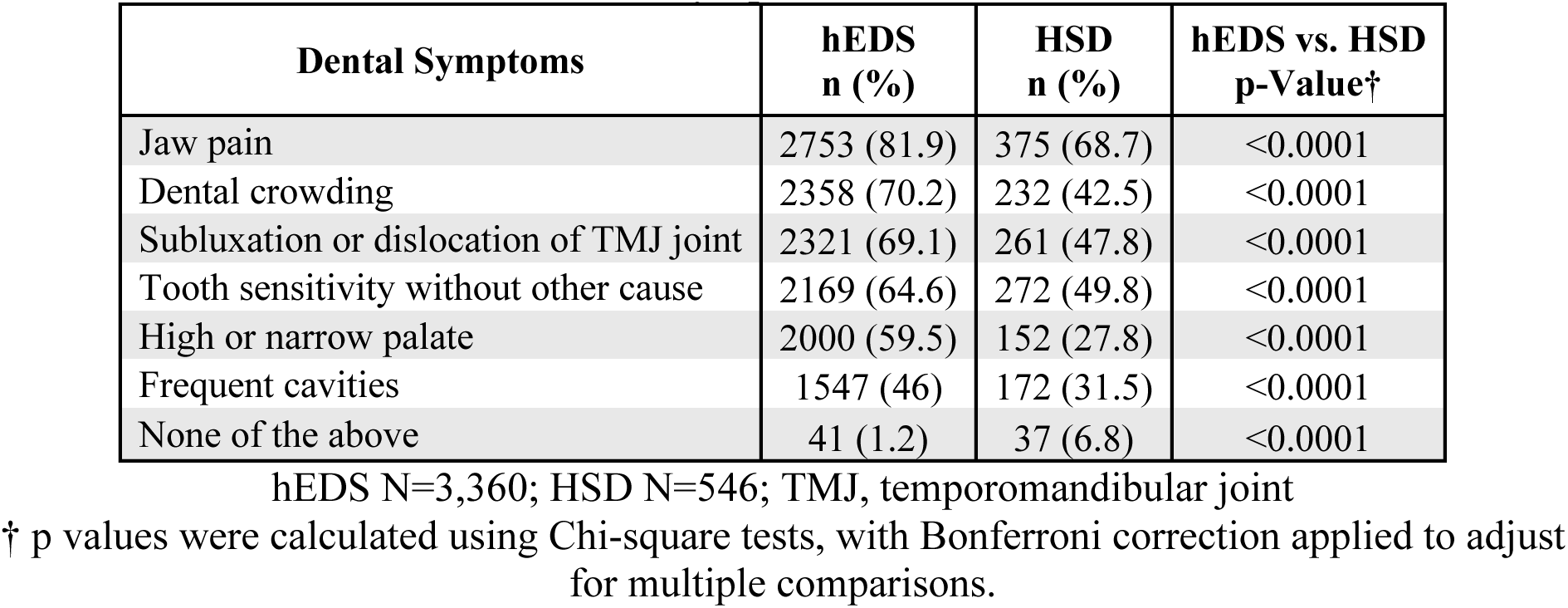
Dental Symptoms in hEDS and HSD.

#### Mental Health and Sleep

Within these populations, a high prevalence of anxiety (74.8% hEDS, 72.3% HSD), depression (68.0% hEDS, 64.5% HSD), and post-traumatic stress disorder (PTSD) (41.3% hEDS, 27.5% HSD) (*p*<0.0001) were reported (**Table 30**). Sleep disturbances were also widely reported, including insomnia (38.1% hEDS, 27.1% HSD, *p*<0.001), restless legs syndrome (16.5% hEDS, 10.3% HSD), and obstructive sleep apnea (12.7% hEDS, 9.3% HSD). Each mental health and sleep-related disorder included in this survey were reported at a significantly higher rate in hEDS compared to All of Us (each *p*<0.0001), except obstructive sleep apnea and substance use disorder were significantly lower in hEDS (each *p*<0.0001) and bipolar disorder showed no significant difference.

**Table 30.** Mental Health and Sleep Disorders in hEDS, HSD, and All of Us.

| Mental Health and Sleep Disorders | hEDS<br>n (%) | HSD<br>n (%) | hEDS<br>vs. HSD<br>p-Value† | All of Us<br>n (%) | hEDS<br>vs. All<br>of Us<br>p-Value† |
| --- | --- | --- | --- | --- | --- |
| Anxiety | 2512 (74.8) | 395 (72.3) | ns | 112320 (31.7) | <0.0001 |
| Depression | 2285 (68) | 352 (64.5) | ns | 112440 (31.7) | <0.0001 |
| PTSD | 1388 (41.3) | 150 (27.5) | <0.0001 | 21540 (6.1) | <0.0001 |
| Insomnia | 1281 (38.1) | 148 (27.1) | 0.0004 | 56320 (15.9) | <0.0001 |
| Panic disorder | 602 (17.9) | 74 (13.6) | ns | 14260 (4) | <0.0001 |
| Eating disorder | 592 (17.6) | 77 (14.1) | ns | 4820 (1.4) | <0.0001 |
| Restless leg syndrome | 553 (16.5) | 56 (10.3) | ns | 10860 (3.1) | <0.0001 |
| Obstructive sleep apnea | 427 (12.7) | 51 (9.3) | ns | 60340 (17) | <0.0001 |
| Bipolar disorder | 230 (6.8) | 13 (2.4) | 0.0386 | 20540 (5.8) | ns |
| Substance use disorder | 96 (2.9) | 15 (2.7) | ns | 57920 (16.3) | <0.0001 |
| None of the above | 409 (12.2) | 95 (17.4) | ns | - | - |

hEDS N=3,360; HSD N=546; All of Us N=354,400; PTSD, post-traumatic stress disorder
† p values were calculated using Chi-square tests, with Bonferroni correction applied to adjust for multiple comparisons.
– indicates the response was excluded from analysis.

#### Neurodivergence

Neurodivergent diagnoses were common among respondents, with 49.1% of hEDS and 39.7% of HSD participants reporting at least one neurodiversity-related disorder (*p*<0.05) (**Table 31**). The most frequently reported diagnoses included attention deficit hyperactivity disorder (ADHD/ADD) (34.9% hEDS, 28.6% HSD), autism spectrum disorder (ASD) (13.9% hEDS, 10.6% HSD), and obsessive-compulsive disorder (OCD) (13.9% hEDS, 8.2% HSD). When hEDS and All of Us data was compared, ADHD, ASD, OCD, and learning disorder were present at higher rates in hEDS than in the general population (*p*<0.0001).

**Table 31.** Neurodiversity-Related Diagnoses in hEDS, HSD, and All of Us.

| Neurodiversity-Related Disorders | hEDS<br>n (%) | HSD<br>n (%) | hEDS vs.<br>HSD<br>p-Value† | All of Us<br>n (%) | hEDS vs.<br>All of Us<br>p-Value† |
| --- | --- | --- | --- | --- | --- |
| ADHD/ADD | 1172 (34.9) | 156 (28.6) | ns | 13740 (3.9) | <0.0001 |
| ASD | 466 (13.9) | 58 (10.6) | ns | 1060 (0.3) | <0.0001 |
| OCD | 466 (13.9) | 45 (8.2) | ns | 2860 (0.8) | <0.0001 |
| Sensory processing disorder | 306 (9.1) | 34 (6.2) | - | - | - |
| Learning disorder | 299 (8.9) | 37 (6.8) | ns | 880 (0.2) | <0.0001 |
| Tourette's syndrome | 38 (1.1) | 4 (0.7) | - | - | - |
| None of the above | 1710 (50.9) | 329 (60.3) | 0.0247 | - | - |

hEDS N=3,360; HSD N=546; All of Us N=354,400; ADHD, attention deficit hyperactivity disorder; ADD, attention deficit disorder; ASD, autism spectrum disorder; OCD, obsessive compulsive disorder
† p values were calculated using Chi-square tests, with Bonferroni correction applied to adjust for multiple comparisons.
– indicates the response was excluded from analysis.

#### Multimorbidity in hEDS

To examine the multimorbidity of co-occurring disorders, a matrix was created to illustrate the percentage of participants who reported both conditions among those who reported at least one (**Fig 6**). A clear pattern emerged among condition pairs with a Jaccard index of 28.0% or greater, revealing frequent overlap between allergies, migraine, anxiety, depression, POTS, GERD, constipation, tendonitis, TMJ disorder, insomnia, PTSD, bursitis, and asthma. Notably, the strongest co-occurrences were observed across psychiatric and immunologic domains, with anxiety-depression (43.3%), anxiety-allergies (39.3%), and depression-allergies (37.2%) ranking highest. Allergies and migraine also demonstrated a high rate of co-occurrence (37.0%).

**Fig 6.**
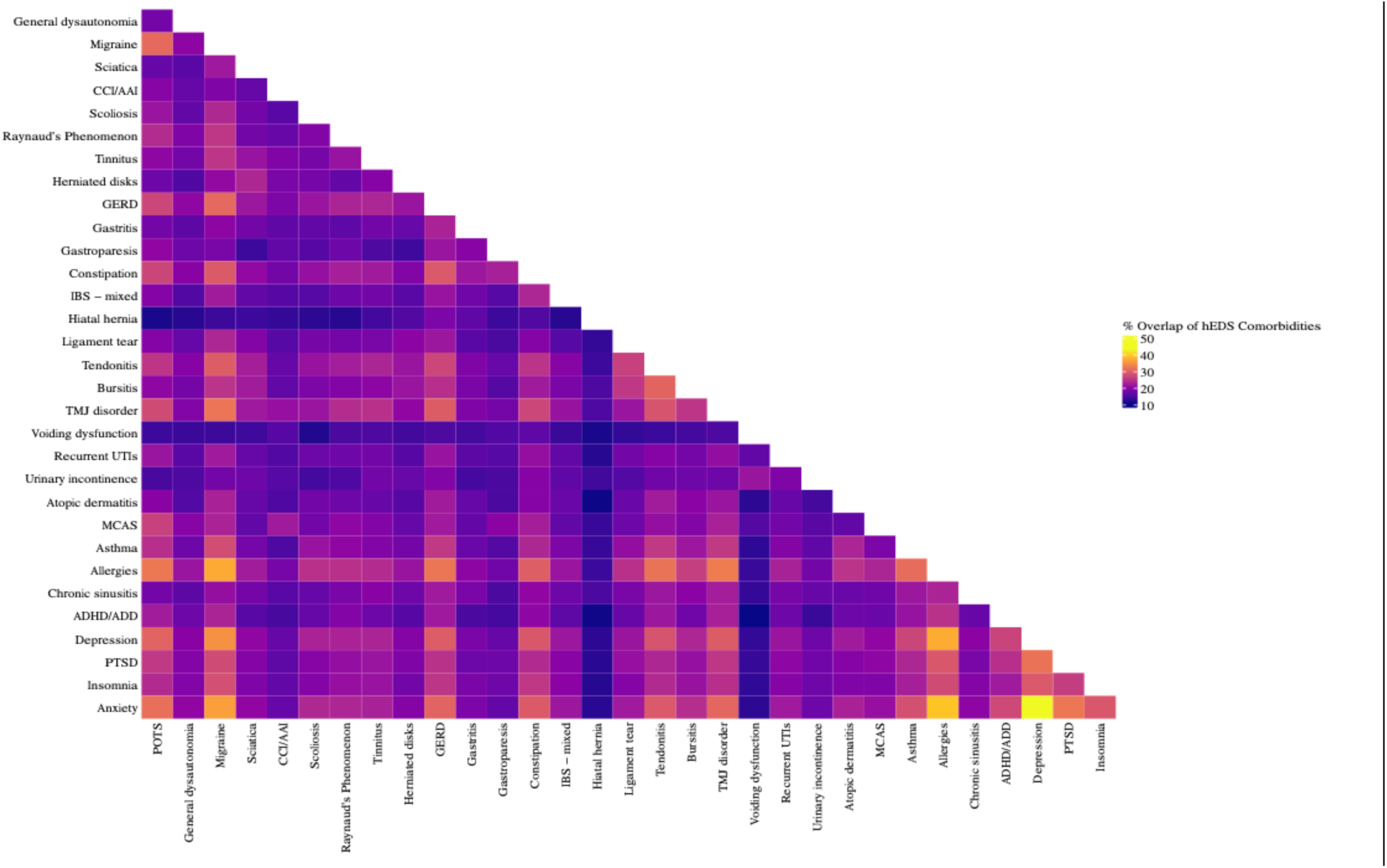
Co-occurrence matrix of comorbidities in hEDS participants. Heatmap showing pairwise overlap of formal diagnoses in hEDS participants (n = 3,360). Percent overlap was calculated using the Jaccard Index, defined as the proportion of participants who reported both conditions among those who reported either. Conditions with higher overlap included anxiety, allergies, depression, insomnia, GERD, and POTS, highlighting multimorbidity clusters in autonomic, allergic, and psychiatric domains.

## Discussion

This study provides a comprehensive analysis of the multisystemic manifestations of hEDS and HSD, leveraging a large, global cohort. These findings reveal the critical need to appropriately address gaps in the continuum from diagnosis to treatment, as well as uncovering the underlying biological mechanisms of hEDS, HSD, and their associated comorbidities. Patient responses indicate challenges beginning with the diagnostic process, where significant delays, averaging over 20 years, and misdiagnoses are common due to limited awareness among healthcare providers and complex overlap between these multisystemic conditions. The overwhelming healthcare burden associated with hEDS and HSD has been previously reported (29) and is evident in the extensive time and financial resources patients devote to managing their conditions. Many participants in our survey reported spending five or more hours per week coordinating care, seeing an average of six specialists annually, and paying significant out-of-pocket costs for necessary treatments.

### Multisystemic Manifestations

Our data support growing evidence of the substantial multisystemic involvement in hEDS and HSD, with early and severe manifestations including chronic pain, gastrointestinal symptoms, autonomic dysfunction, as well as joint instability. Autonomic dysfunction emerged as one of the most burdensome aspects of hEDS and, to a lesser extent HSD, with fatigue, dizziness, and orthostatic intolerance affecting the vast majority of participants. A significant proportion of participants also reported muscle weakness, numbness, tingling, and migraines, aligning with prior research linking hypermobility to small fiber neuropathy (SFN) and dysautonomia (30–32). Despite their inclusion in the 2017 hEDS diagnostic criteria, the prevalence of structural cardiac abnormalities, like mitral valve prolapse and aortic root dilatation, were relatively low in our dataset. This aligns with literature suggesting that autonomic and vascular dysfunction, rather than structural cardiac abnormalities, may be more clinically relevant for patients with hEDS and HSD (33).

Symptoms often attributed solely to hypermobility-related musculoskeletal issues appear to stem from coexisting conditions rather than hEDS itself. Autonomic dysfunction, MCAD, including MCAS, gastrointestinal dysmotility, and neurological complications can often drive symptom exacerbations, making it essential for healthcare providers to assess the broader clinical picture. GI symptoms were also reported at high rates in both hEDS and HSD groups, with gastroesophageal reflux disease, irritable bowel syndrome, and gastroparesis being particularly prevalent. These trends are consistent with prior research linking hypermobility to gut motility issues and visceral hypersensitivity, including studies demonstrating a higher prevalence of functional gastrointestinal disorders in individuals with hEDS and HSD (34, 35).

Chronic pain was nearly universal among participants (98.8% in hEDS, 92.7% in HSD), with an average of six painful joints per participant. In addition to joint-related pain, participants frequently reported abdominal, neuropathic, headaches, and widespread pain. Only 2% of all participants reported not experiencing chronic pain. These findings align with prior literature on the biomechanical vulnerabilities in hypermobility-related disorders and emerging theories on pain sensitization in this population (36). Despite this pain burden, conventional pharmacologic treatments remain largely ineffective, with patients frequently reporting poor responses to NSAIDs, antidepressants, and anticonvulsants. This may reflect clinical reports suggesting altered drug metabolism or receptor sensitivity in hypermobile individuals (37), underscoring the need for further investigation into pharmacogenetics, absorption differences, and alternative pain management strategies. The high prevalence of sleep disturbances further amplifies pain, fatigue, and cognitive function, highlighting the importance of targeted interventions to improve sleep.

A significant proportion of participants also reported immune-related conditions, including mast cell activation syndrome (MCAS), chronic urticaria, and heightened allergy-like responses. These findings align with prior research suggesting a potential immunological component in hypermobility disorders (37). The frequent co-occurrence of immune dysregulation, neurological, gastrointestinal, and autonomic symptoms further underscore a complex and interconnected pathophysiology that remains poorly understood.

### hEDS, HSD, and Rare Diseases

While some findings align with previous reports, the data also reveals a range of conditions beyond common comorbidities and symptoms, highlighting the overlap between hEDS, HSD, and some rare diseases. Participants in this study exhibited a disproportionately high prevalence of conditions such as Chiari malformation, cerebrospinal fluid leaks, tethered cord syndrome, adrenal insufficiency, gastroparesis, intracranial hypotension, and trigeminal neuralgia compared to the All of Us datasets. It is critical that healthcare providers consider the possibility of coexisting rare conditions in clinical settings. Relying solely on hEDS or HSD as the cause can lead to misdiagnosis and inadequate treatment, worsening patient distress and healthcare disparities.

### Challenges of the Current Diagnostic Criteria

A central finding of this study is the significant overlap in symptoms and comorbidities between hEDS and HSD. The distinction between these conditions has been historically ambiguous, leading to diagnostic uncertainty. Strikingly, 50% of survey respondents with a reported HSD diagnosis met the diagnostic criteria for hEDS, while 26% of those with an hEDS diagnosis did not meet the criteria. These findings highlight the urgent need for more precise and reliable diagnostic tools if these conditions are to be meaningfully differentiated in clinical practice. One of the most apparent distinctions between hEDS and HSD is the degree of joint hypermobility measured by Beighton score. Participants with hEDS reported significantly higher hypermobility scores, along with high rates of joint dislocations, subluxations, and orthopedic. However, the Beighton score has recognized limitations, including its narrow focus on a limited number of joints, age and sex variability and inconsistent application by clinicians (38). These limitations may contribute to misclassification or underdiagnosis, particularly in individuals with systemic features but lower Beighton scores.

While these findings likely reflect the thresholds set by the 2017 diagnostic criteria for hEDS, they should not be interpreted to mean that HSD is less clinically significant. Symptom severity was not deeply assessed, and many respondents with HSD reported substantial symptom burden and multisystem involvement. Many comorbidities such as autonomic dysfunction, mast cell activation syndrome (MCAS), gastrointestinal dysmotility, and neurological complications were common in both groups, though more frequently reported in hEDS participants. The high symptom burden in both groups compared to the general population make differential diagnosis challenging. Future research should focus on refining diagnostic criteria to ensure that individuals with disabling symptoms receive appropriate medical attention, regardless of whether they meet strict diagnostic thresholds. Expanding research into the underlying biological mechanisms of these conditions may also help clarify whether hEDS and HSD are truly separate conditions or variations within the same disorder spectrum.

### Triggering Events

Over two-thirds of participants reported an event preceding symptom onset or worsening symptom severity, such as puberty, viral infections, physical trauma, or pregnancy. These findings reinforce the hypothesis that environmental factors, in combination with genetic predisposition, may play a role in the onset of hEDS and HSD and/or disease progression. The association between viral infections and worsening symptoms further supports a potential link between hEDS, immune dysregulation, and even post-viral syndromes such as long COVID and ME/CFS. The overlap of dysautonomia, MCAS, and chronic fatigue within these conditions suggest shared mechanisms, underscoring the need for further investigation to determine whether hypermobility-related disorders have a latent component that can be exacerbated by physiological stressors.

### Limitations and Future Directions

While this study offers valuable insights, certain limitations are acknowledged. Among them is the reliance on self-reported data, which may introduce biases such as recall error and diagnostic uncertainty. However, the survey was developed with input from a multidisciplinary team of clinicians, researchers, and patients to ensure clarity, relevance, and comprehensive symptom capture, particularly important in conditions as complex and under-characterized as hEDS and HSD, where few validated instruments exist. In such contexts’, self-reported data are often the only practical means of capturing real-world, patient-centered experiences at scale. This approach is well-established in epidemiology, health services research, and rare disease registries, where it remains a critical tool for advancing understanding and improving care. Use of the 2017 criteria to filter participants may have excluded individuals with hEDS who do not meet current guidelines, and access to specialized care likely skewed responses to those with a formal diagnosis. The anonymous nature of the survey also prevented verification of unique responses. International differences in healthcare access and diagnostic practices may have also influenced how participants understood or reported their diagnoses. Furthermore, access to specialized testing (e.g., upright MRI for cranio-cervical instability) may vary significantly, potentially leading to underdiagnosis of certain conditions. These limitations highlight the urgent need for large-scale, clinician-verified studies to corroborate these findings and refine diagnostic criteria. Despite these limitations, the findings of this study underscore the significant multisystemic impact of hEDS and HSD, including chronic pain, autonomic dysfunction, immunological involvement, and numerous comorbidities. Yet, effective treatments remain elusive, leaving many patients to seek non-traditional and unproven therapies. A major challenge in developing standardized care protocols is the lack of research that clearly defines and distinguishes hEDS from HSD, particularly regarding comorbidities not included in the 2017 diagnostic criteria. While existing studies have reported comorbid conditions in hEDS/HSD, most have been limited in scope, lump the two conditions together, or lacked control populations. The findings of this work define overlapping features and distinctions between hEDS and HSD, and their prevalence compared to the general population, providing a more complete understanding of these conditions. Distinguishing between them remains a challenge, as does understanding whether they exist on a continuous spectrum or if HSD can progress to hEDS over time or with exposure to additional triggers.

This study serves as a critical call to action for the medical community. The staggering delays in diagnosis, the overwhelming reliance on self-advocacy, and the systemic lack of treatment options illustrate a glaring gap in healthcare. The lack of standardized care protocols often leads to misdiagnosis, symptom neglect, and, at times, outright denial of these conditions by healthcare providers. Addressing these issues will require collaboration across specialties, heightened awareness among healthcare providers, and a sustained commitment to addressing the full spectrum of challenges faced by this population.

### Reconsidering Etiology

While hEDS is currently classified as a heritable connective tissue disorder, findings from this study suggest that connective tissue symptoms may arise alongside, or in response to, other underlying physiological processes. The high prevalence of immune dysregulation, autonomic dysfunction, and gastrointestinal dysmotility suggests a role for immune or neuroimmune mechanisms in the broader disease process and raise the question of whether hEDS is more of an autoimmune/autoinflammatory condition than a primary connective tissue disease. This hypothesis underscores the need to expand current research efforts beyond the connective tissue paradigm and explore alternative etiologies, which may have profound implications for both diagnosis and treatment. As researchers, clinicians and patient communities advocate for updated diagnostic frameworks, expanded research efforts, and improved treatment pathways, it is imperative that patient experiences remain at the forefront of these advancements. Given the significant multimorbidity associated with hEDS and HSD, future research, particularly incorporating biomarker studies and genetic approaches, may help refine classifications and improve our ability to diagnose and treat these conditions. Recognizing the full impact of hEDS and HSD is key to driving meaningful change, breaking down barriers to care, advancing research and ensuring that individuals and families impacted by these conditions receive the medical care and support they deserve.

## Data Availability

All data produced in the present work are contained in the manuscript

## Acknowledgements

We are deeply grateful to the individuals living with Ehlers-Danlos syndrome who generously shared their experiences, completed the survey, and helped disseminate it within their communities. Their participation and insights were essential to shaping this study and will continue to inform future research directions. We extend our sincere thanks to the healthcare collaborators, advocacy organizations, and community groups that supported recruitment efforts, as well as to the dedicated committee of patient-scientists who played a critical role in the development and refinement of the survey instrument. The continued commitment and advocacy of patients remain the driving force behind this work. We also acknowledge and thank the participants of the All of Us Research Program and the National Institutes of Health for making participant data available and accessible for this study. Finally, this work would not have been possible without the generous support of the Maltz Family Foundation, The Fullerton Foundation, The Connective Tissue Coalition, and the many individual donors who have supported, and continue to support, our efforts. Their contributions have been vital to advancing the understanding of hEDS and HSD through comprehensive survey-based research, genetic and biomarker discoveries, and the pursuit of novel therapies aimed at improving outcomes for this commonly affected, yet often underserved, patient population.

---

**Supplementary Figure 1.**
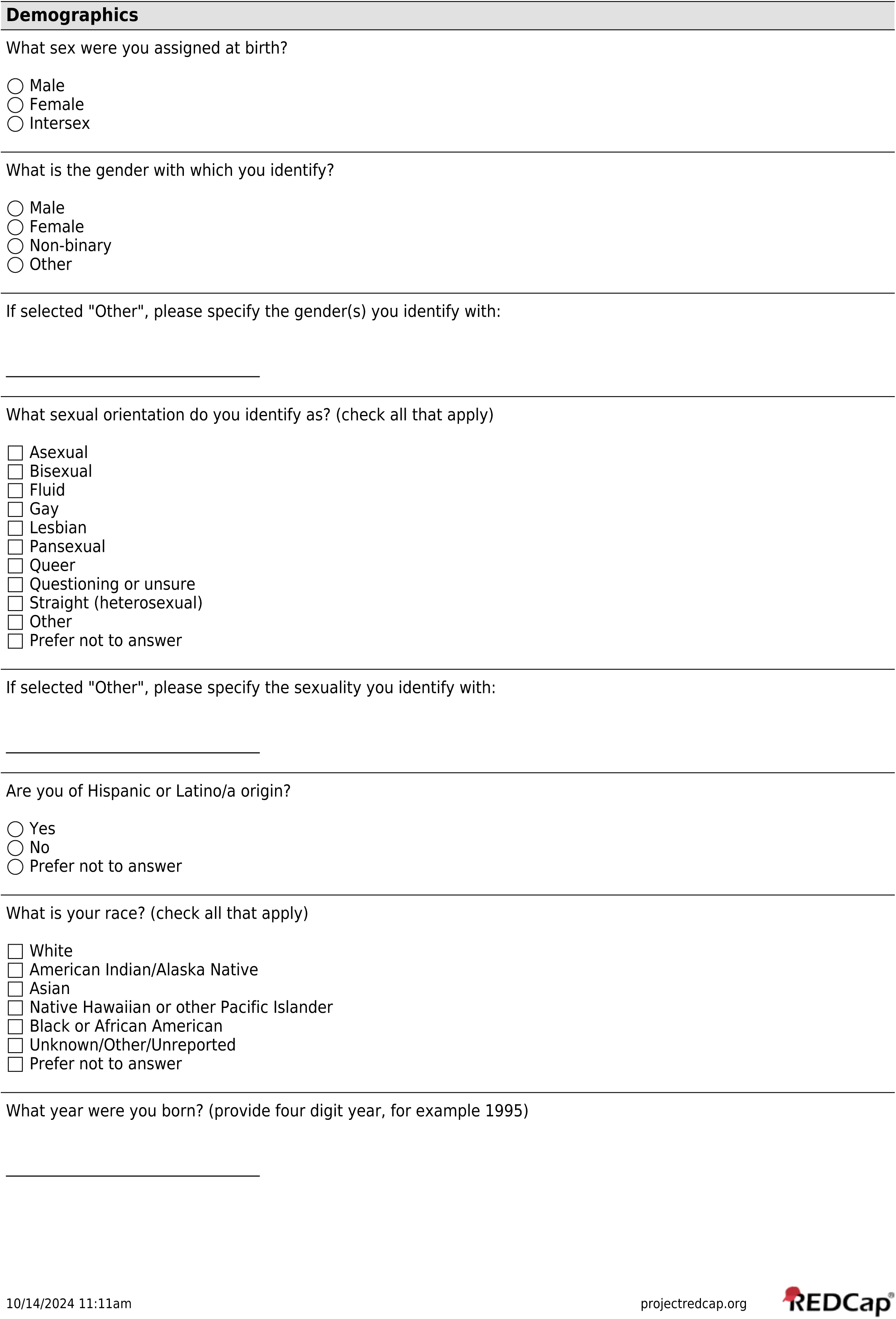
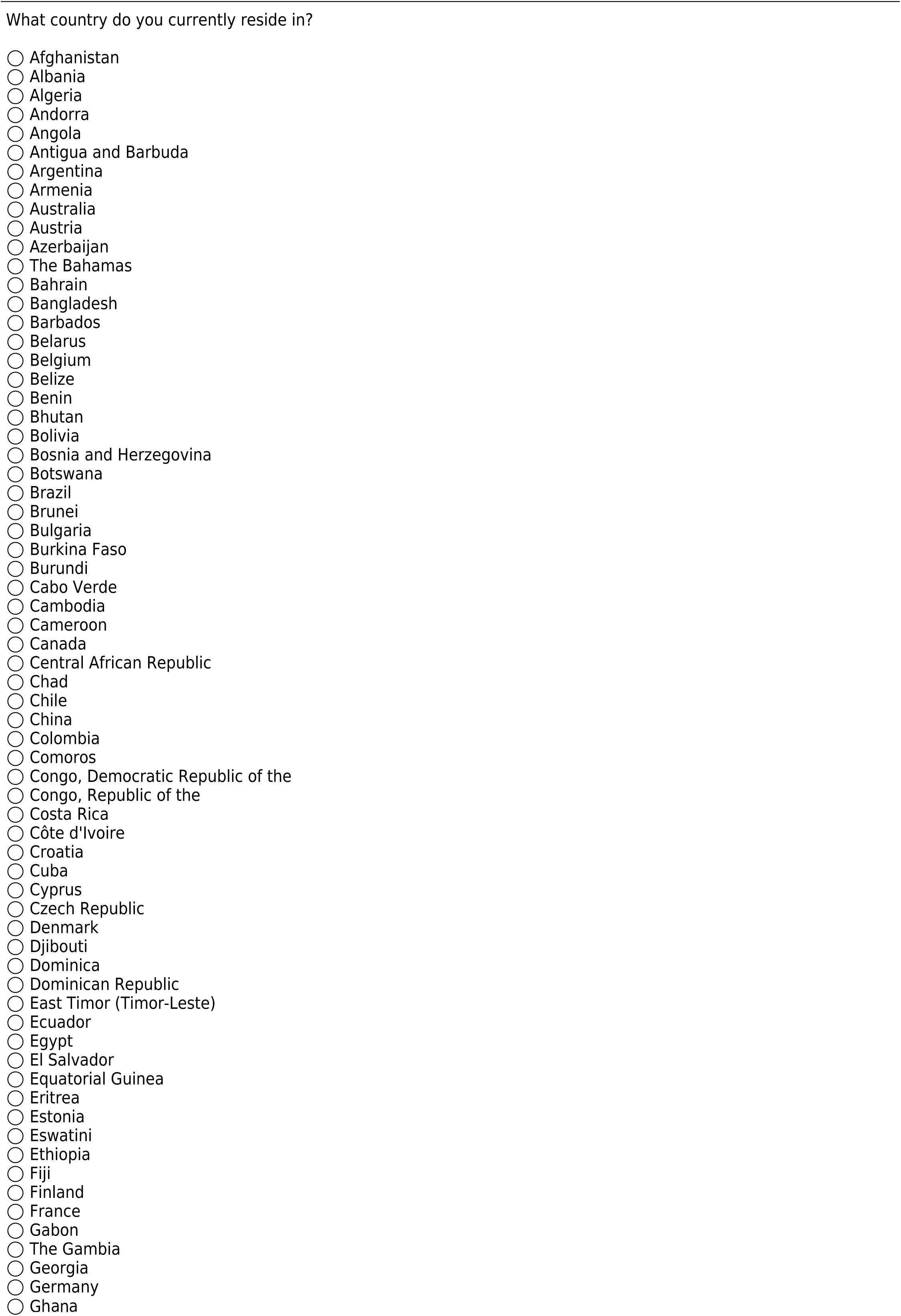
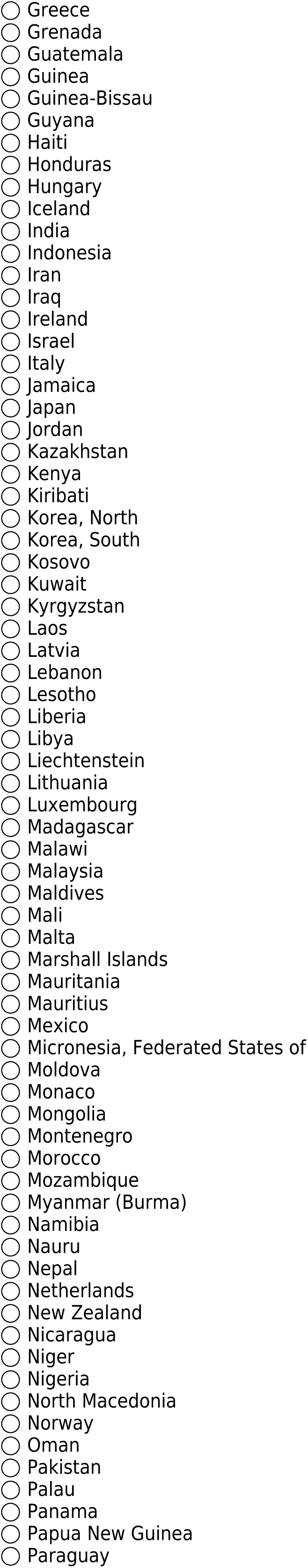
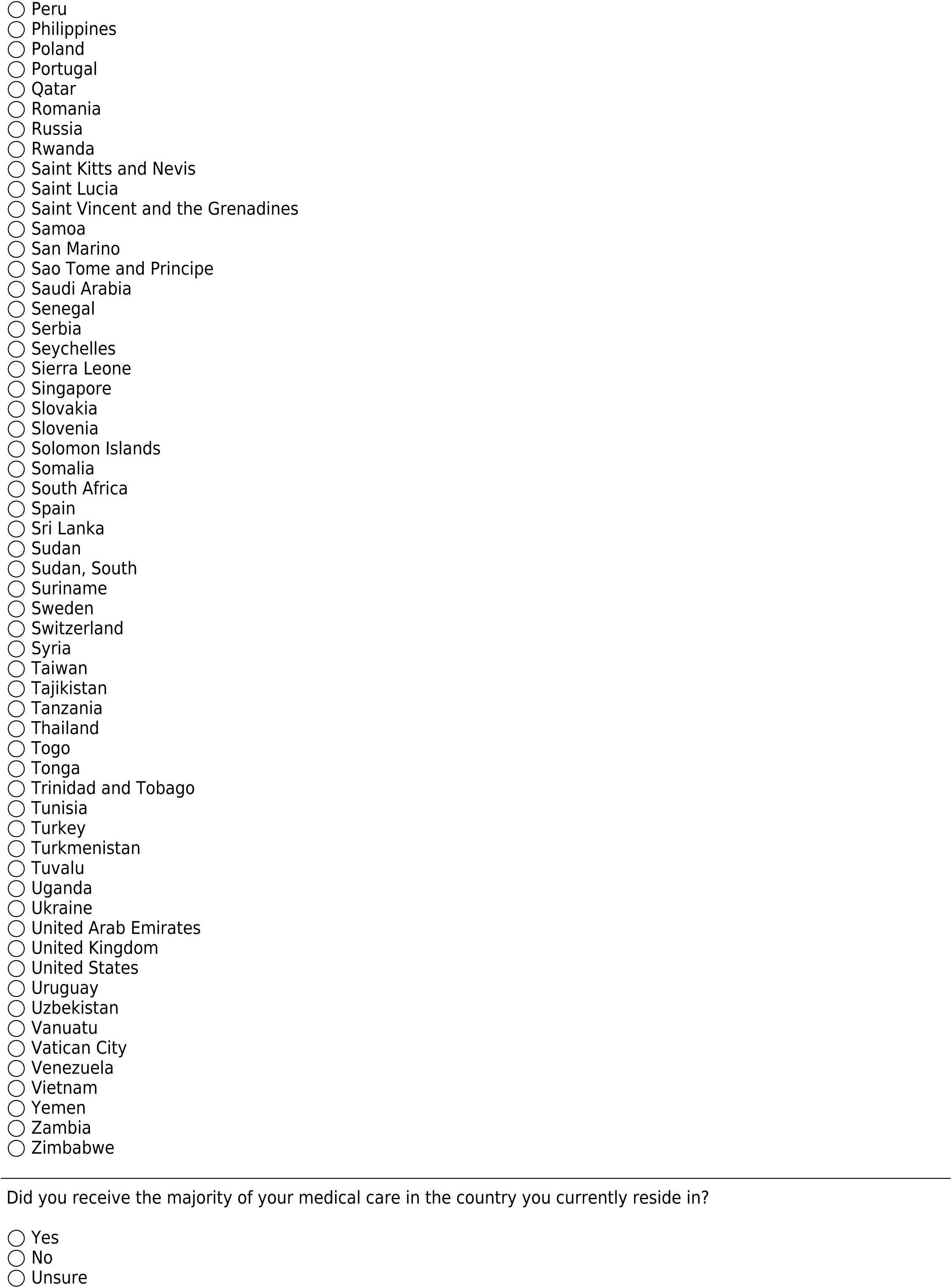
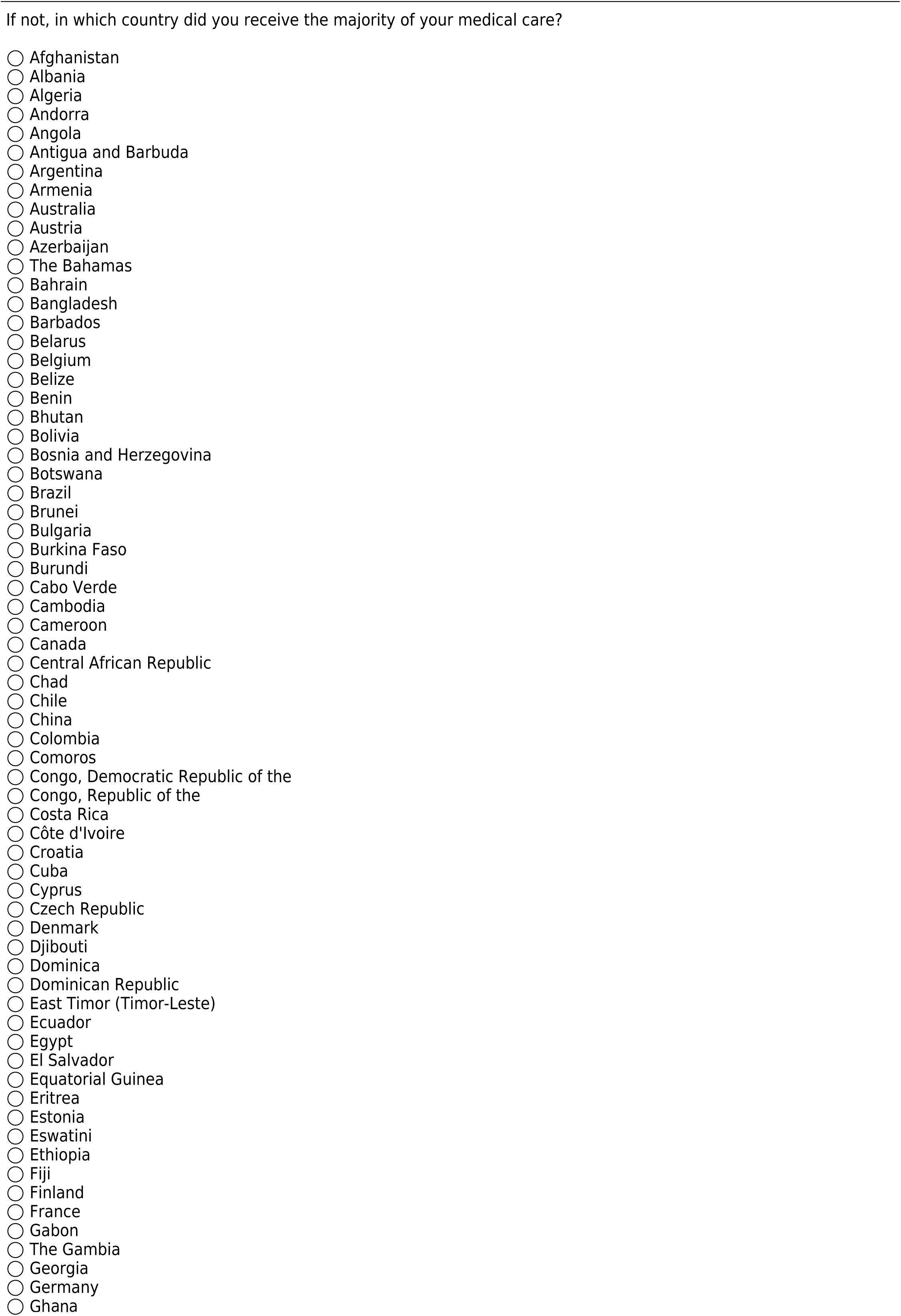
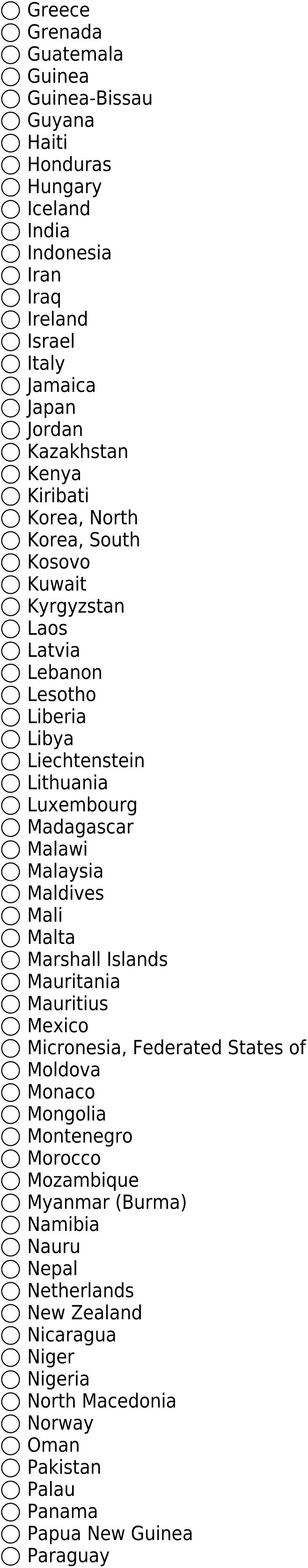
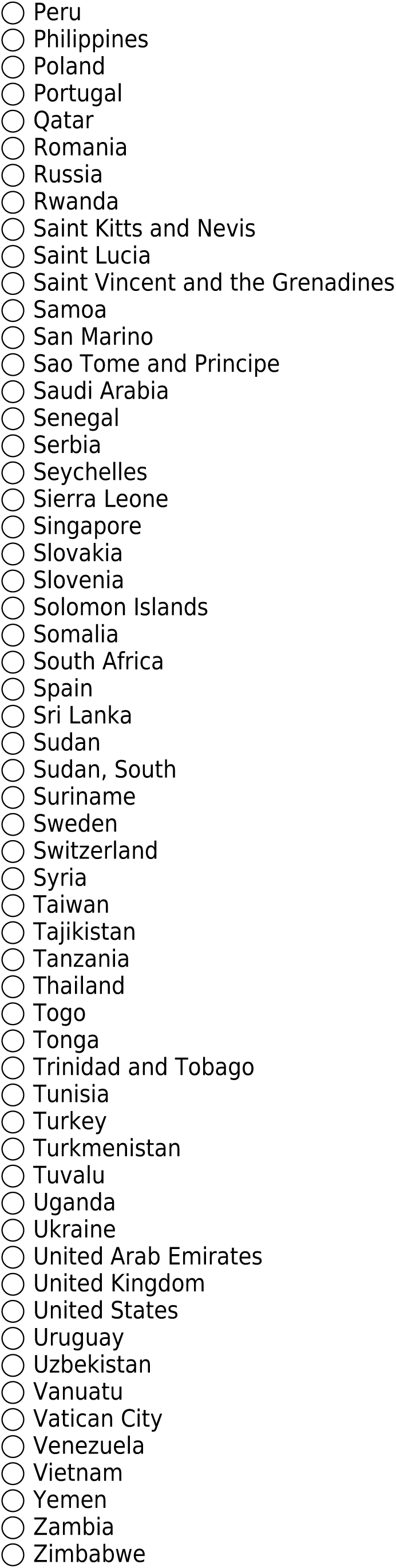
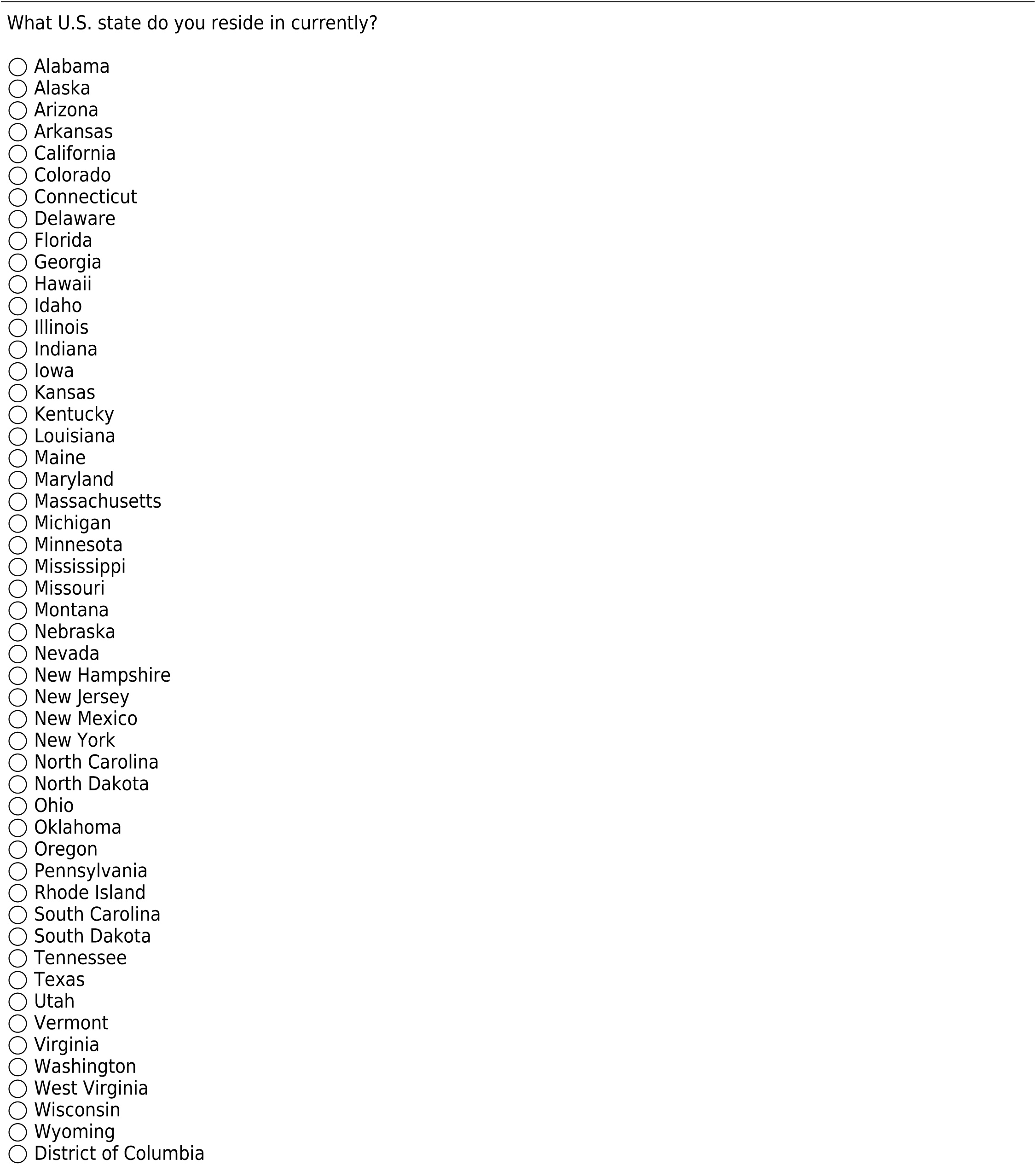
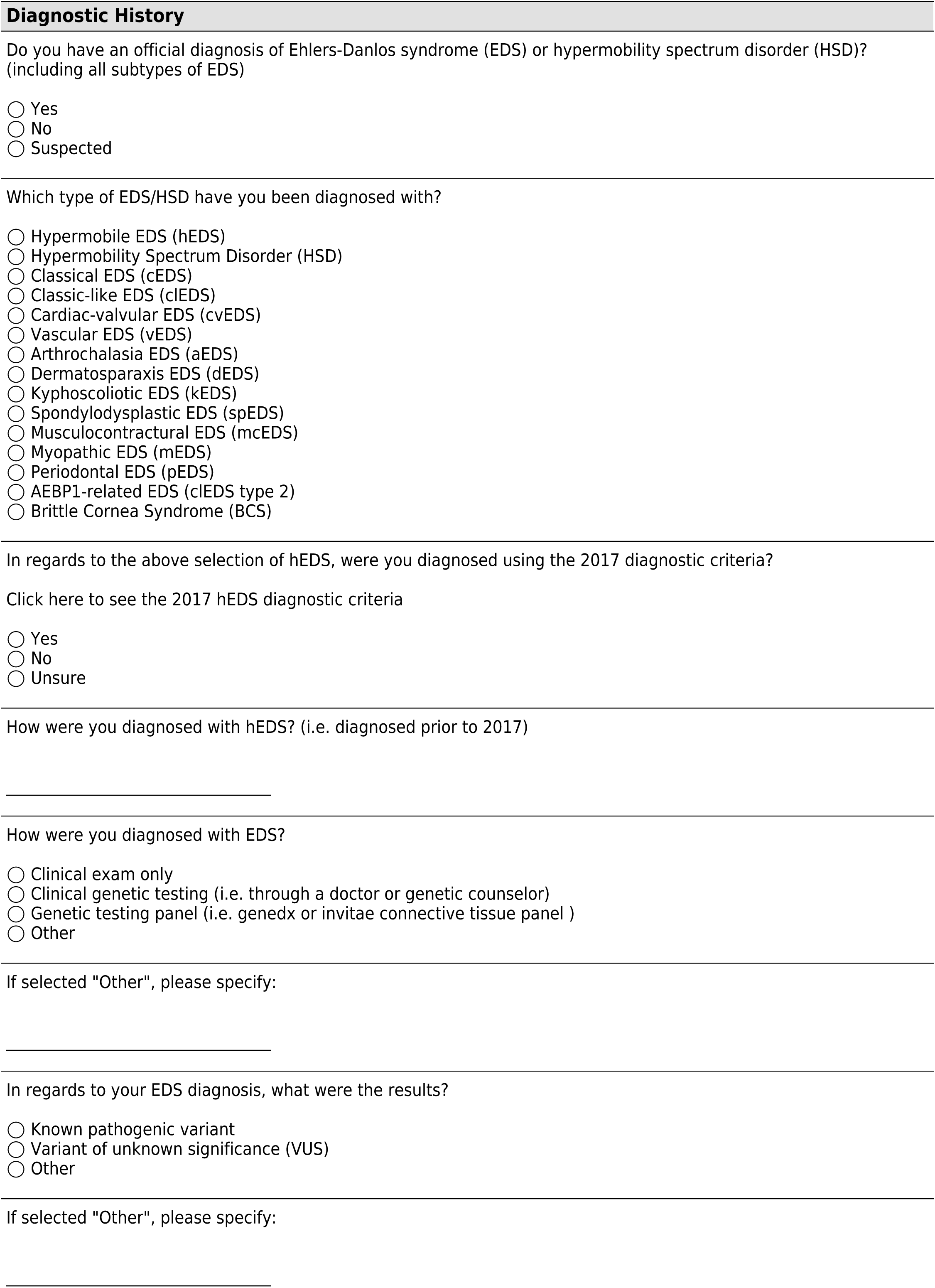
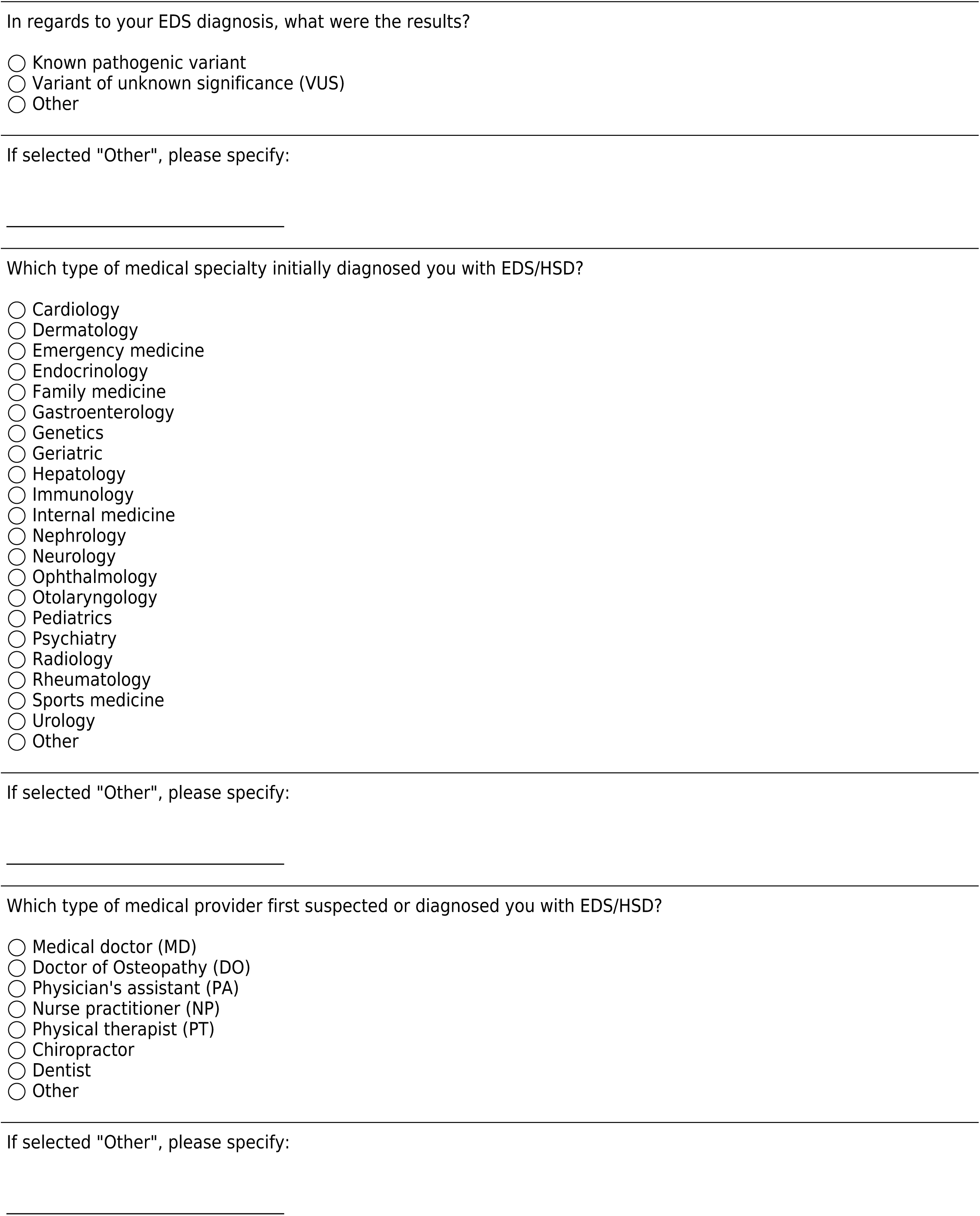
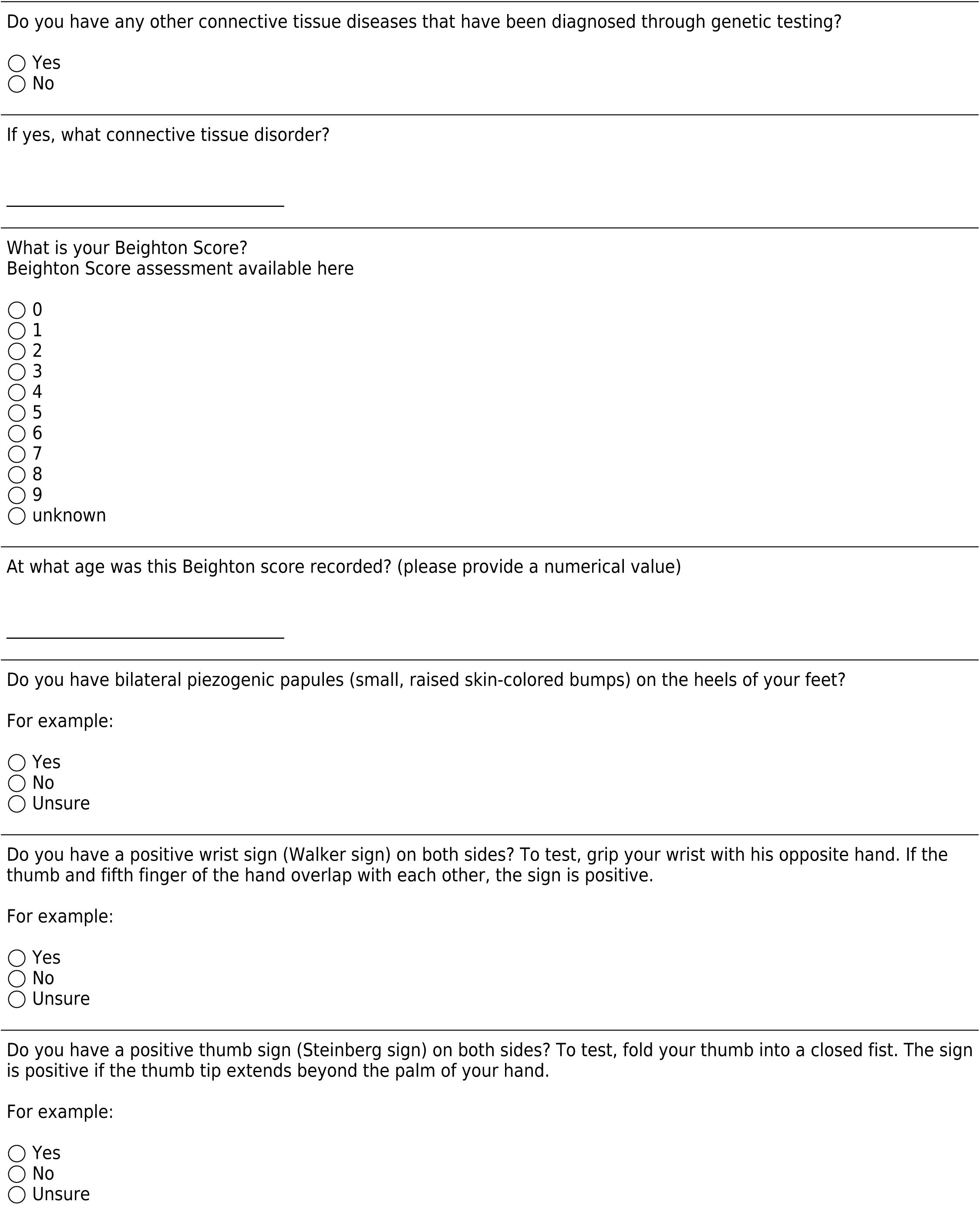
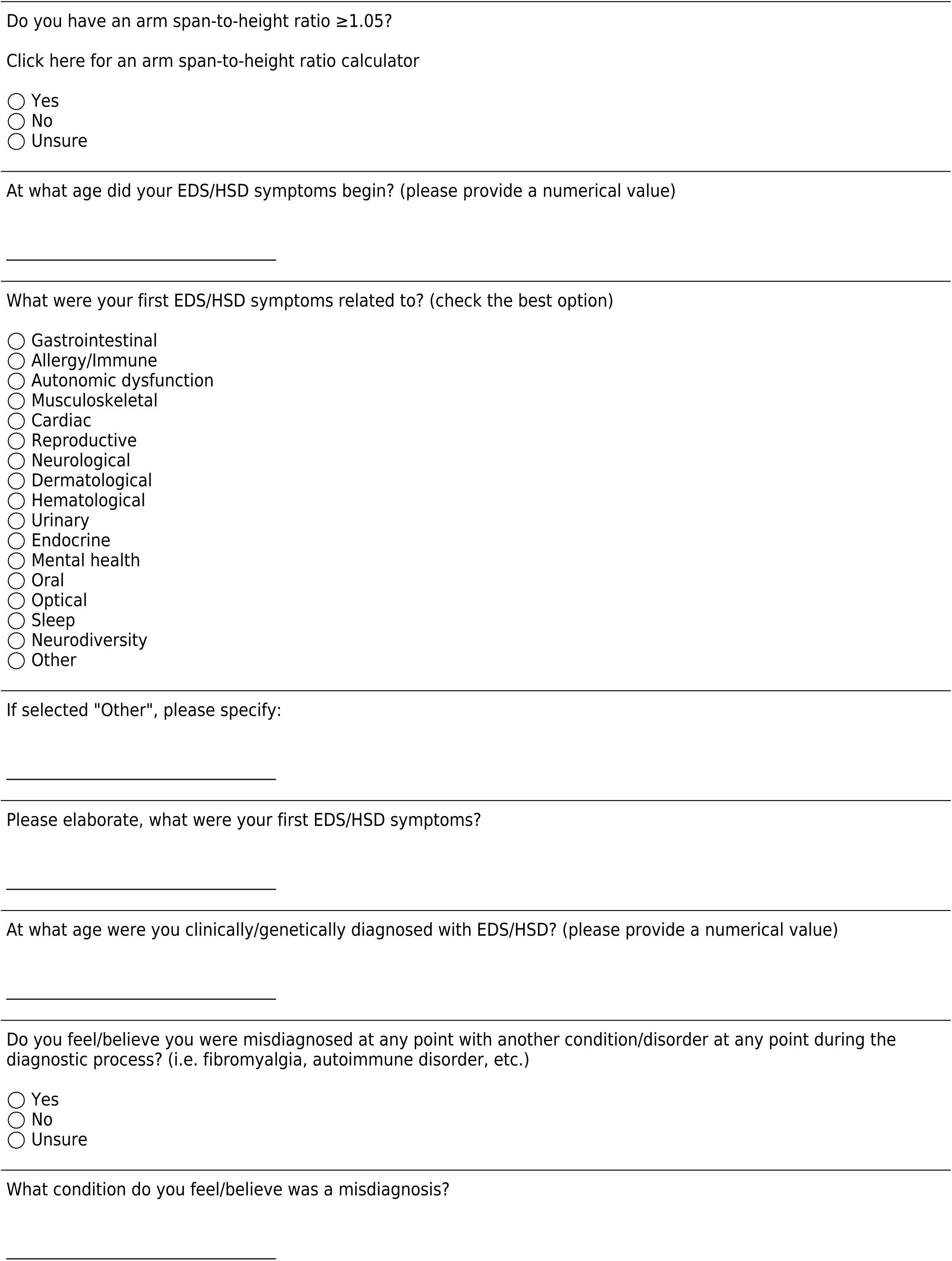
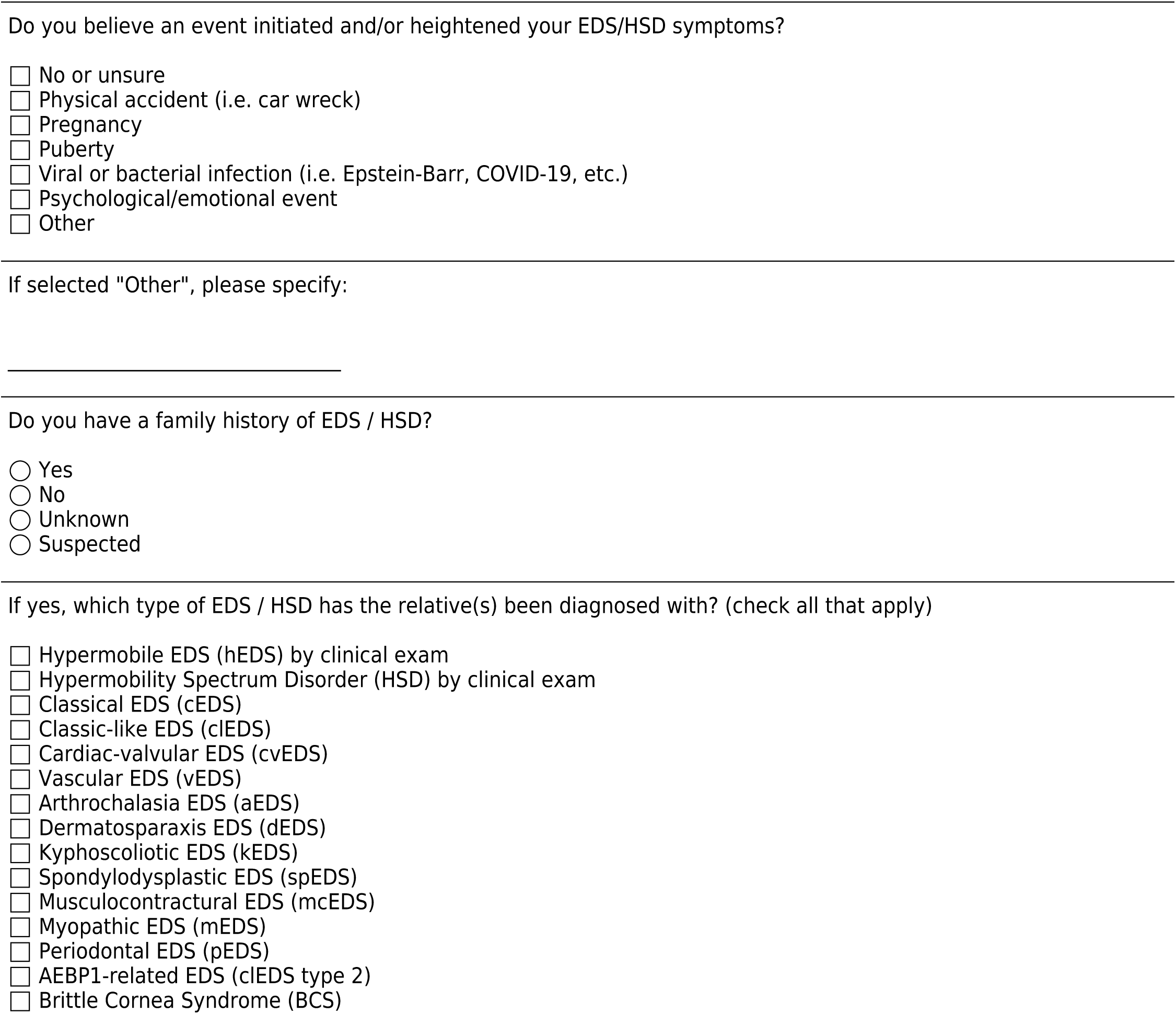
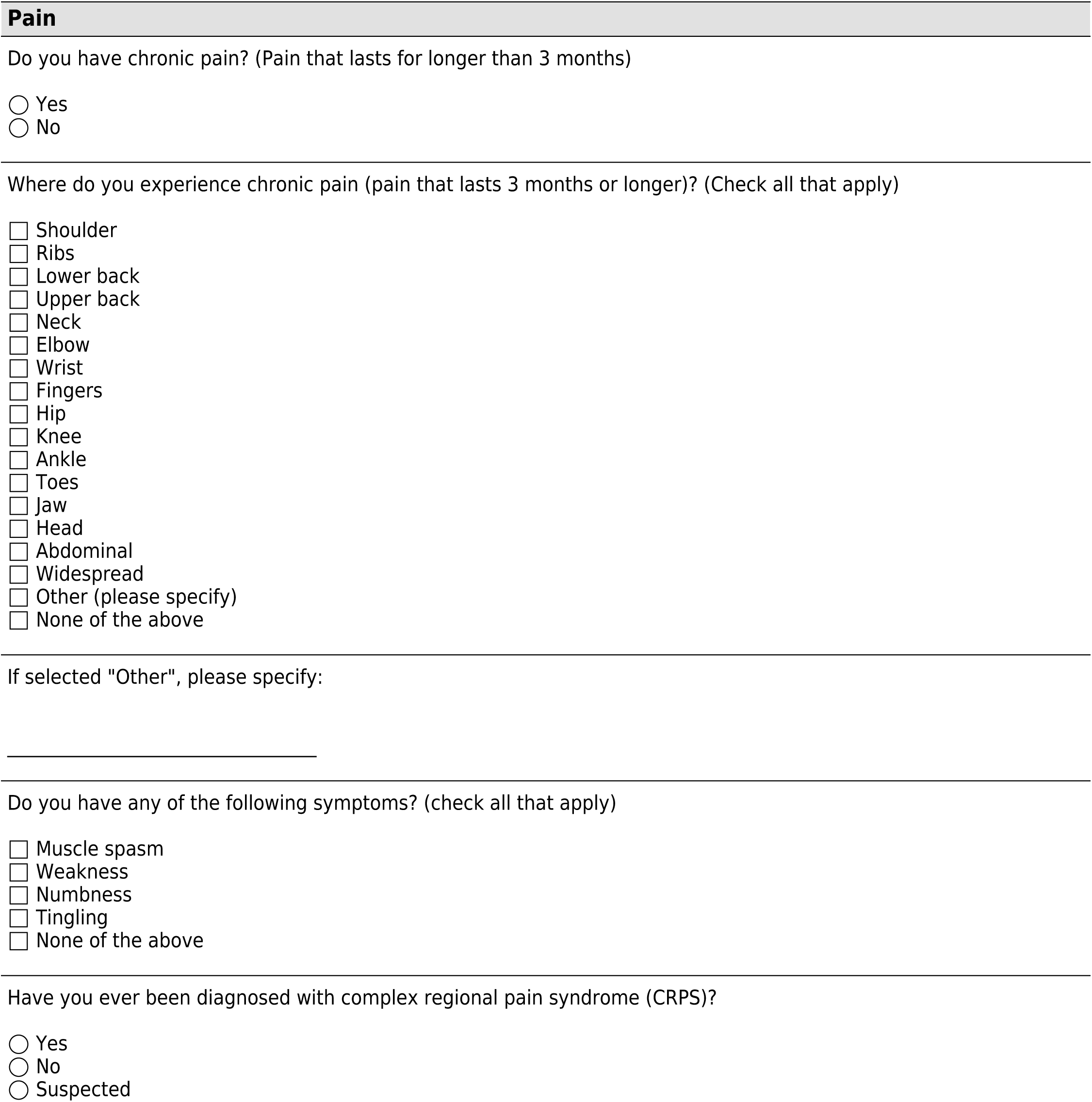
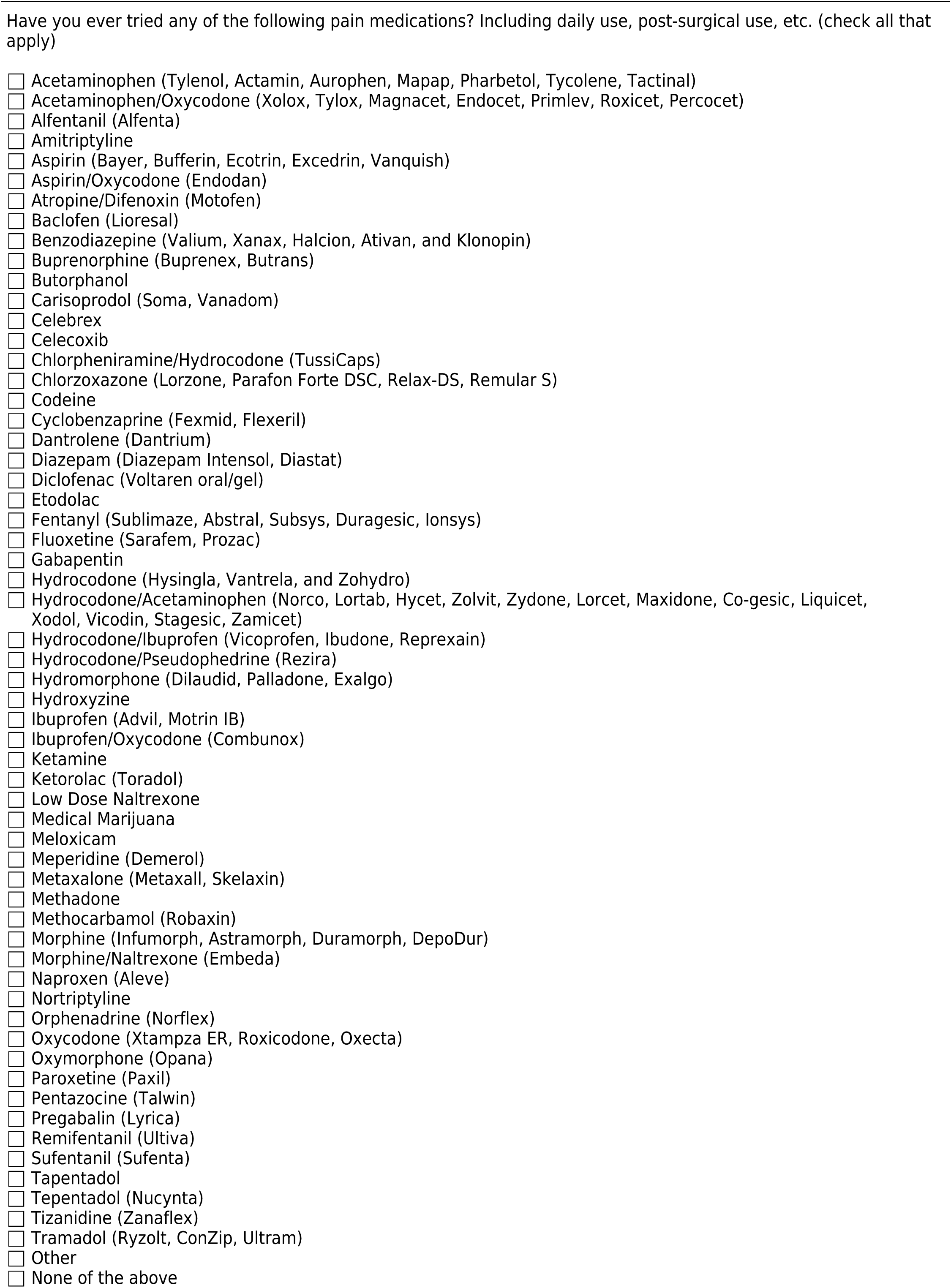
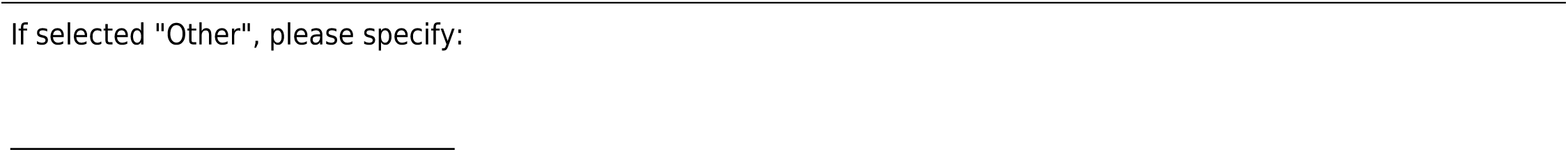
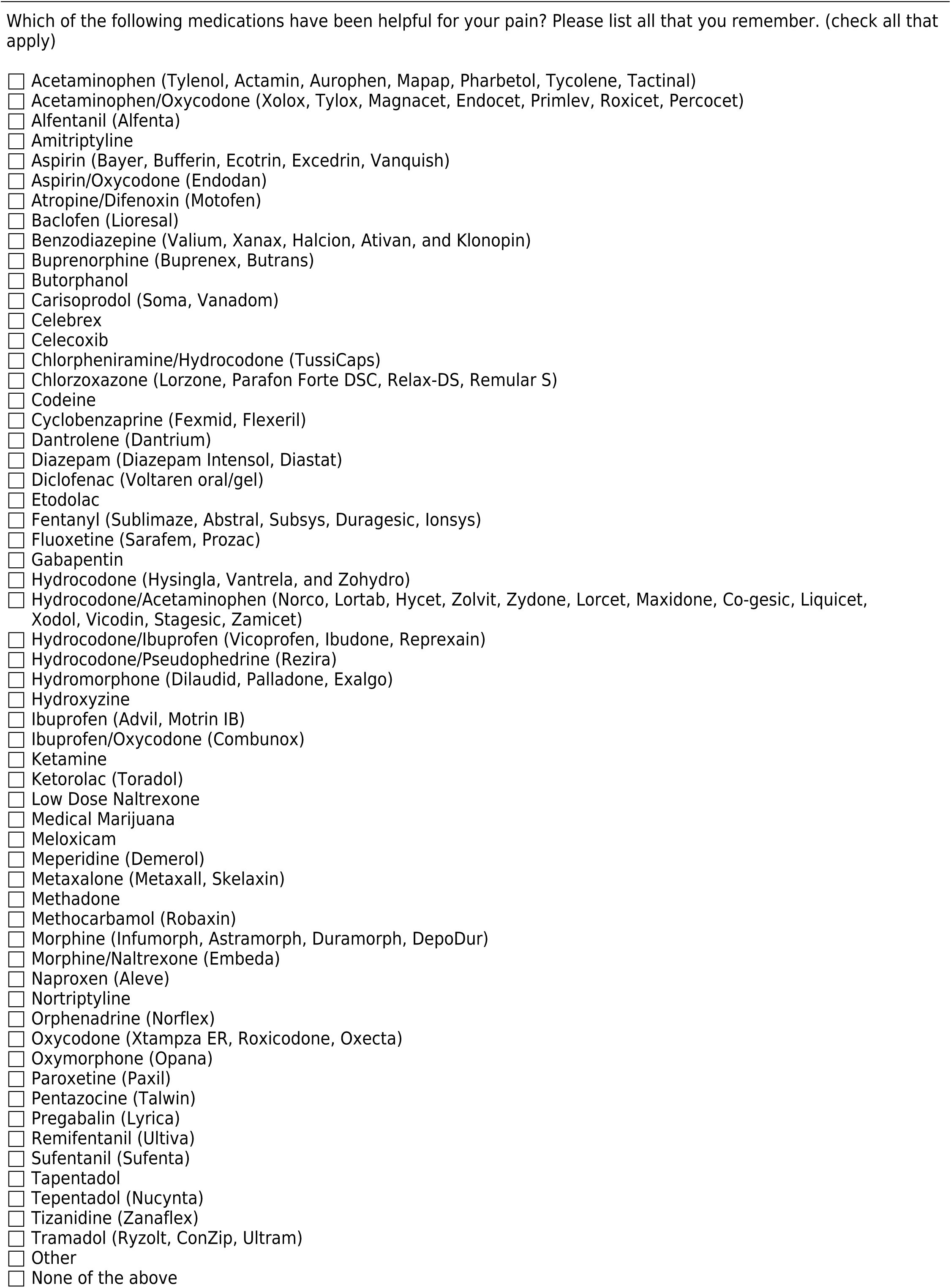
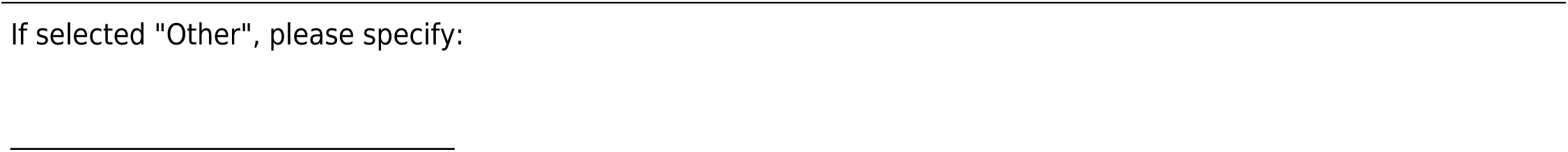
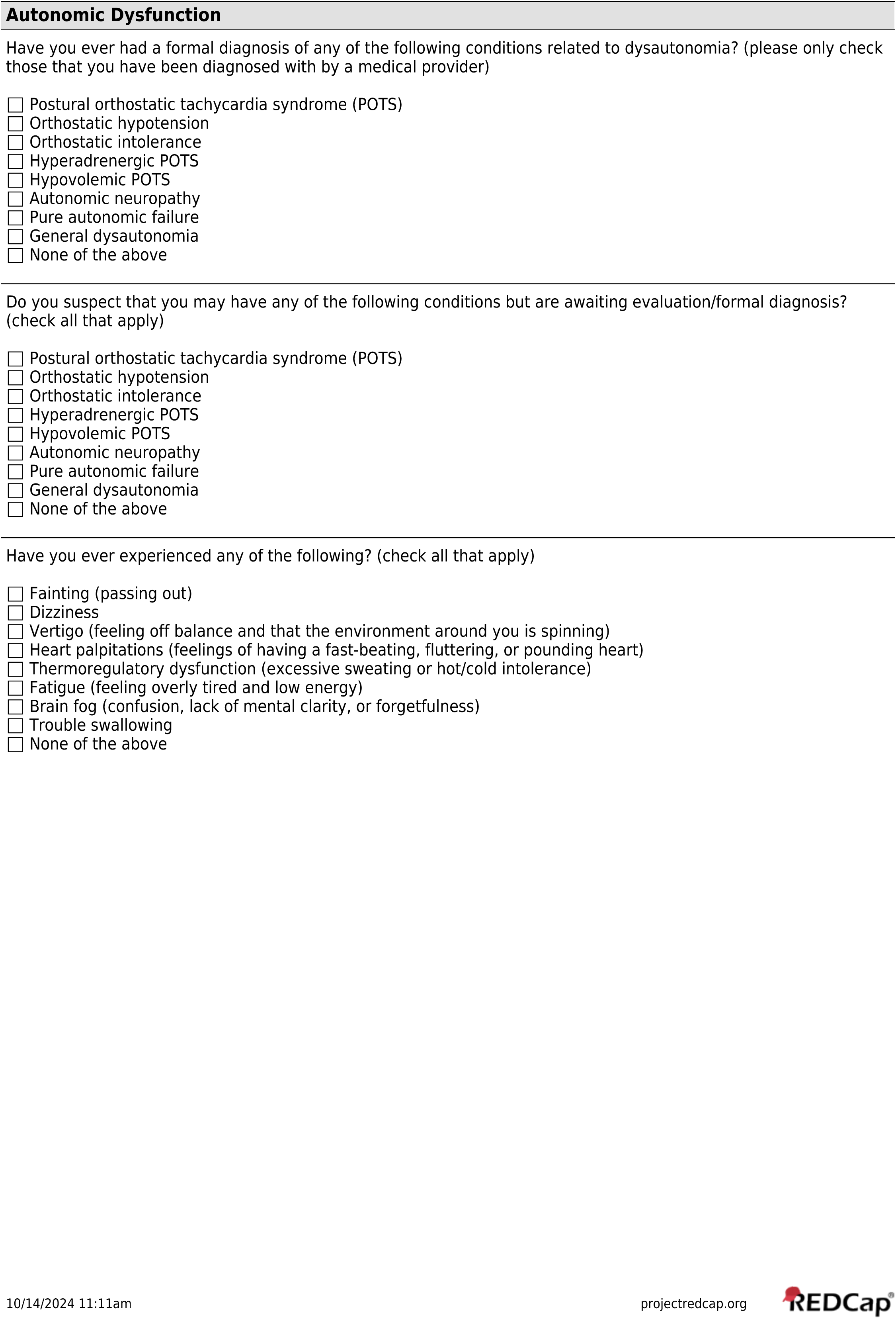
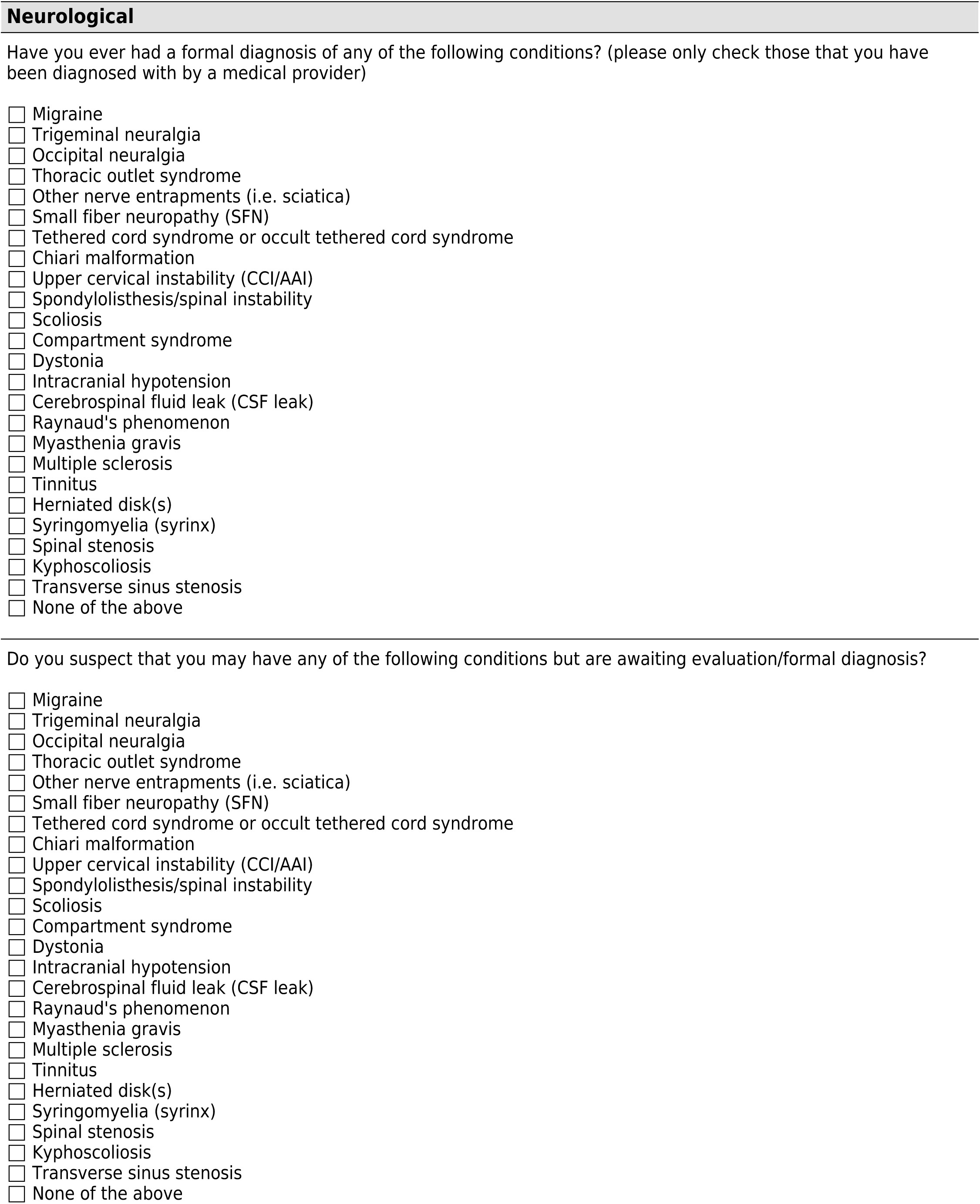
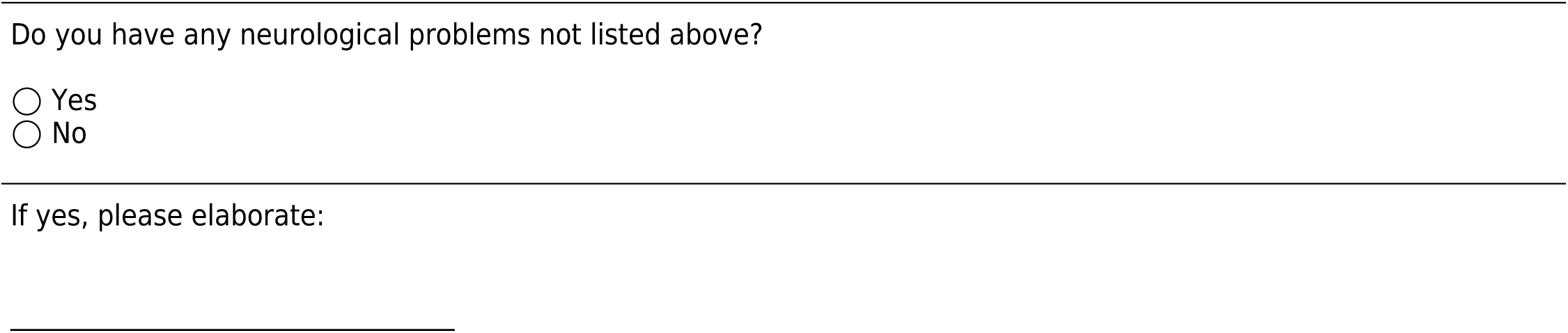
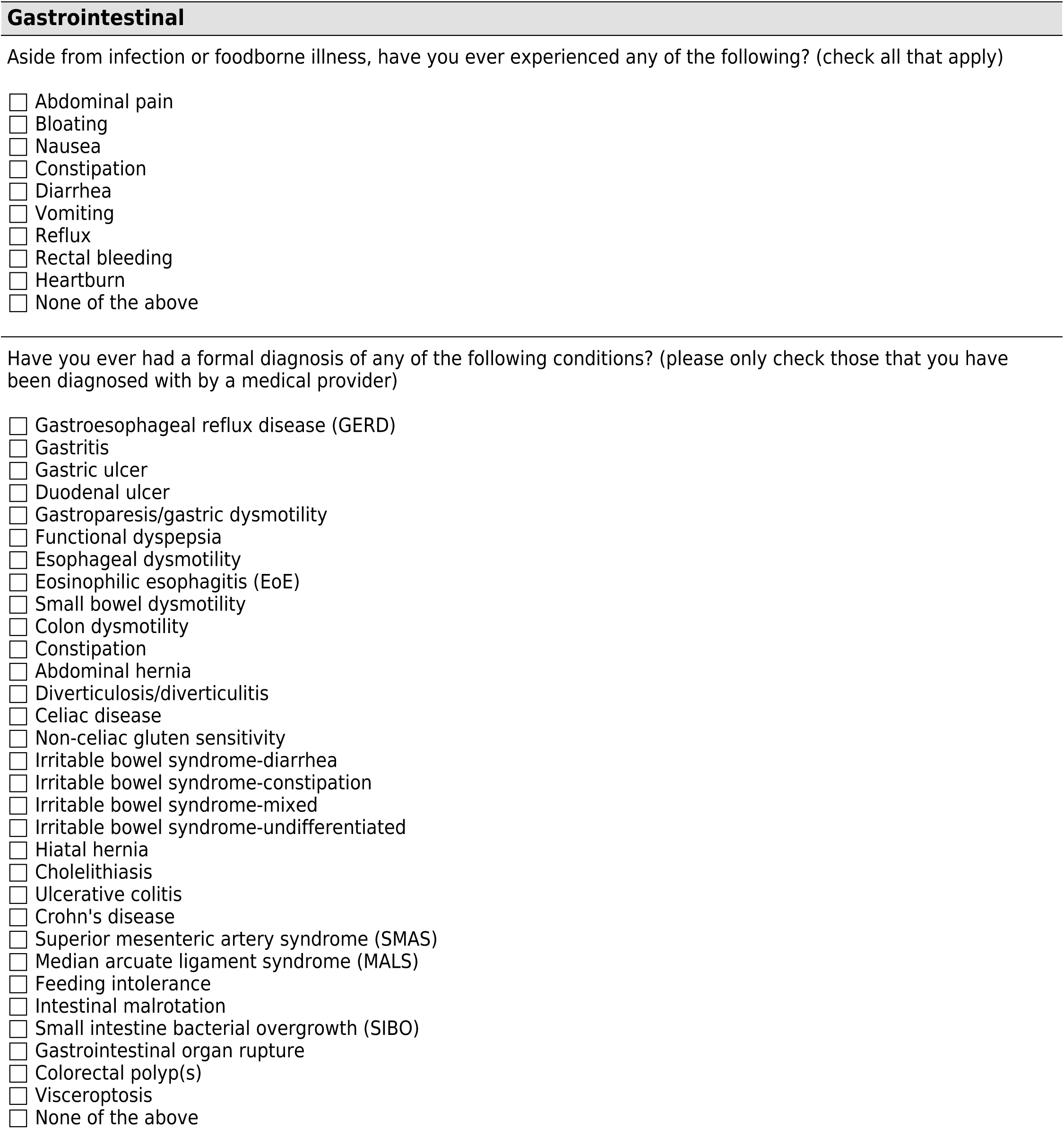
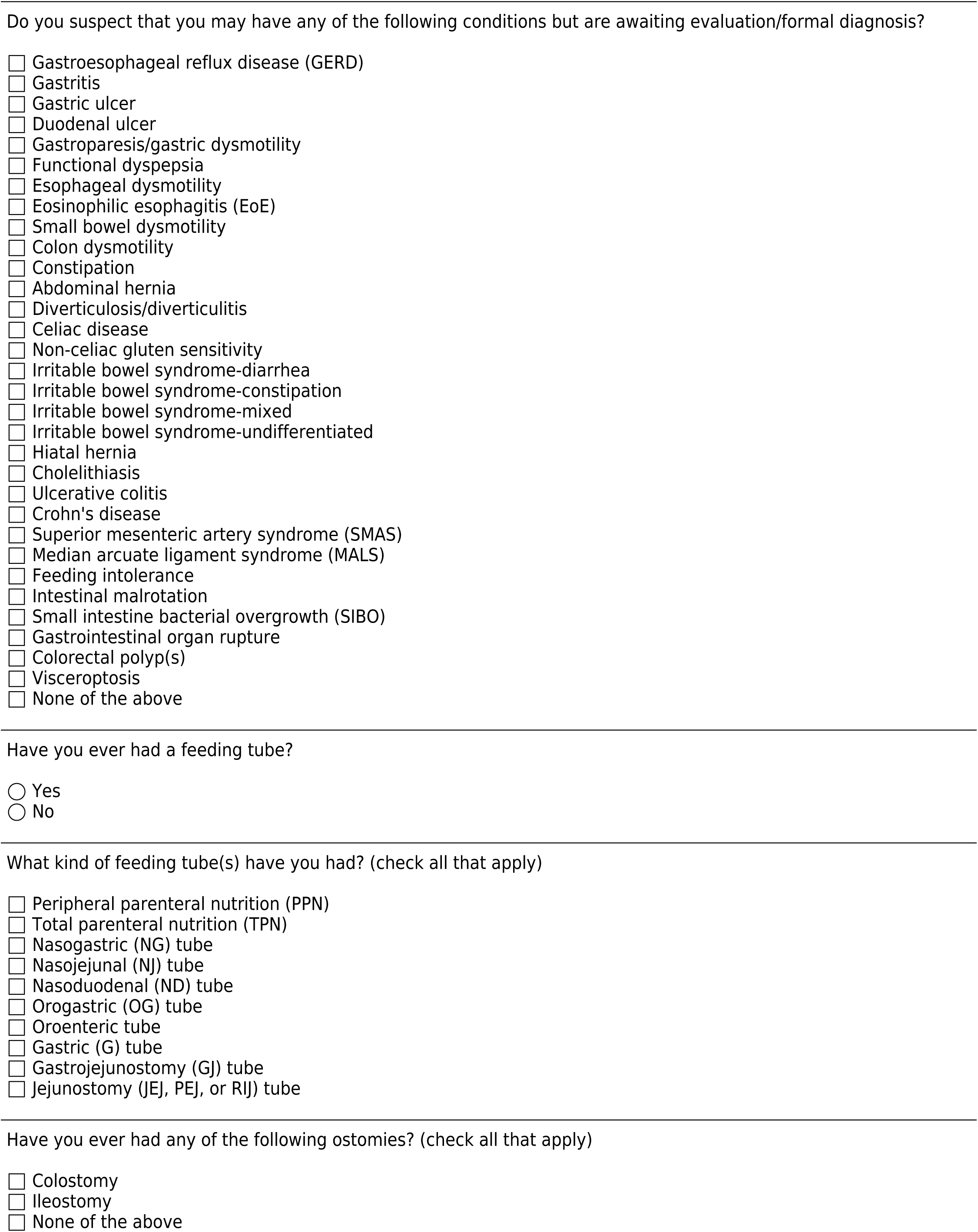
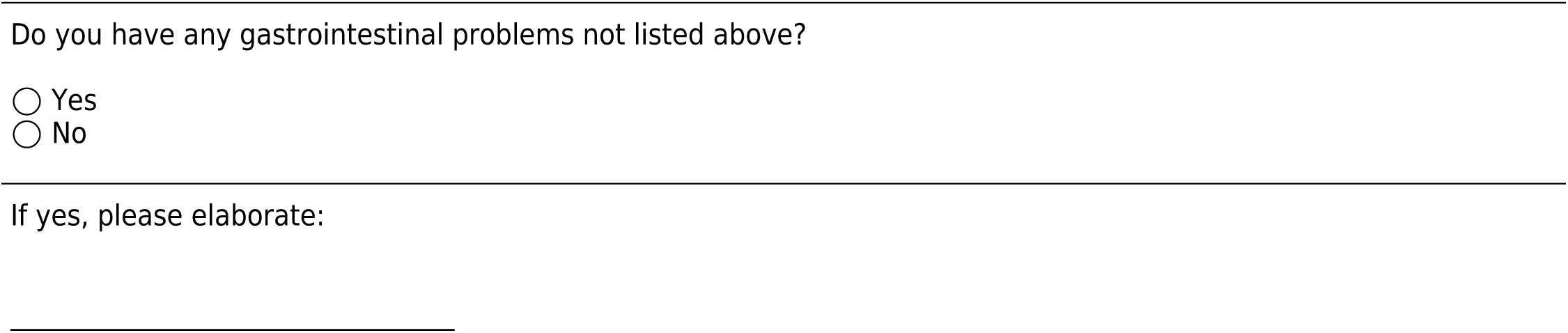
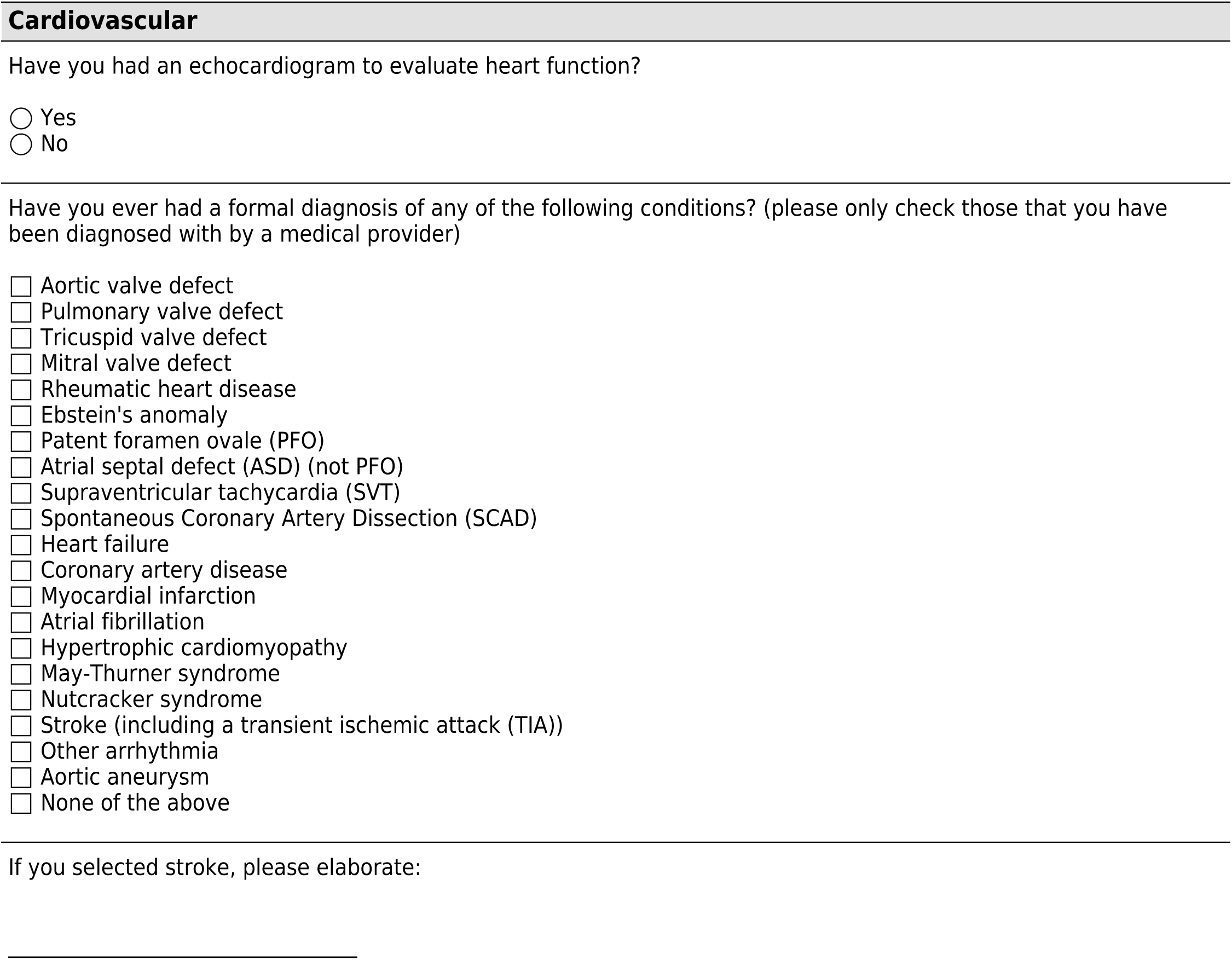
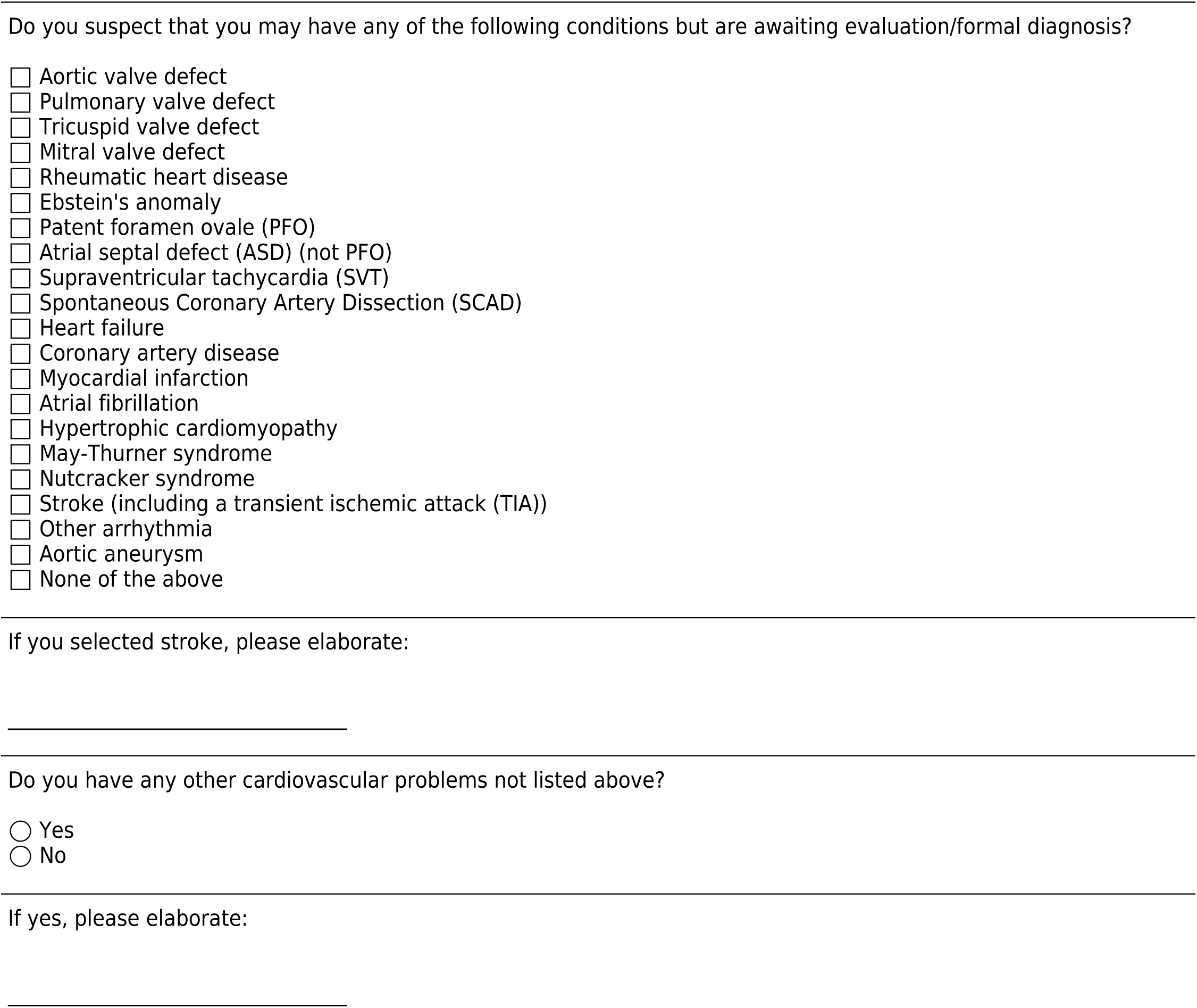
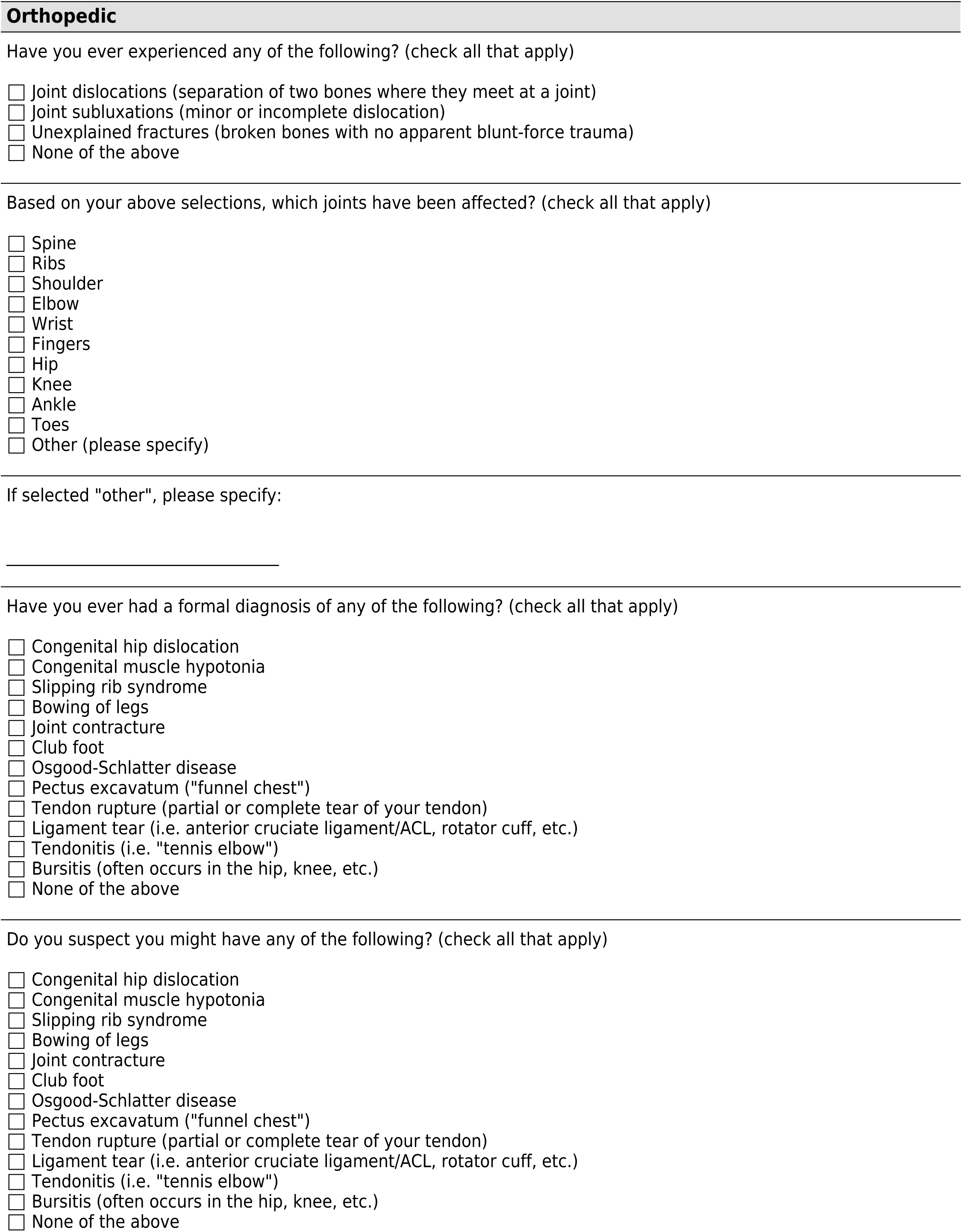
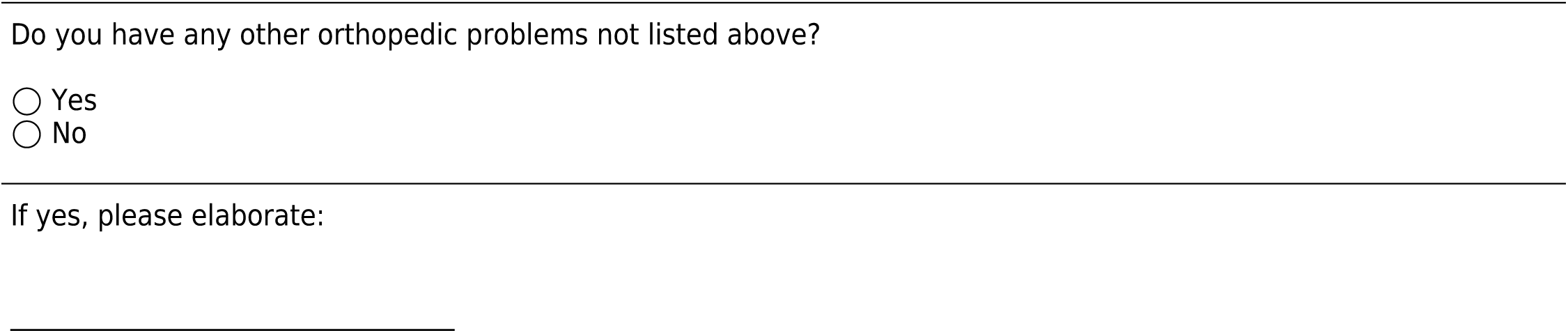
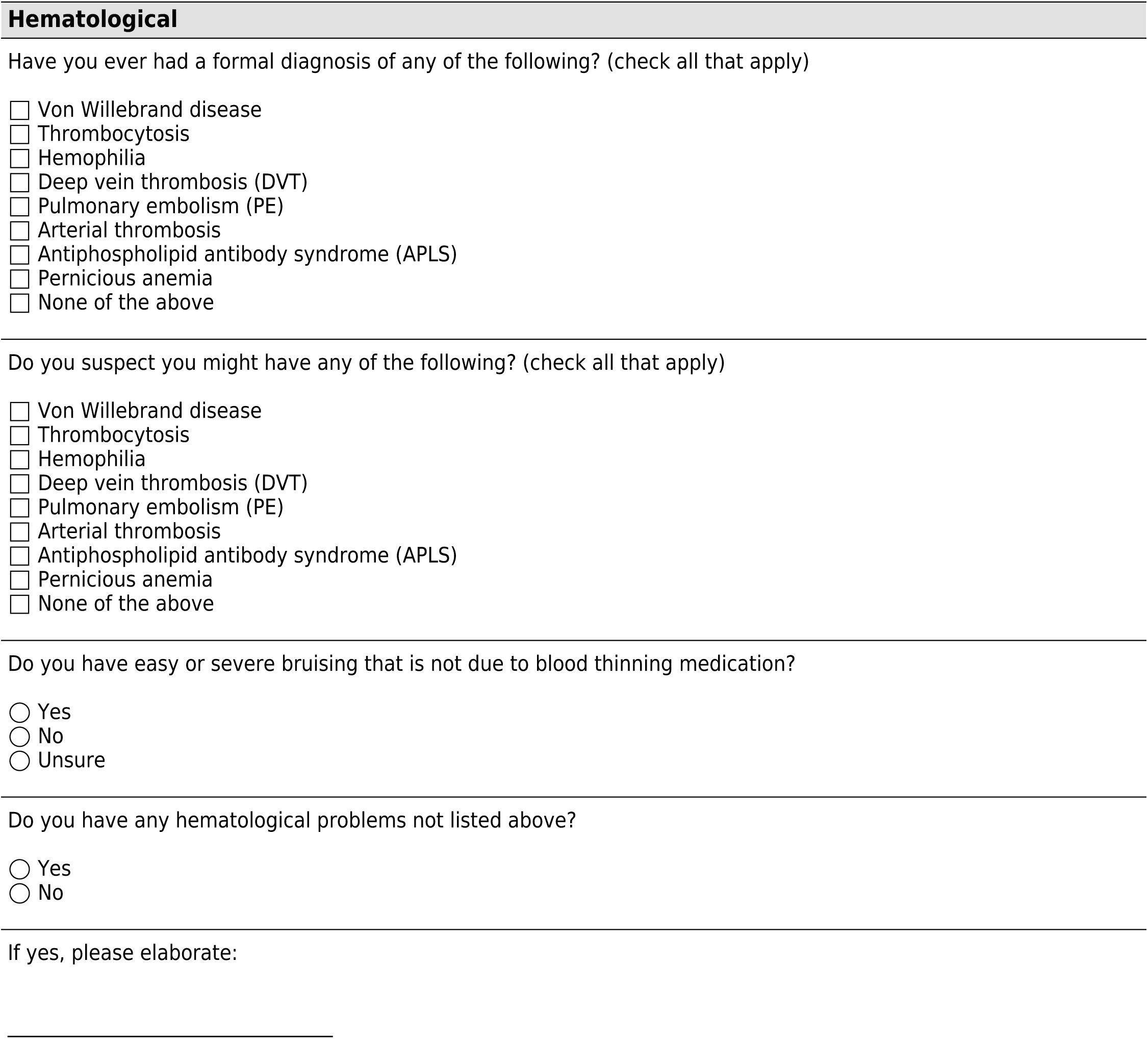
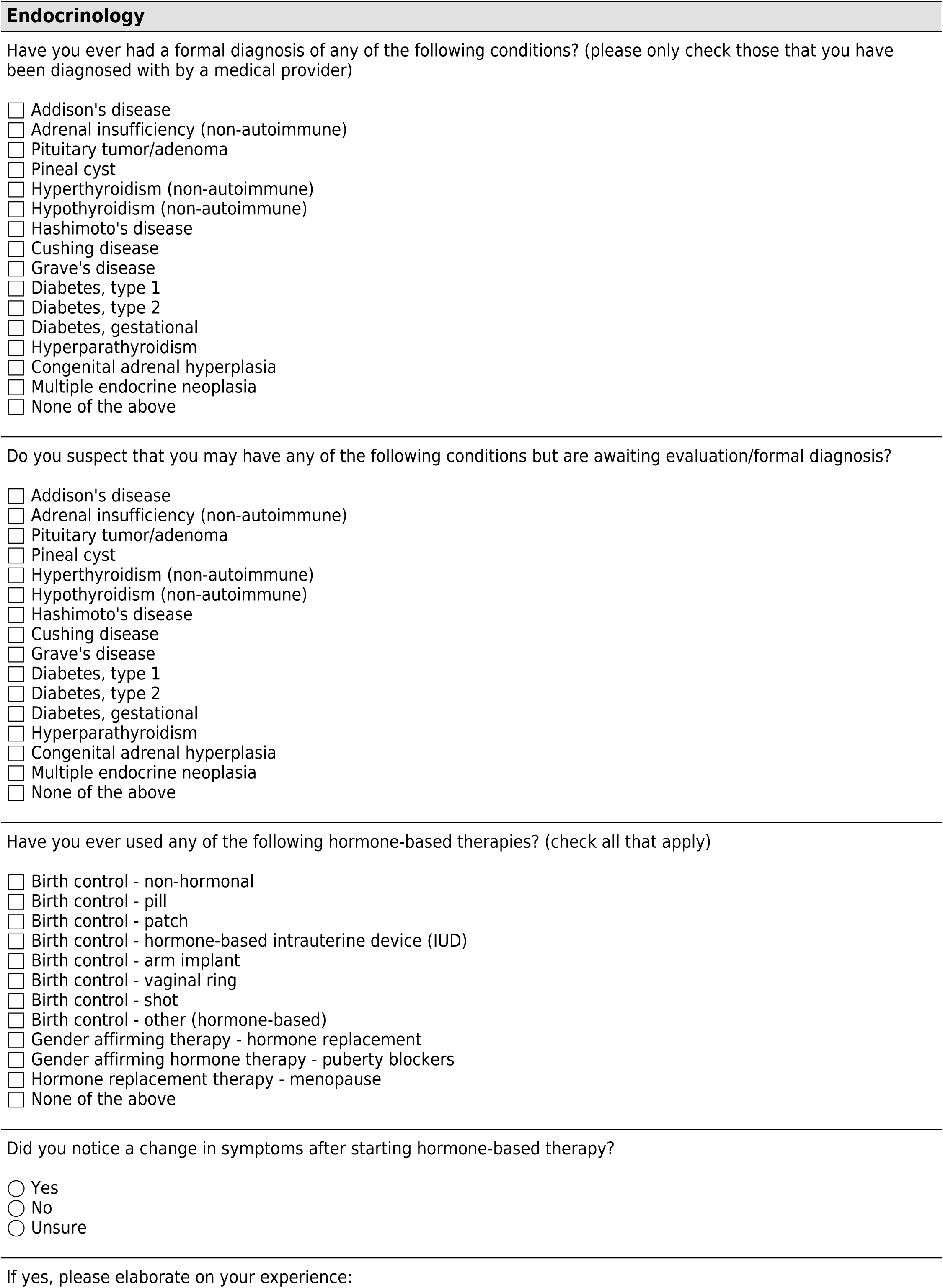
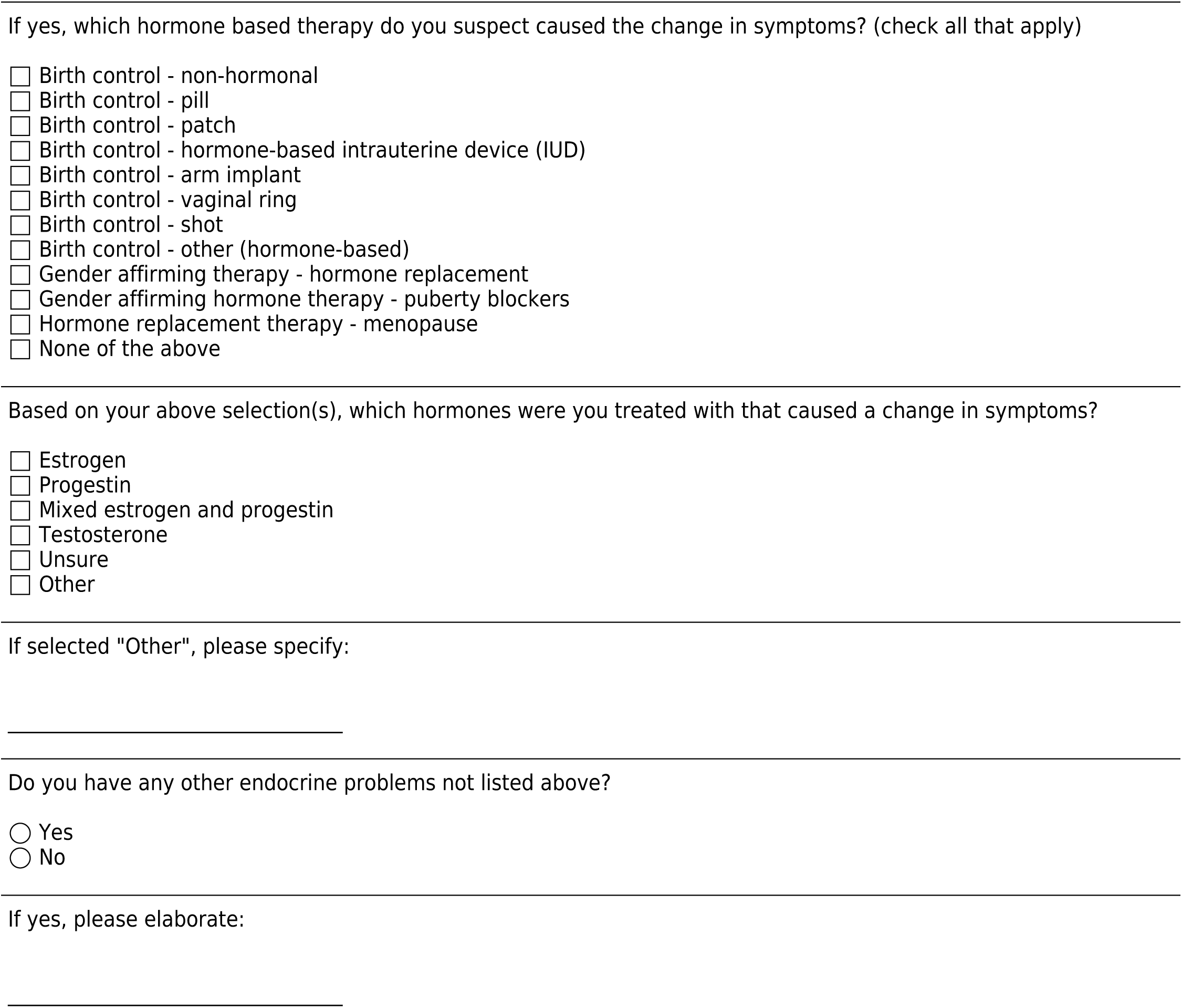
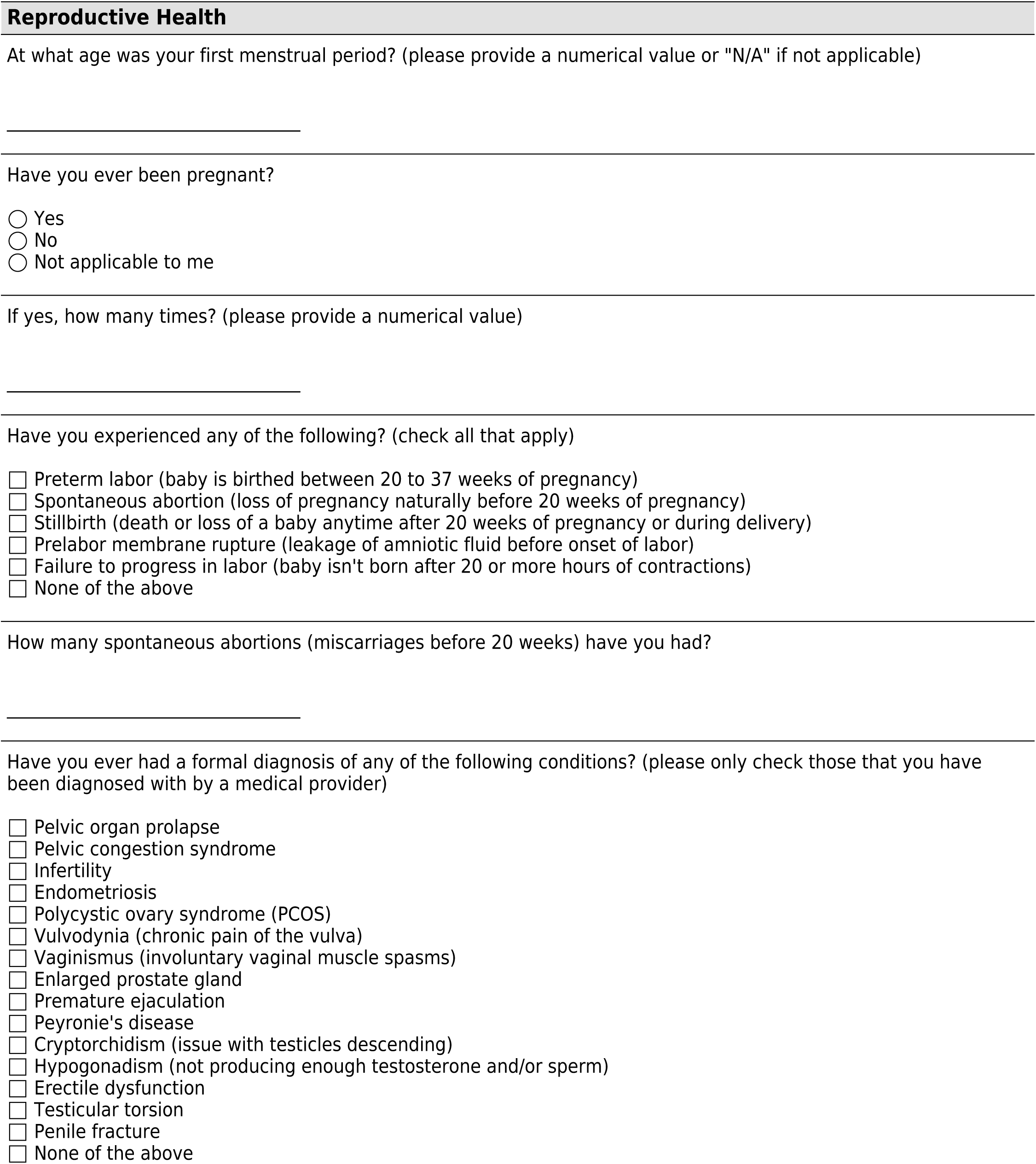
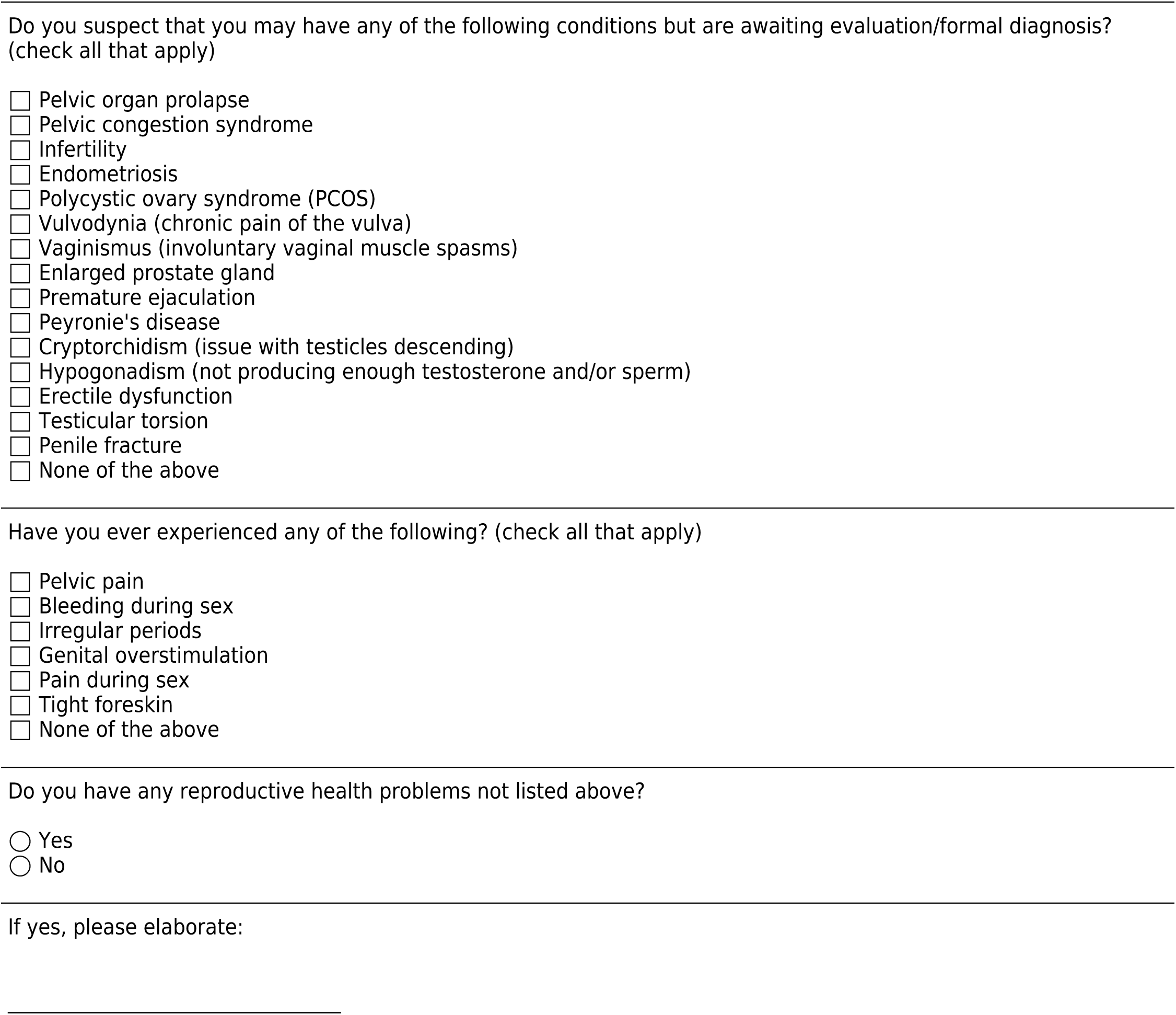
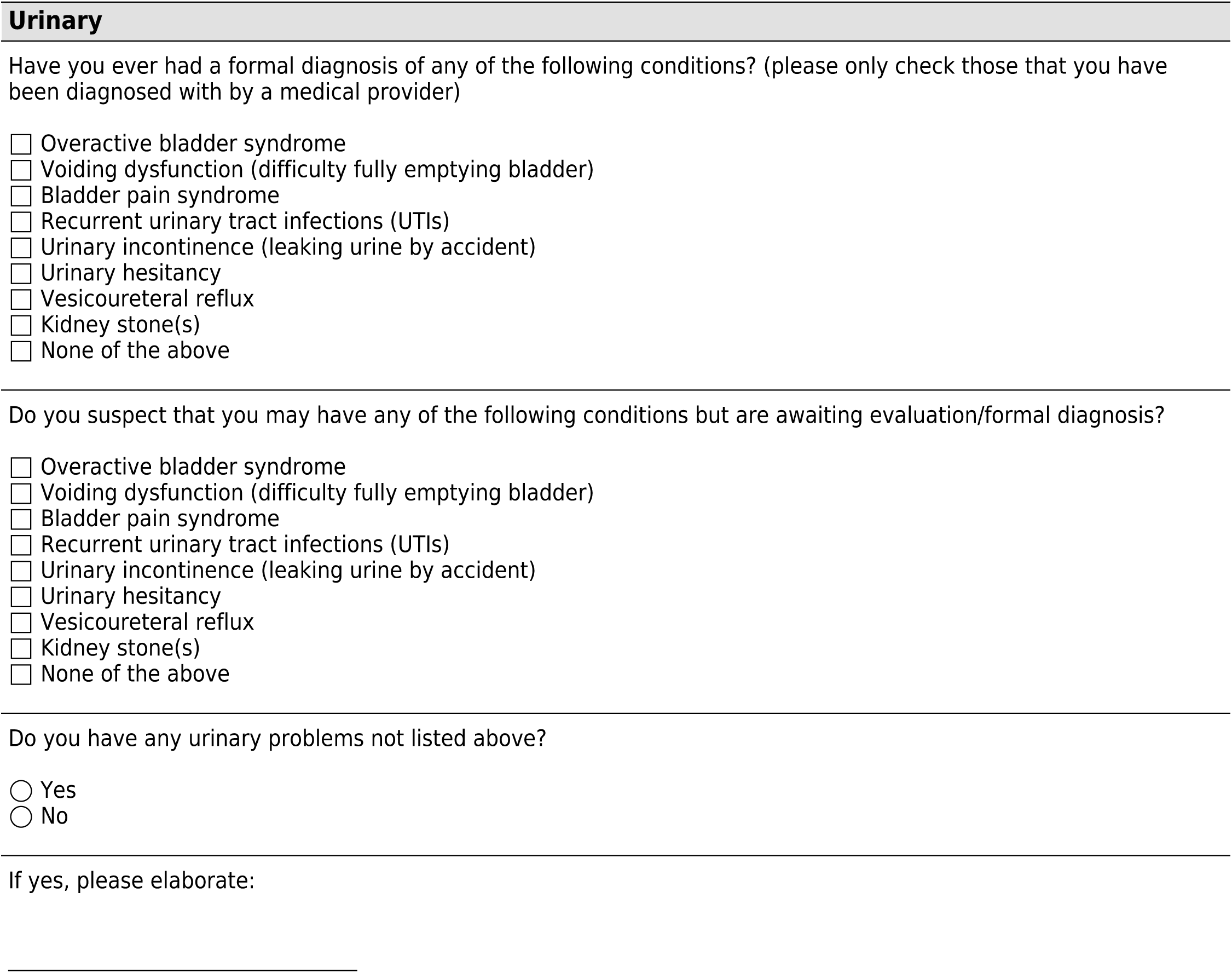
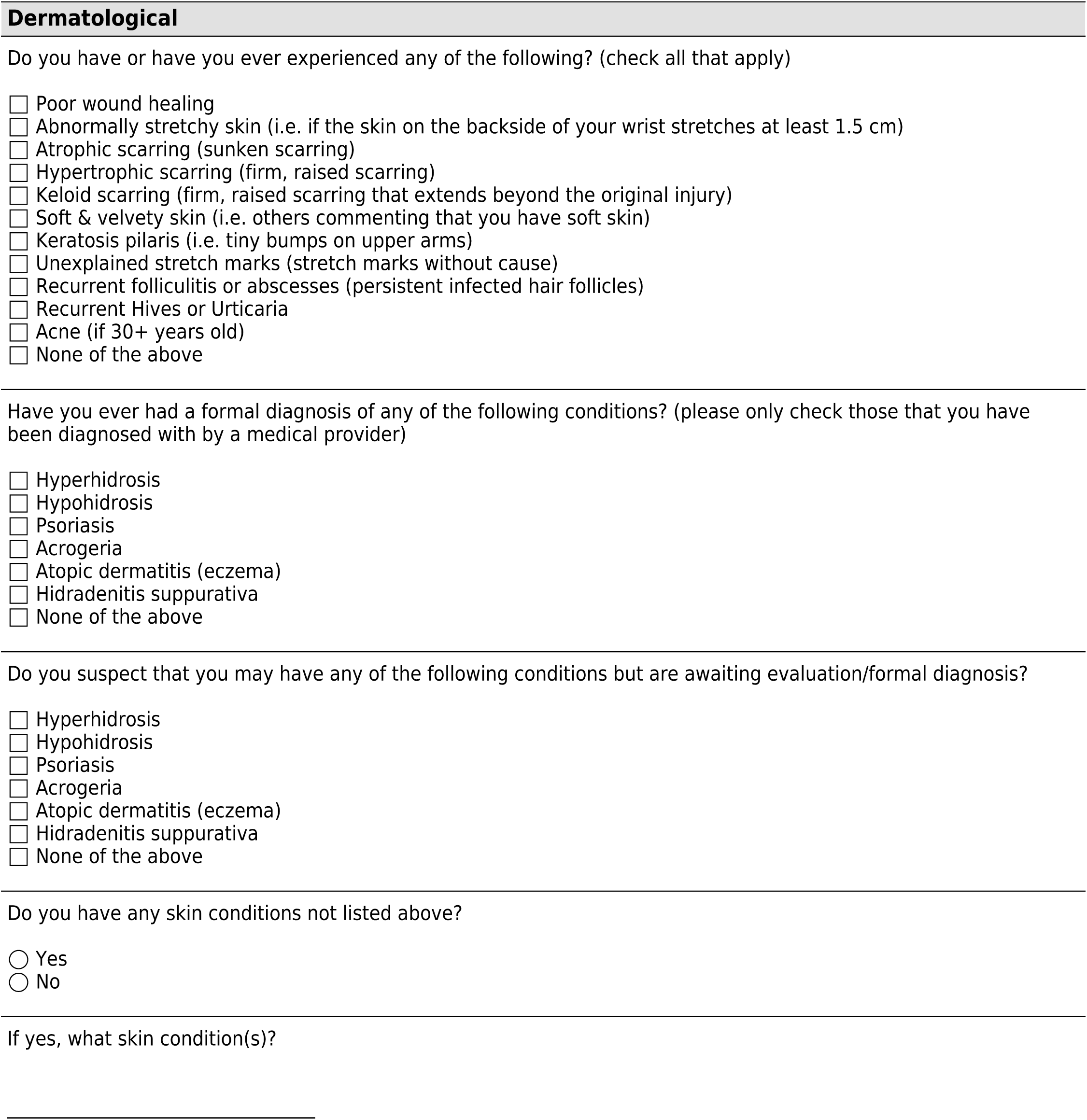
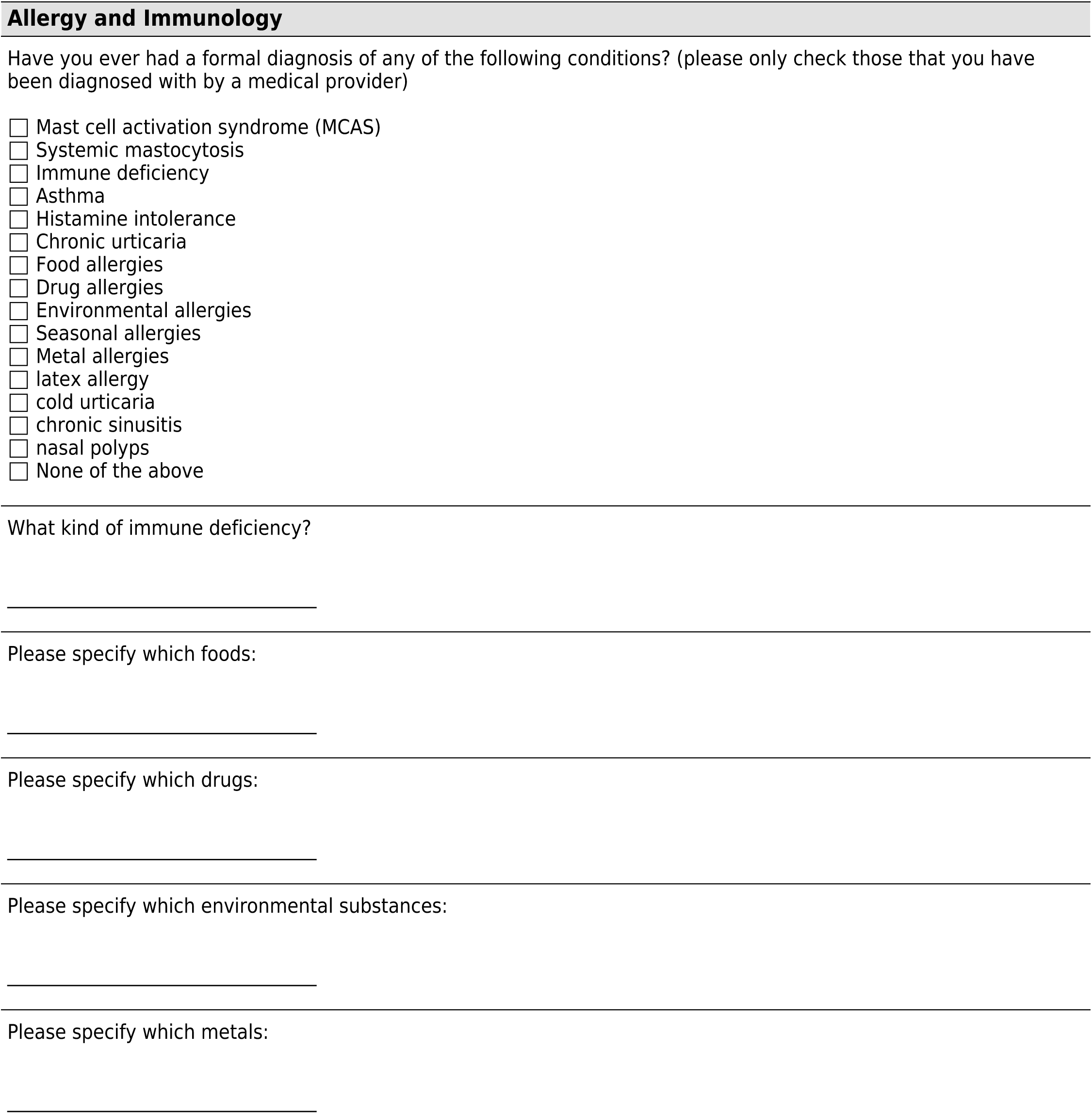
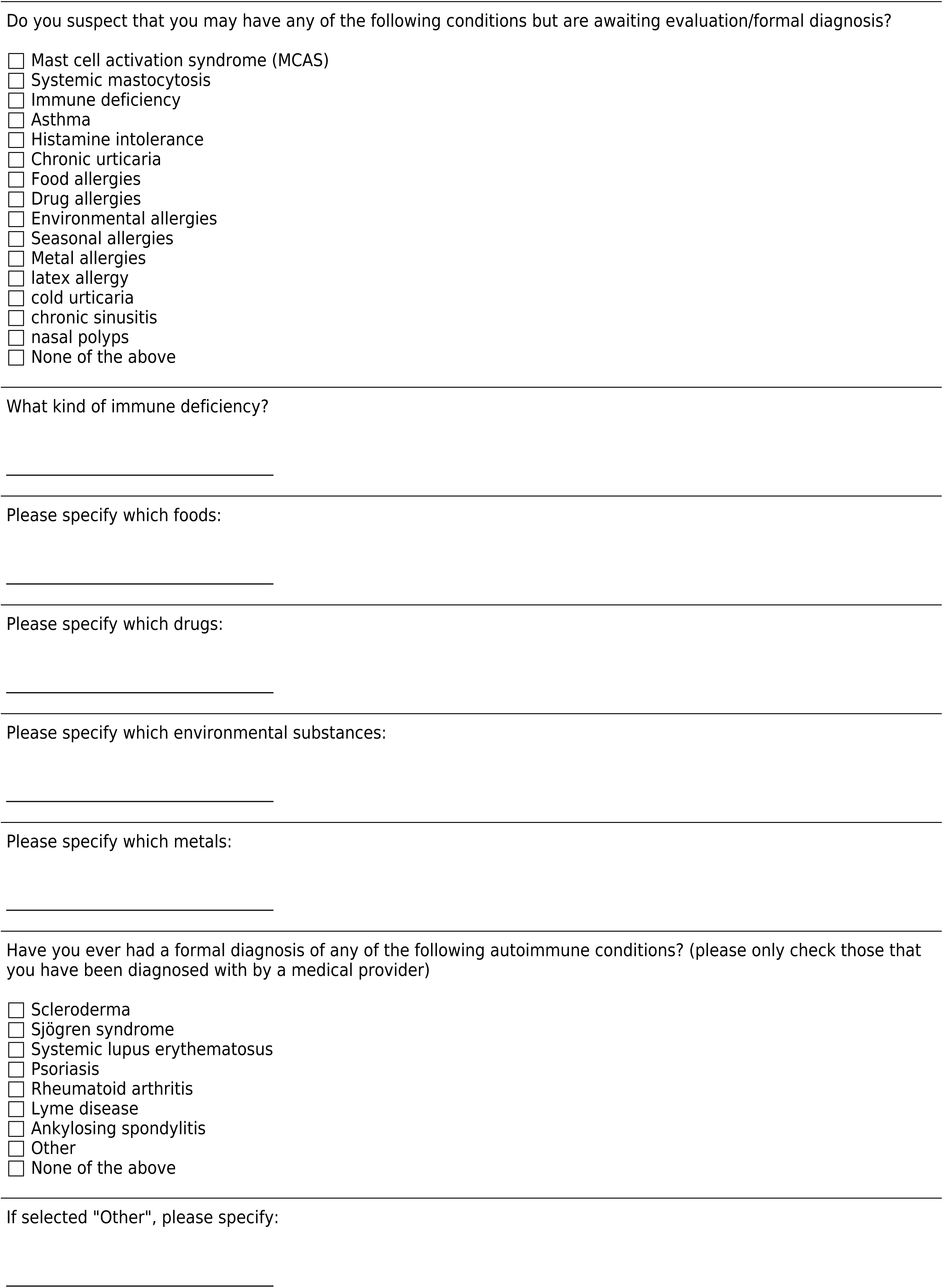
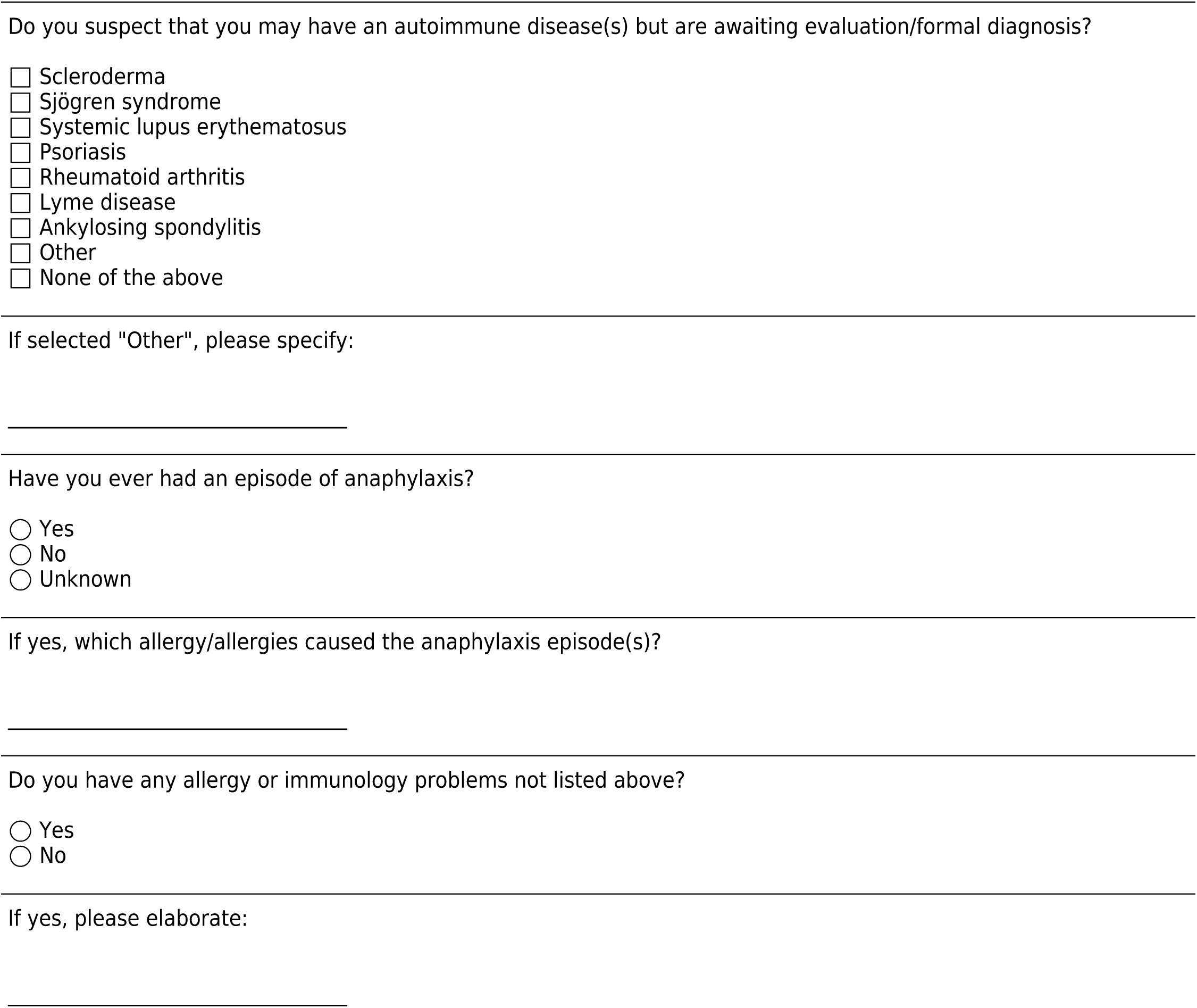
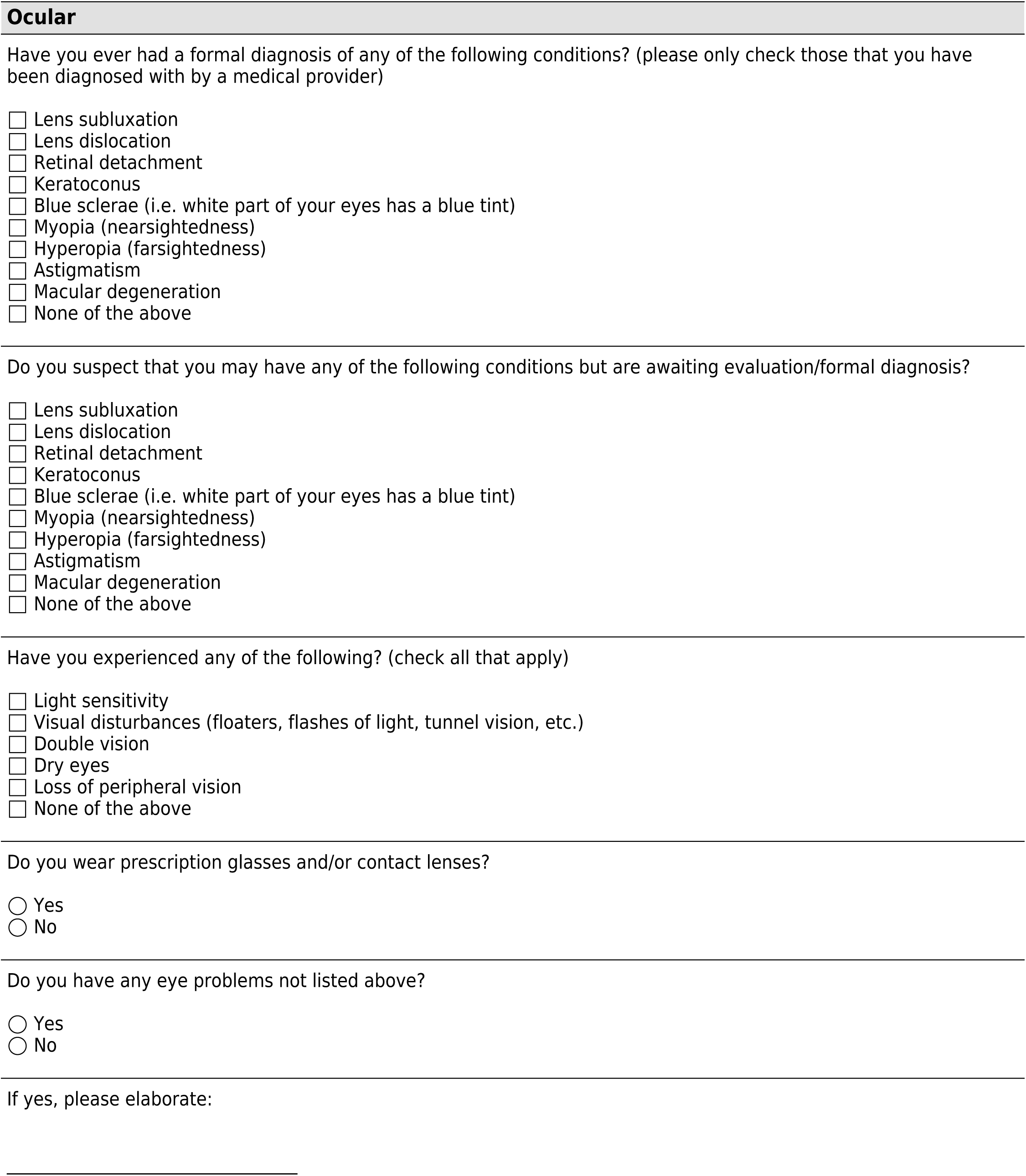
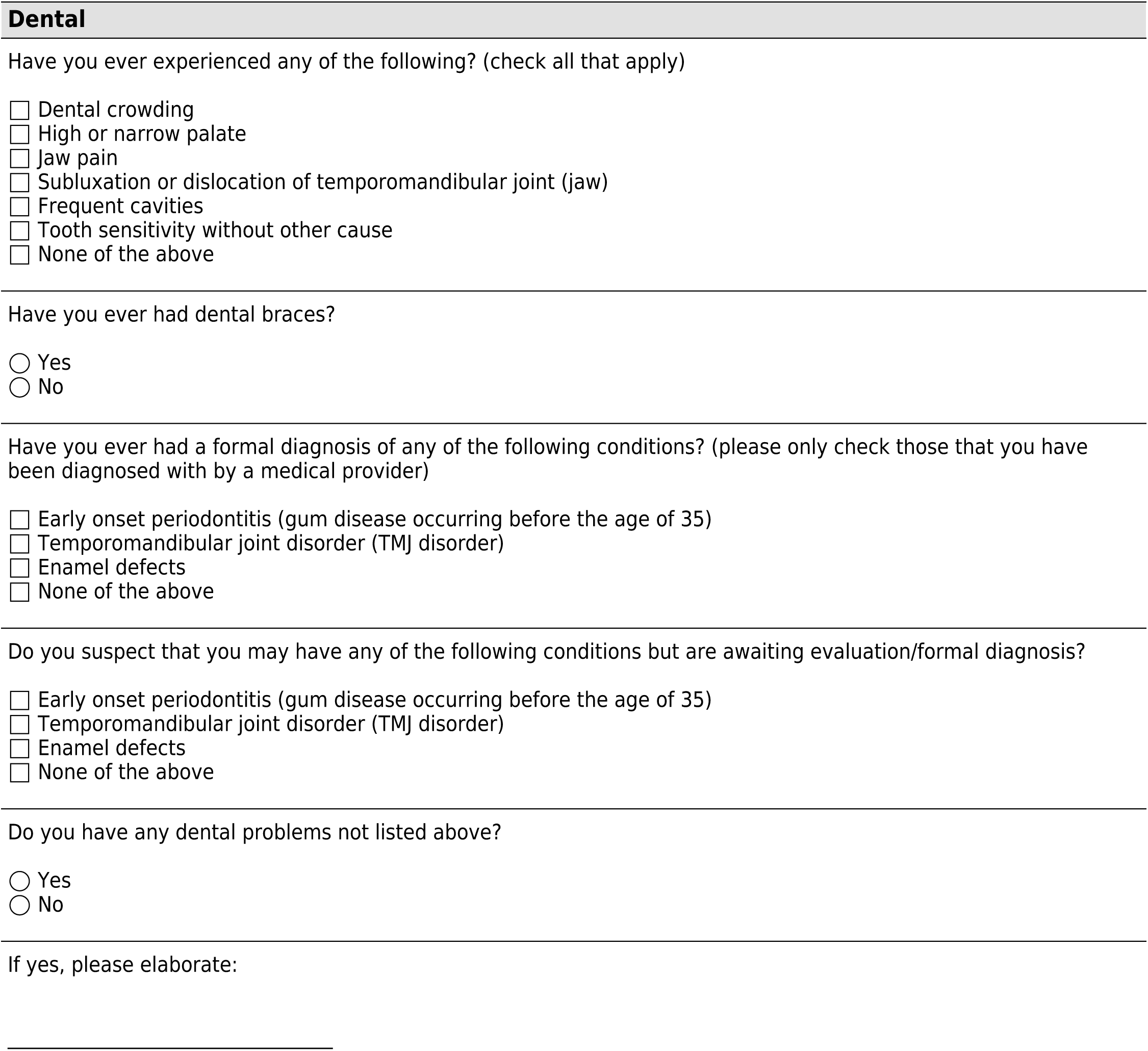
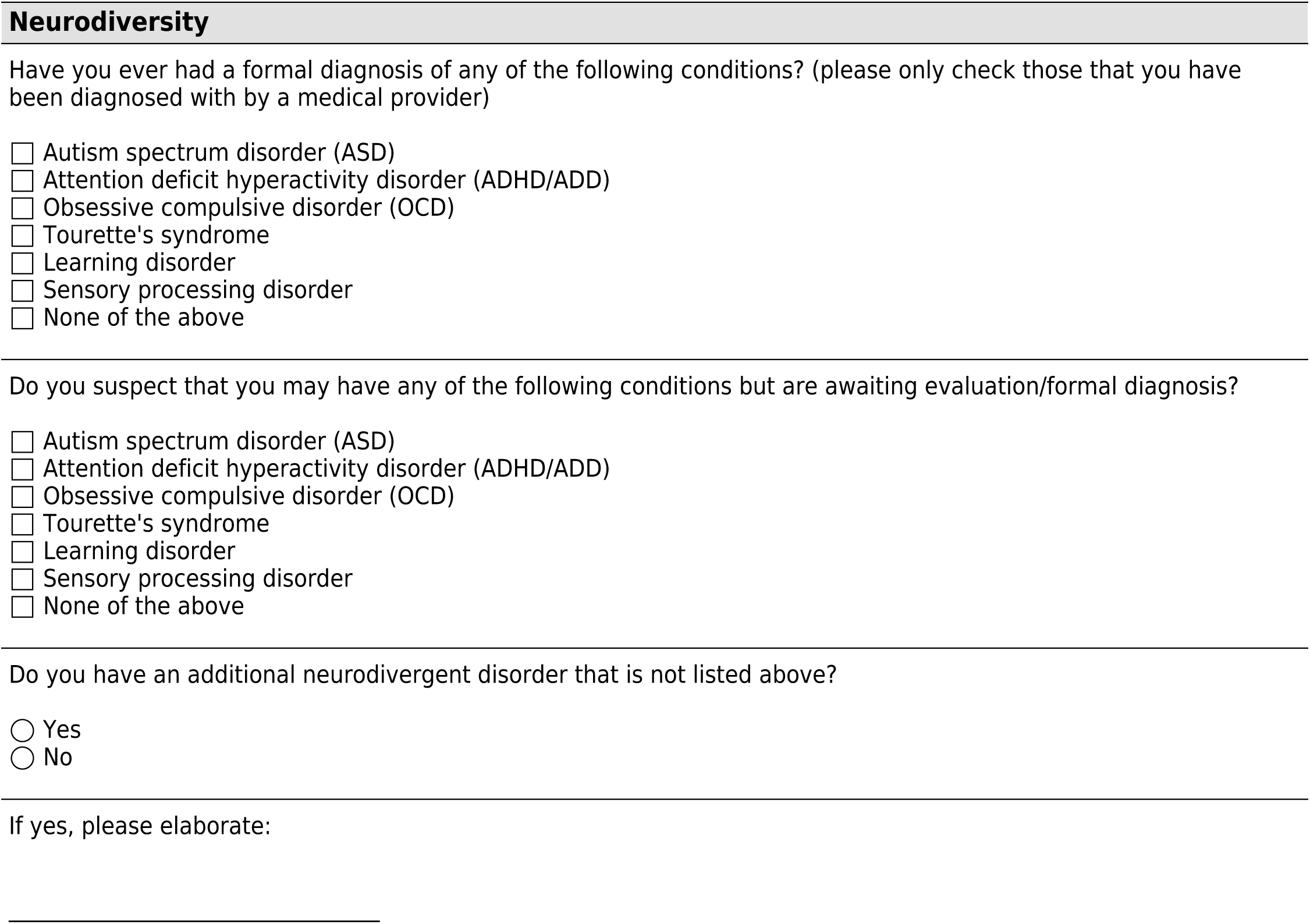
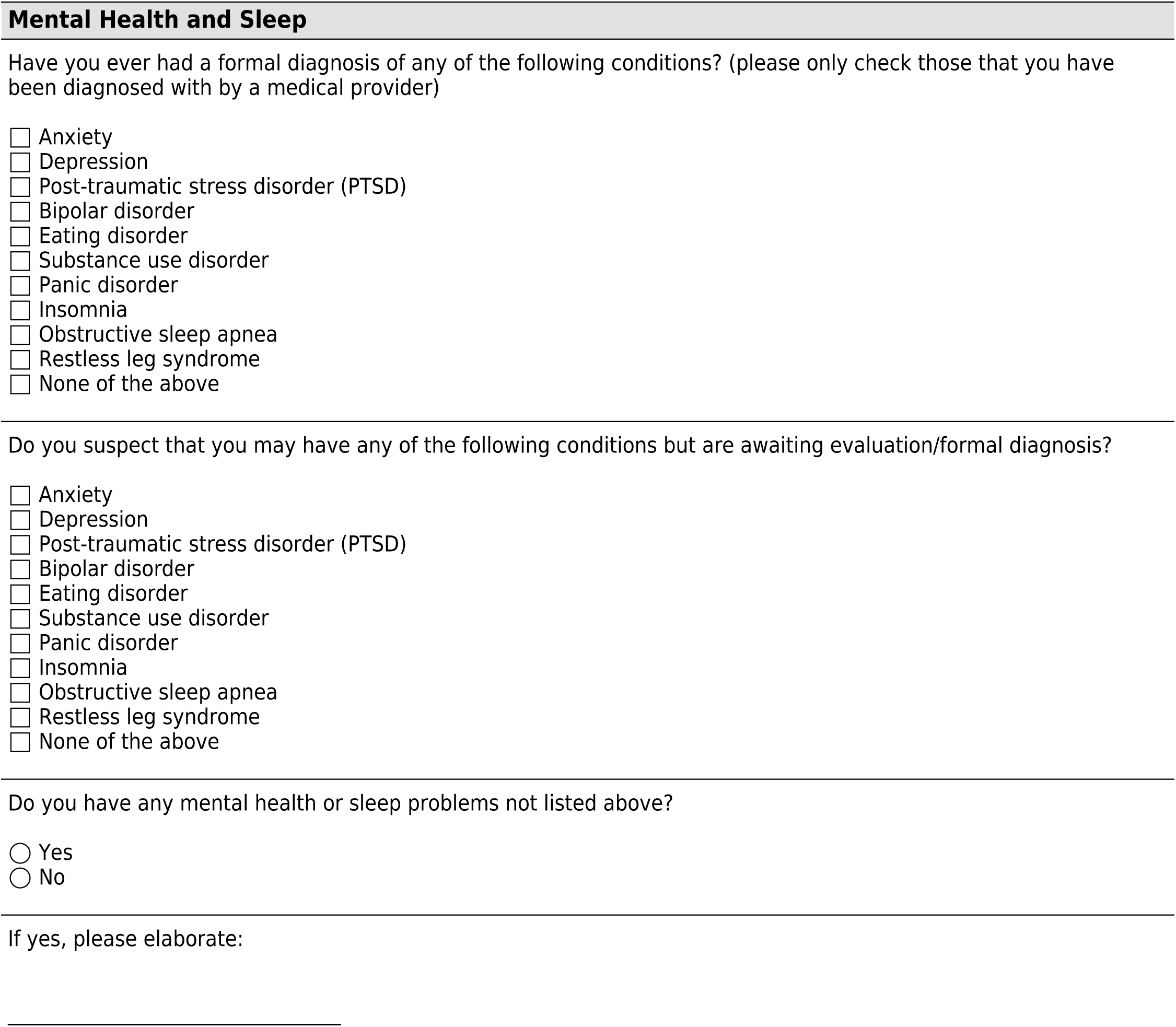
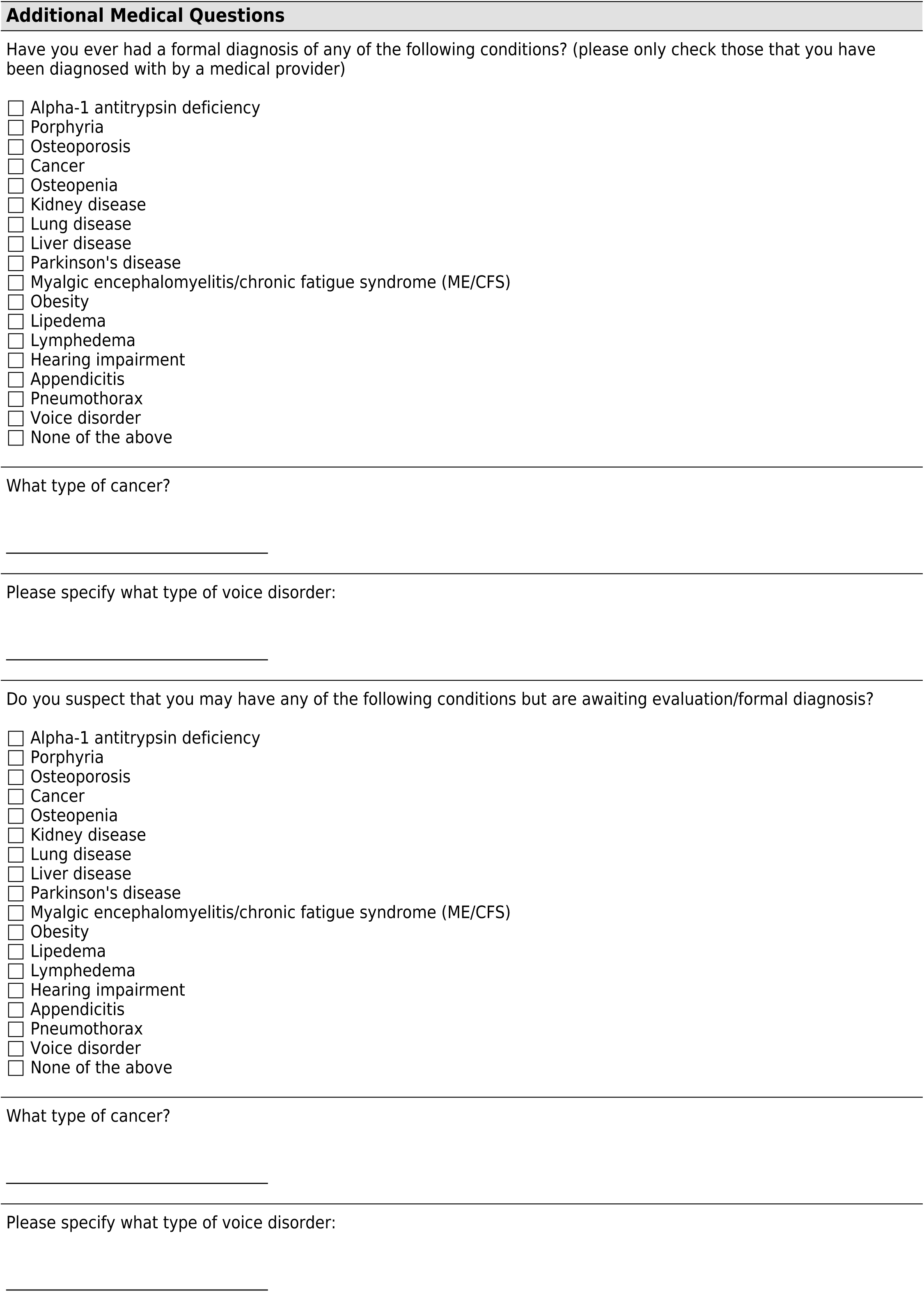
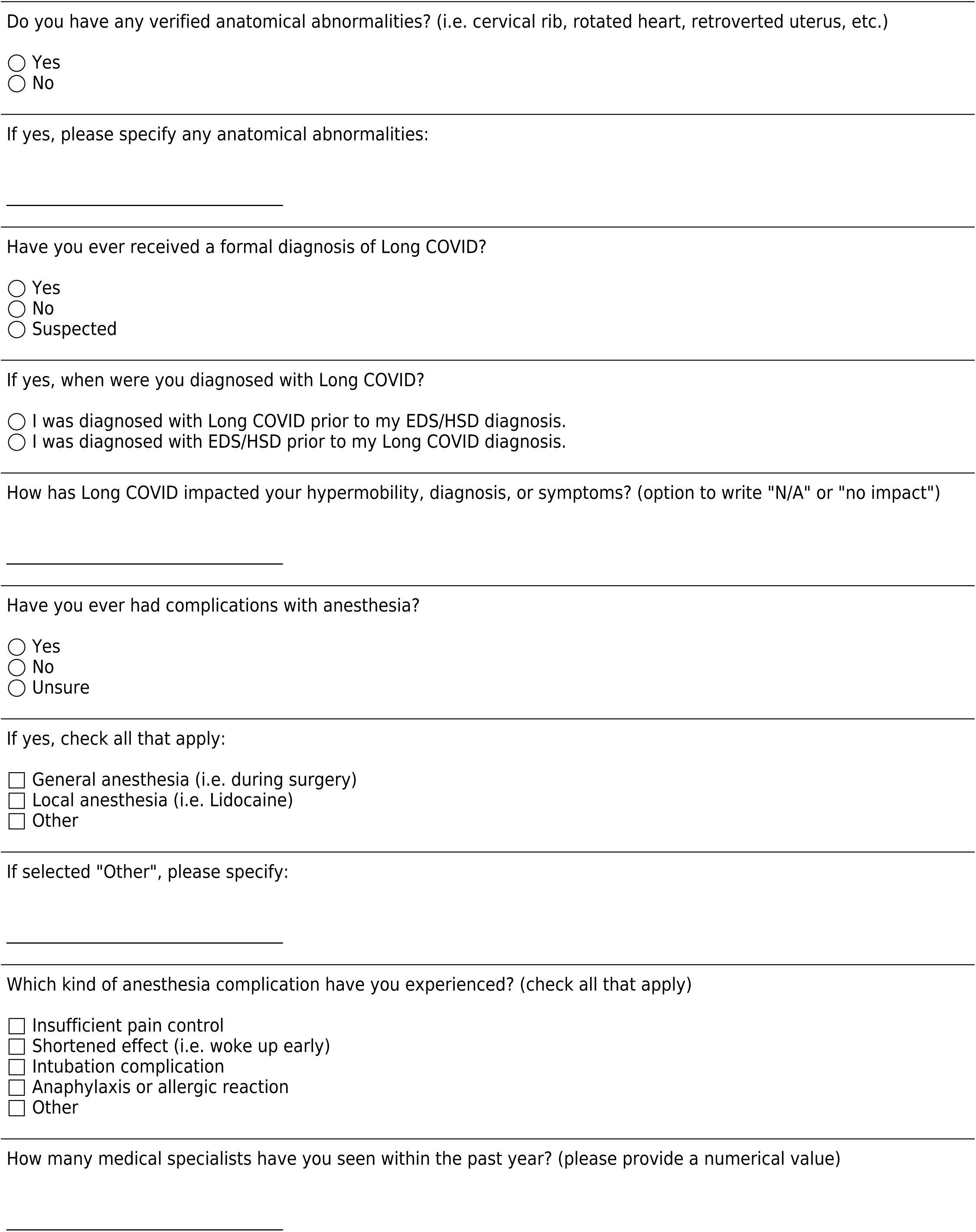
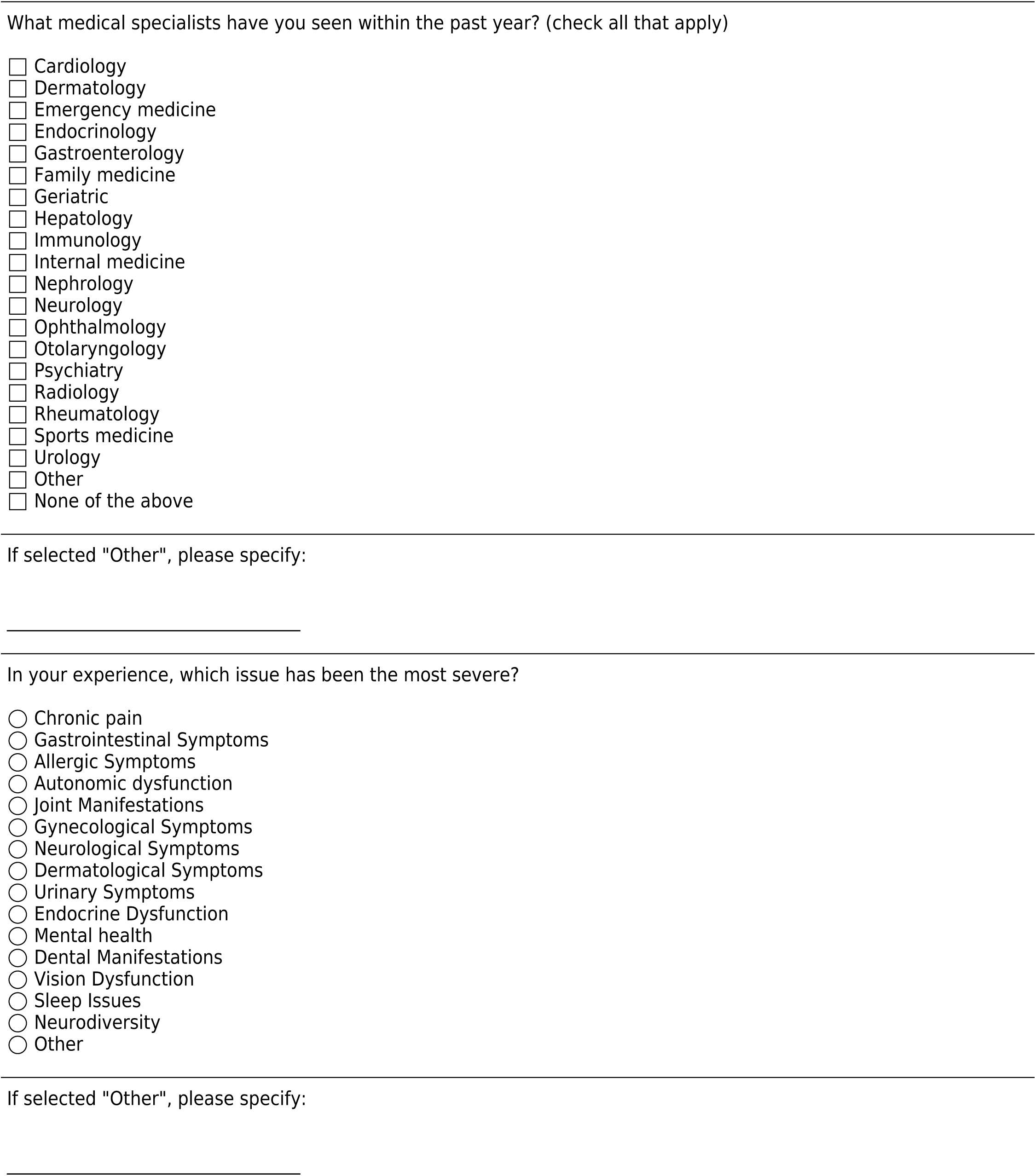
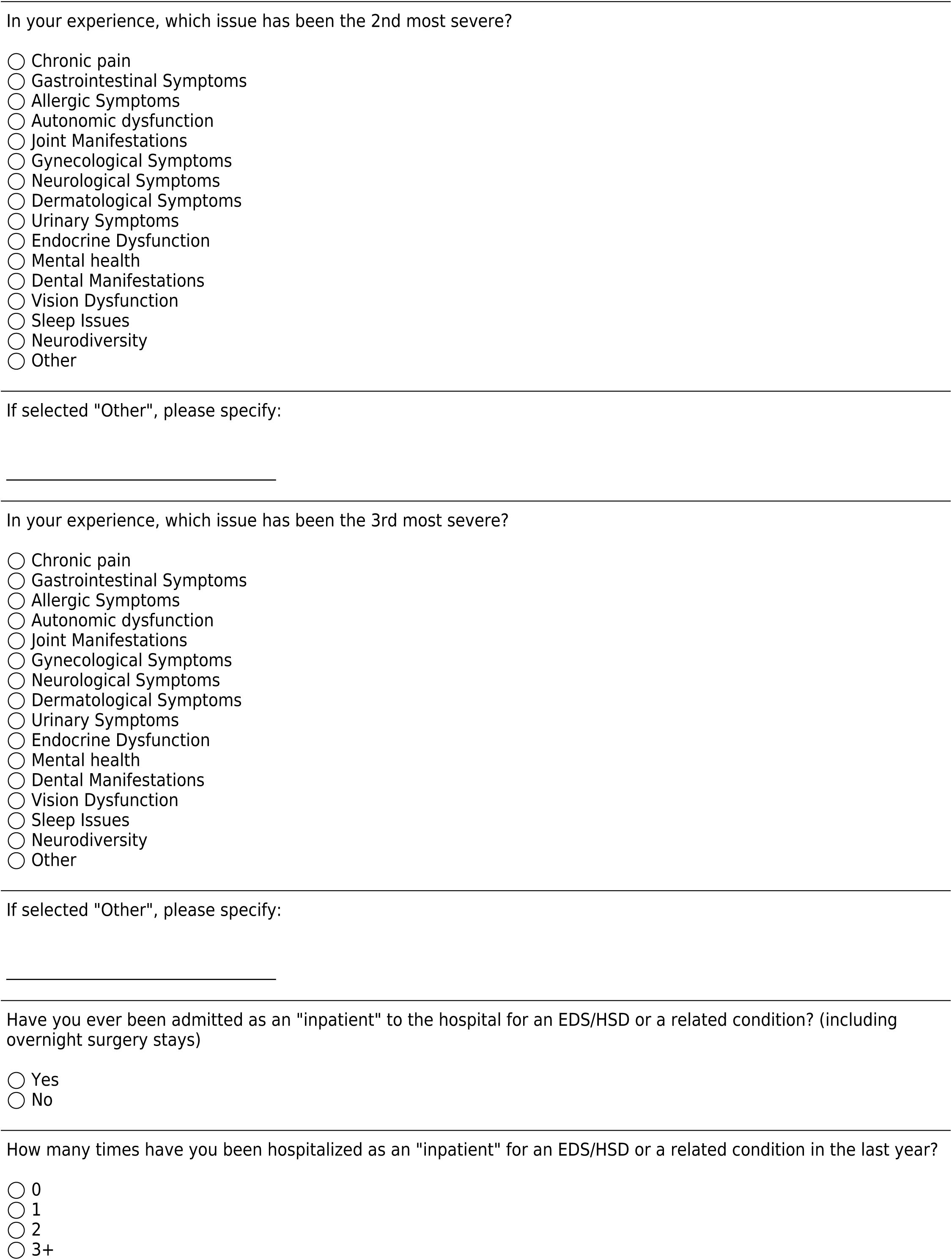
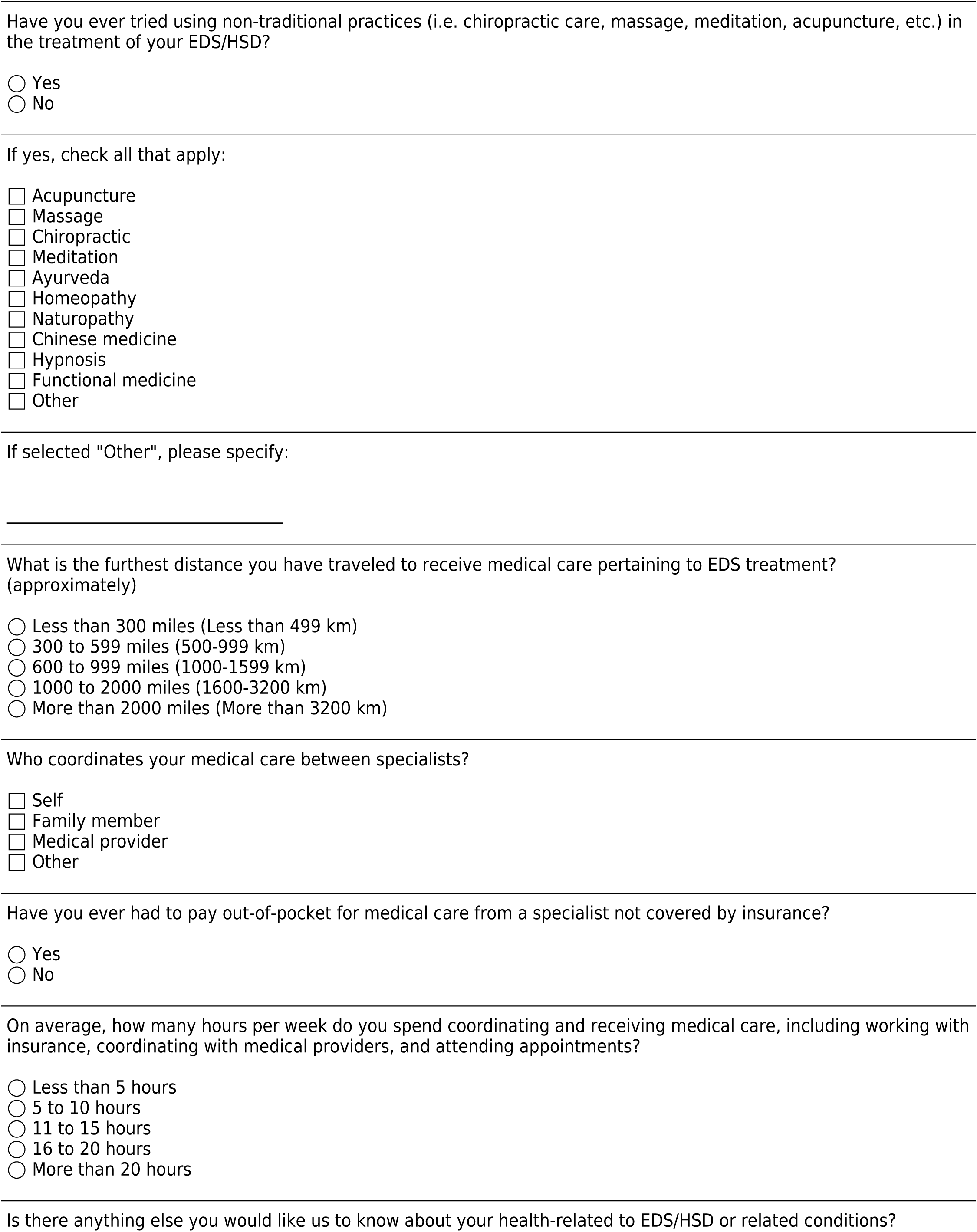
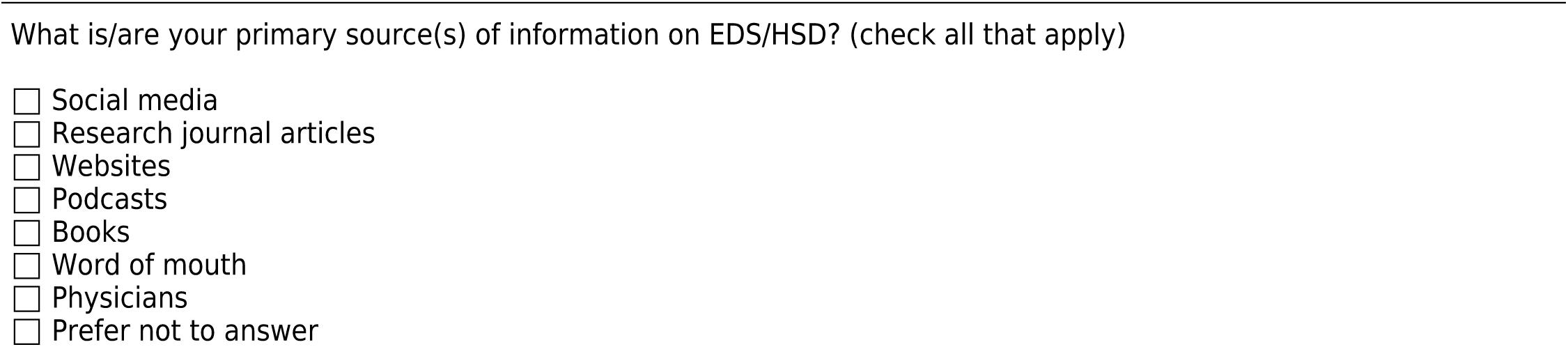

**Supplementary Figure 2.**
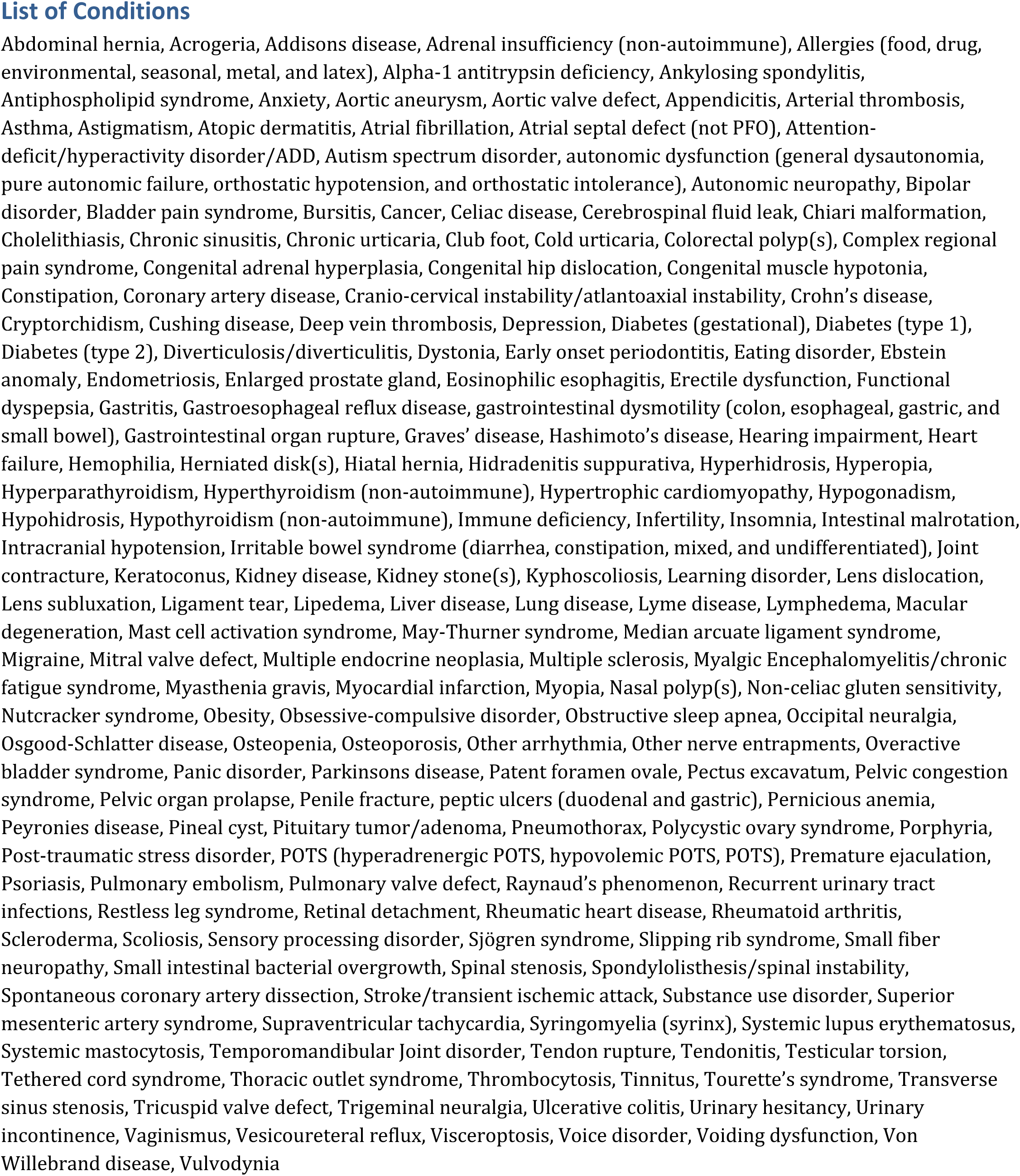

